# Estimating the effect of cesarean delivery on long term childhood health across two countries

**DOI:** 10.1101/2020.12.15.20248167

**Authors:** Ayya Keshet, Hagai Rossman, Smadar Shilo, Shiri Barbash-Hazan, Guy Amit, Maytal Bivas-Benita, Chen Yanover, Irena Girshovitz, Pinchas Akiva, Avi Ben-Haroush, Eran Hadar, Arnon Wiznitzer, Eran Segal

## Abstract

**Background:** Previous studies have demonstrated cesarean delivery (CD) may influence a variety of long term childhood outcomes. However, assessing the impact of CD on long term outcomes is challenging, as conducting a randomized controlled trial is not feasible and inferring it from observational data may be biased.

**Methods:** Utilizing over a decade of data obtained from electronic health records of 737,904 births, with a mean child follow-up of 5.66 (SD 2.96) years, we defined and emulated a target trial to estimate the effect of CD on several predefined long term pediatric outcomes. Causal effects were estimated using pooled logistic regression and standardized survival curves, leveraging data breadth to account for potential confounders. Diverse sensitivity analyses were performed including replication of results in an external validation set from the UK including 625,044 births with a mean child follow-up of 6.77 (SD 4.54) years.

**Findings:** 10-year risk difference (95% CI) between infants born by CD versus vaginal delivery were 0.74% (−0.06, 1.52) for atopy, 0.64% (0.31, 0.98) for asthma and 0.47% (−0.32, 1.28) for allergy. Increased risk for these outcomes was also observed in the UK cohort. In addition, an average treatment effect of 0.10 (0.07-0.12) on body mass index (BMI) z-scores at age 5 years old and 0.92 (0.68-1.14) on the number of respiratory infection events until 5 years of age were observed for CD. For other examined outcomes, including autism and autoimmune diseases, an increased risk for infants born by CD was not observed consistently when employing different methods and across both countries.

**Interpretation:** Our findings add to a growing body of evidence on the long term effects of CD on pediatric morbidity, and may assist parents and clinicians in the decision to perform CD when not medically indicated. Furthermore, this study may pave the way to future research on the mechanisms underlying these effects and intervention strategies targeting them.

**Funding:** E.S. is supported by the Crown Human Genome Center; Larson Charitable Foundation New Scientist Fund; Else Kroener Fresenius Foundation; White Rose International Foundation; Ben B. and Joyce E. Eisenberg Foundation; Nissenbaum Family; Marcos Pinheiro de Andrade and Vanessa Buchheim; Lady Michelle Michels; Aliza Moussaieff; and grants funded by the Minerva foundation with funding from the Federal German Ministry for Education and Research and by the European Research Council and the Israel Science Foundation. H.R. is supported by the Israeli Council for Higher Education (CHE) via the Weizmann Data Science Research Center and by a research grant from Madame Olga Klein – Astrachan.

**Research in context:** *Evidence before this study:* Previous studies have investigated the relationship between CD and long term pediatric health. However, the majority of studies thus far have focused on the association of CD with pediatric outcomes, without accounting for possible confounding factors that might affect these associations. In addition, some of these studies were based on relatively small cohorts from a single geographic location, with a short follow-up period.

*Added value of this study:* In this study we emulated a target trial on a large nationwide dataset, to estimate the causal effect of CD on several long term pediatric health outcomes. Further, we validated our results using several sensitivity analyses and through replication in an external validation set originating from a different country and ethnicity.

*Implications of all the available evidence:* Given the increasing incidence of CD, a deeper understanding of its long term health effects on women and children is crucial. Our findings that CD confers an increased risk for several long term pediatric health outcomes contribute to the ongoing discussion on the optimal rate of CD, and may assist parents and caregivers in the decision to perform CD when not medically indicated. Moreover, studies on the mechanisms mediating these effects have the potential to inform novel prevention strategies.

## Introduction

Medically indicated cesarean delivery (CD) is a life saving procedure for both mother and newborn, although it bears risk for short and long term pediatric adverse health outcomes ^1^. In the past decades, the incidence of CD increased annually by 4%, and is highly variable geographically, ranging from less than 5% of deliveries in southern Africa to almost 60% in some parts of Latin America ^2^. This rise, which also reflects an increase in CDs without a medical or obstetric indication ^3^, is the result of cultural, personal and medico-legal reasons, with limited consideration to the impact that mode of delivery may have on long-term pediatric health.

Previous studies have highlighted childhood obesity ^4^; atopy, asthma, allergies, atopic dermatitis ^5^; attention deficit hyperactivity disorder (ADHD), autistic spectrum disorder (ASD) ^6^; autoimmune diseases ^7^ such as type 1 diabetes ^8^ as possible long term outcomes of children born by CD. However, many of the studies were based on relatively small cohorts from a single geographic location, had a short follow-up period, did not account for possible confounders that might affect these associations and yield diverse results in regard to the magnitude of the effect ^1^.

While the optimal incidence of CD is debatable ^9–11^, a better understanding of how delivery mode affects long term health outcomes in children may influence health policies and decision processes of both clinicians and women. It could also serve as a basis for further research investigating the underlying biological mechanisms of these effects and possible interventions to improve pediatric outcomes of those born by CD. As conducting randomised controlled trials (RCTs) of mode of delivery is impractical and may be viewed as unethical, we utilized state-of-the-art causal inference methods on high quality, high volume, longitudinal observational data that originates from Israel’s largest healthcare provider, to assess long term adverse outcomes of children born by CD.

## Methods

Following an appropriate study design and methodologies tailored to the use of observational data for causal analysis has shown promise to overcome some of the issues that rise when working with such data ^12^. One such study design is the *Target Trial* ^*13*^ framework. Effect estimates from observational data obtained using this framework were comparable to those from RCTs. In accordance with this framework, we envisioned an hypothetical RCT which could estimate the causal effect of CD on pediatric outcomes, and utilized observational data to emulate it as close as possible. Table 1 summarizes key components of the desired hypothetical RCT versus their corresponding definition in our observational data. *Time-zero* (which can be viewed as target trial initiation), the eligibility-determining date in which treatment strategies are assigned and follow-up starts, was defined as the time of birth.

**Table 1.** Specification and emulation of a target trial of mode of delivery childhood health outcomes.

| Protocol component | Target trial specification: hypothetical RCT | Target trial emulation: existing observational data |
| --- | --- | --- |
| Eligibility criteria | 1. Births between 1 January 2007 and 31 December 2018<br>2. Only women who were in Clalit HMO for at least 5 years prior to birth<br>3. Exclude women who have more than one previous CD<br>4. Exclude preterm births, defined as delivery prior to 37 completed gestational weeks<br>5. Exclude children born small for gestational age (SGA), defined as birth weight less than 2500 gr<br>6. Exclude multiple gestations | 1. Same as for the target trial<br>2. We identified women with at least 5 years of documented medical history in Clalit's registry data.<br>3. We excluded women with more than one previous CD in their medical history data<br>4. We define preterm births by premature_flag available from hospital data, or gestational age at birth below 37 weeks<br>5. We exclude children with birth weight less than 2500 grams<br>6. We exclude multiple gestations<br>7. We require information on mother-child linkages, delivery date, mode of delivery and neonatal sex |
| Treatment strategies | vaginal or cesarean deliveries | Same as for the target trial |
| Treatment assignment | Mothers are randomly assigned to a delivery strategy (cesarean section or vaginal delivery) at birth and will be aware of the strategy to which they have been assigned. | Mothers were classified by a delivery strategy (cesarean section or vaginal delivery) according to their birth data. Randomization was emulated by adjusting for baseline confounders. |
| Outcomes | ➤ Asthma<br>➤ Overweight \ Obesity (defined by BMI z-score)<br>➤ Atopic dermatitis<br>➤ Celiac disease<br>➤ Allergies<br>➤ Autistic spectrum disorder (ASD)<br>➤ Attention deficit hyperactivity disorder (ADHD)<br>➤ Inflammatory bowel disease (IBD)<br>➤ Type 1 diabetes<br>➤ Juvenile idiopathic arthritis<br>➤ Respiratory infections<br>➤ Autoimmune diseases<br>➤ Atopy<br>➤ Death | Same as for the target trial |
| Follow-up | Follow-up starts at treatment assignment (birth time) and ends at loss to follow-up, 10 years after baseline, or on 31 December 2018, whichever occurs first. | Same as for the target trial |
| Causal contrasts | Intention-to-treat effect | Observational analog of intention-to-treat effect |
| Statistical analysis | Intention-to-treat effect is estimated by fitting a pooled logistic regression model | Same as for the target trial - also adding to the pooled logistic model the baseline covariates |

### Data

Data of the main cohort - Israel cohort, were extracted from the Clalit Health Services (Clalit) database, which is the largest Health maintenance organization (HMO) in Israel ^14^. Clalit is a nongovernmental, nonprofit organization with an electronic health record (EHR) database of more than 5 million patients, representing over 50% of Israel’s adult population (Section 1a in the Appendix). The data includes anthropometrics measurements (height and weight), blood pressure (BP) measurements, blood and urine laboratory test results, diagnoses recorded by physicians, dispensed pharmaceuticals and family linkage.

### Replication data - UK cohort

Data of the replication cohort - UK cohort, were extracted using primary care electronic health records from IQVIA Medical Research Data (IMRD), incorporating data from The Health Improvement Network (THIN, a Cegedim database). This database contains records of more than 12.5 million patients, covering approximately 6% of the UK population, and is representative of the population in terms of demographics and condition prevalence ^15^. The data includes patient demographics, medical diagnoses, medication prescriptions, anthropometrics measurements and laboratory test results, which were transformed to the OMOP common data model ^16^.

### Study population

We analyzed a total of 737,904 births, from 2002 until 2018. Birth rates in general, and specifically CD rates in Israel were relatively stable, but with a small linear decreasing trend, during this time period, with roughly 17% of births delivered by CD (Fig. S2.1).

Eligible birth records were required to contain information which we defined as critical for our analysis, including mother-child linkage and at least 5 years of documented maternal medical history in Clalit’s EHR data prior to delivery. Additional exclusion criteria were preterm birth (delivery prior to 37 completed gestational weeks) and low birth weight (birth weight below 2500 grams). While clinicians, supported by guidelines ^17^, generally recommend or offer a trial of vaginal delivery after one CD, it is generally not offered after two or more CDs. Therefore, women with a history of 2 or more CDs were also excluded from the study. Figure 1 describes eligibility criteria and flow chart of cohort selection, and Table 2 summarizes baseline characteristics of the 238,159 eligible children.

**Table 2.** Baseline characteristics of the study population - Israeli cohort.

|  |  |  |  |
| --- | --- | --- | --- |
| **Characteristic, mean (SD) or counts %** | **Vaginal (n = 200,879) (84.3%)** | **Cesarean (n = 37,280) (15.7%)** | **All (n = 238,159)** |
| **Neonatal characteristics** |  |  |  |
| Sex - (Male) | 102,717 (51.13%) | 20,264 (54.36%) | 122,981 (51.64%) |
| Birth weight (grams) | 3,291 (391) | 3,306 (460) | 3,293 (403) |
| Gestational age at delivery (weeks) | 39.32 (1.14) | 38.74 (1.24) | 39.23 (1.18) |
| **Maternal characteristics** |  |  |  |
| Maternal age (years) | 29.78 (5.34) | 32.11 (5.44) | 30.14 (5.42) |
| Maternal weight pre-pregnancy (Kg) | 62.37 (12.78) | 66.92 (15.51) | 63.12 (13.38) |
| Maternal height (meters) | 1.62 (0.06) | 1.61 (0.07) | 1.62 (0.06) |
| Diabetes Mellitus | 2,461 (1.23%) | 941 (2.52%) | 3,402 (1.43%) |
| Chronic Hypertension | 1,642 (0.82%) | 814 (2.18%) | 2,456 (1.03%) |
| Gravidity | 0.77 (0.81) | 0.53 (0.65) | 0.73 (0.79) |
| Previous cesarean deliveries | 6,604 (3.29%) | 11,514 (30.89%) | 18,118 (7.61%) |
| Sister's cesarean deliveries, percent* | 12.25 (27.98) | 18.7 (34.38) | 13.06 (28.95) |
| **Pregnancy characteristics** |  |  |  |
| Gestational Diabetes | 9,439 (4.7%) | 3,588 (9.62%) | 13,027 (5.47%) |
| Gestational Hypertension | 1,867 (0.93%) | 607 (1.63%) | 2,474 (1.04%) |
| Gestational weight gain z-score | -0.51 (2.42) | -0.31 (2.56) | -0.47 (2.45) |

Gestational weight gain z-score was calculated according to Hutcheon et al. 2013.
\*Sister's CD percent was calculated as the percentage of CDs over all sister's pregnancies which ended prior to the relevant participant's date of birth.

**Figure 1:**
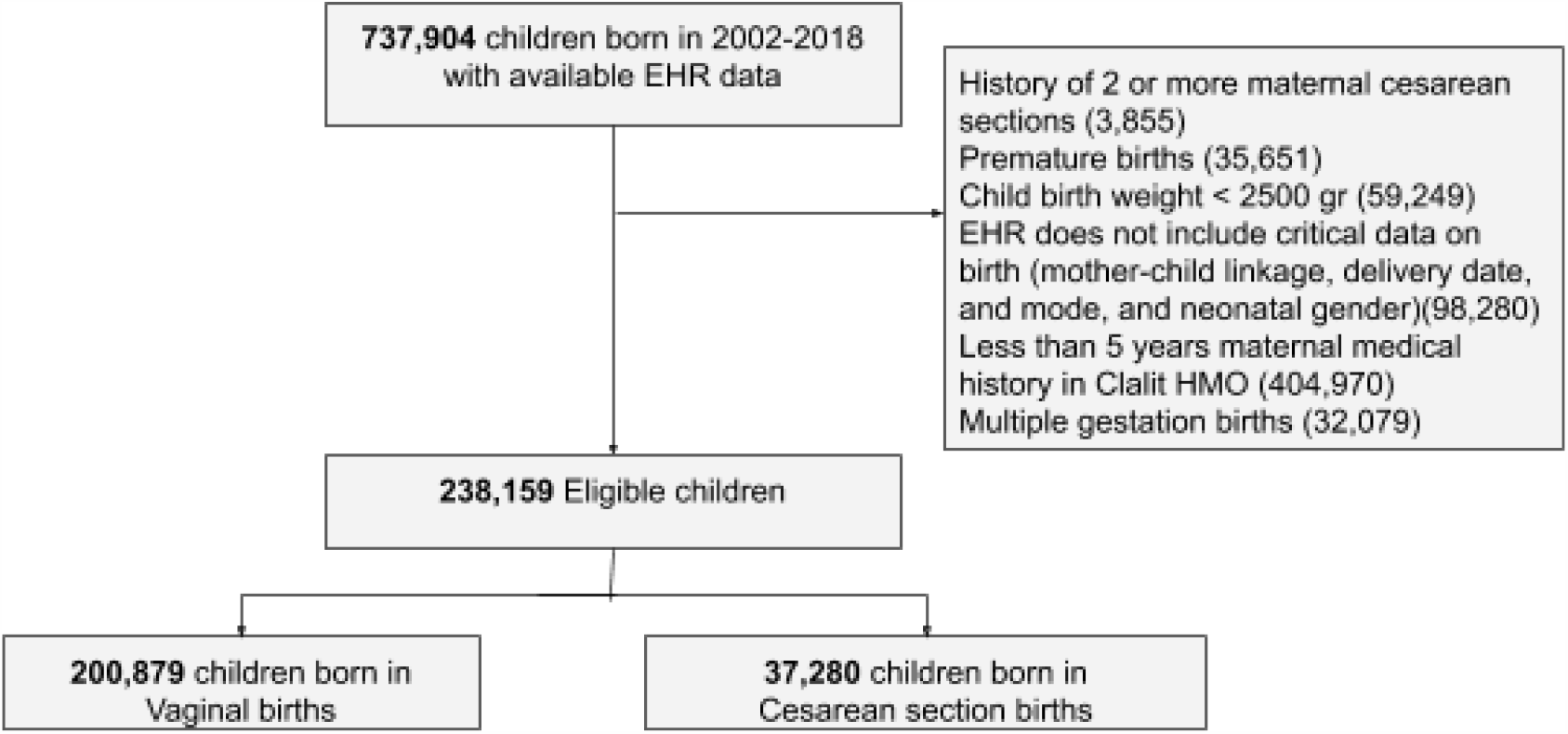
Cohort selection process. Flowchart for selection of eligible individuals from the Clalit database for emulating a target trial of birth delivery mode and childhood health outcomes. Numbers in parentheses represent unique individuals in each group. Abbreviations: EHR- Electronic health records, HMO- Health maintenance organization

625,044 births, from 1994 until 2019 were analyzed in the UK cohort. Of these, 250,269 births could be linked to the child’s medical record. Table S2.1 summarizes baseline characteristics of the 163,272 eligible children from this cohort.

### Statistical analysis

Directed Acyclic Graphs (DAGs) ^18,19^, were constructed together with domain expert physicians both in obstetrics and pediatrics (Fig. S8.1). We trained a propensity model ^20^ estimating the probability of being treated, i.e. giving birth by CD, using variables selected using the DAG described above. For learning the propensity model with a large number of covariates and to allow nonlinearities, we trained Gradient Boosting trees ^21^ (see Section 5 in the Appendix for detailed information on the model). Evaluation of the propensity model and covariate balance was done in a similar manner to the workflow described by Shimoni, Y. *et al*. ^22^. In Fig. 2A the distributions of propensities for each treatment group are plotted, and overlap between delivery modes is observed at least up to a score of 0.4. Covariate balance before and after reweighting is presented in Fig. 2B (see Section 5 in the Appendix for a detailed analysis).

**Figure 2:**
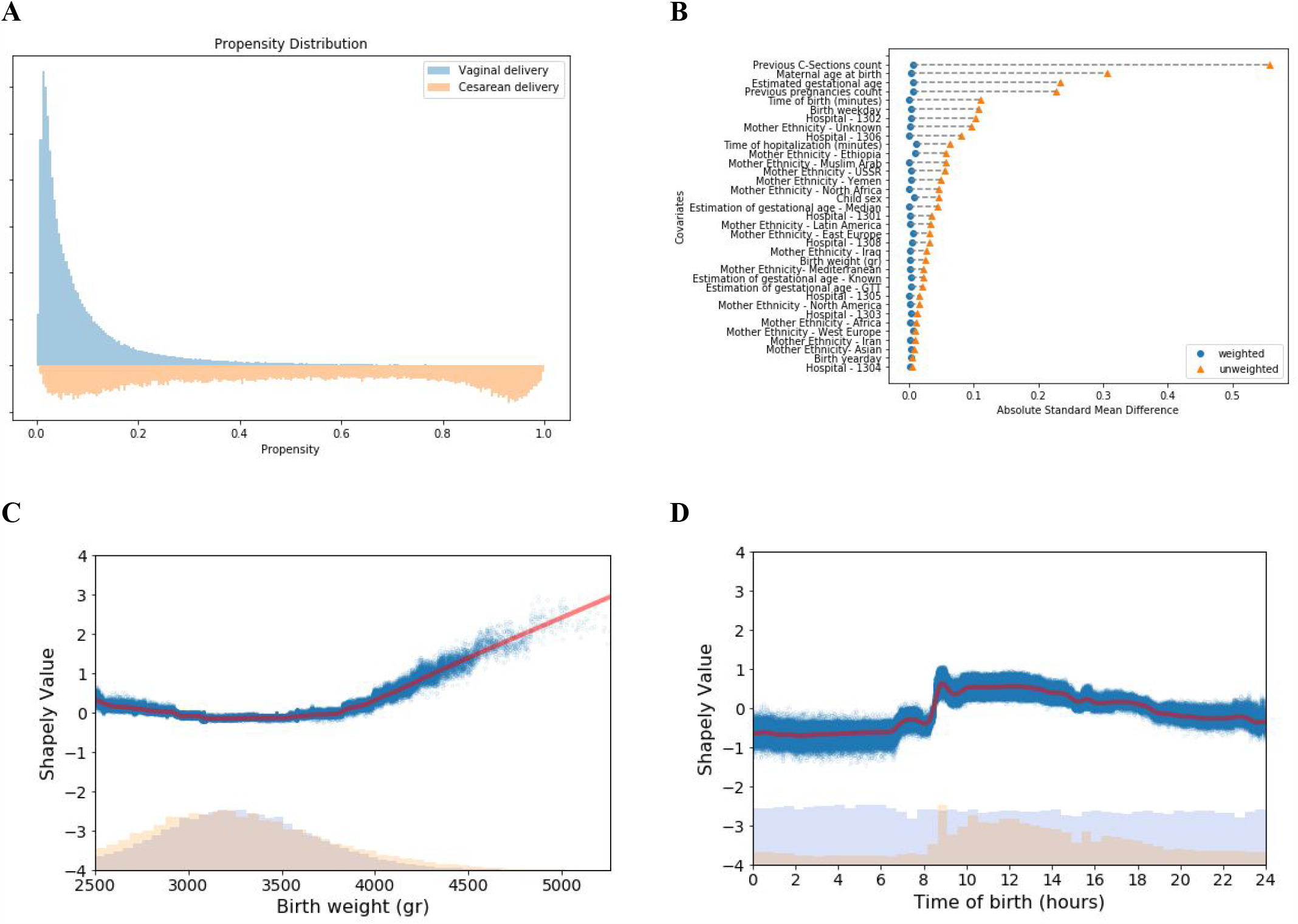
Propensity model evaluation for cesarean delivery. **A:** Distribution of propensity predictions; Blue-Vaginal delivery, Orange-CD. **B:** Absolute standardized mean differences of baseline covariates between birth mode groups. Orange triangles are the differences before weighing; Blue circles - differences after applying weighting. **C, D**: Dependence plots of birth weight and time of birth; lower part shows a histogram of the distribution of number of births, and the upper part shows a dependence plot of the log-odds Shapley value for a CD. Blue dots represent individual covariate values and Shapley values, red line represents a locally weighted scatterplot smoothing (LOWESS) regression curve

Applying a feature attribution framework for machine learning models based on estimated *Shapley values* ^23^, we were able to estimate the contribution of the baseline covariates to the estimated propensity. This setup allowed us to capture any non-linear relationships between a covariate’s contribution and the prediction value. For example, we observed a non-linear *U-shaped* impact of birth weight on the propensity score (as demonstrated for the upper range of birth weights > 2500 grams in Fig. 2C). This relation is well-known ^24^ and was predicted to exist by Obstetricians who were asked to investigate our model. Another interesting variable that had a non-linear impact on the propensity model was the time of day at delivery. It is known that elective CDs are scheduled for daytime working hours (8am-4pm), while vaginal deliveries are expected to distribute uniformly throughout the entire day (see Fig. S3.3 and Fig. 2D). Utilizing the additive property of Shapley values, we were also able to analyze groups of related features according to domain knowledge (Fig. S5.3).

### Outcomes

Long term childhood outcomes of CD were first identified by previous studies that demonstrated associations between these outcomes and the mode of delivery ^1,7^. Each outcome was then defined and ascertained separately by a trained pediatrician. The diagnoses were based on the relevant ICD-9 codes, laboratory test results, and prescriptions of medications for the relevant medical conditions. When feasible, the diagnoses were based on previously published diagnostic approaches from EHR available in the literature or https://phekb.org/phenotypeswebsite, and in accordance with the Israeli healthcare policies and definitions (see section 4 in the Appendix for a detailed description). Only outcomes with at least 10 diagnosed children who were born by each delivery mode were included in the main analysis, and the rest are presented in section 6 of the Supplementary Appendix.

### Estimation of delivery mode effect

For each of the described outcomes, we estimate the causal effect with 3 different strategies: (1) Weighting by inverse-probability-weighting (IPW) ^25^, (2) Weighting by overlap-weights (OW) ^26^, and (3) Standardization ^27^. The different outcomes can be categorized to 2 different types: (I) time-to-event outcomes (such as asthma onset), for which survival curves were estimated; and (II) fixed continuous outcomes (such as BMI z-score measured at the age of 5 years), for which average treatment effect (ATE) differences were estimated. For time-to-event outcomes (such as asthma), survival curves were constructed by fitting a pooled logistic regression model ^28,29^, on a person-time data format. To adjust for baseline confounders we either added baseline selected variables to the logistic model (standardization), or performed weighting using either IPW or OW. Both weighting methods were evaluated when constructing the propensity model, OW resulted in a weighted population with better covariance balance (Fig. S5.5).

### Sensitivity analysis

As some of our assumptions cannot be verified from the data, we performed several sensitivity analysis as described below (full results are derailed in Section 6 of the Supplementary appendix.

### Negative controls

One tool to detect unmeasured confounding is the use of negative controls ^30^. Here, upper forearm fracture (upper end of radius and ulna), a relatively common diagnosis in children, was chosen as negative control since no studies thus far indicated that there is a plausible association between this diagnosis and CD.

### Elective CD subpopulation

In the UK cohort, data on whether CD was unscheduled or elective was available, allowing us to create an elective CD subpopulation, which includes 141,222 children. Baseline characteristics of this subpuliation can be found in the supplementary Appendix (Table S6.6). In the Israel data, a clear distinction between unscheduled and elective CD was not available. To estimate the probability of elective CD in this cohort, we utilized an additional data set, in which the type of CD was specified, from in-hospital electronic records, obtained from Rabin Medical Center, the third largest medical center in Israel. The data set contained information on 56,260 births between 2012-2020, of which 3,827 were by unscheduled CD and 3,972 by elective CD. Using this additional data, we built a model that predicts elective CD from variables that are also present in the original Clalit EHR database (see section 7 in the supplementary Appendix). We used this trained model to obtain predictions for elective CD in our cohort, and created a subpopulation which contained vaginal deliveries from our full study population and CDs which were predicted as elective with a high probability (greater or equal to 0.5) (Fig. S7.2). Baseline characteristics of this subpopulation of 217,258 children can be found in the supplementary Appendix (Table S6.5).

### Clinics matched subpopulation

In this subpopulation, children were grouped by maternal clinics (defined as the most frequently visited clinic prior to pregnancy) and year of birth. In each group we sampled an equal number of CDs and vaginal deliveries. Baseline characteristics of this subpopulation of 66,464 children can be found in the Appendix (Table S6.7).

### Siblings matched subpopulation

To create a subgroup of siblings we made use of the longitudinal and large scale family data available in the Clalit EHRs and built a sensitivity analysis based on a subpopulation of *discordant siblings* comprised of pairs of siblings born in discordant birth modes, where both siblings are with the same sex, and matching the birth order by the number of first CD and first vaginal pairs. Baseline characteristics of the 3,936 children in the *discordant siblings* subpopulation can be found in the supplementary Appendix (Table S6.8).

## Results

Overall, 238,159 eligible births, 84.35% vaginal and 15.65% cesarean deliveries, were included in the Israel cohort (Table 2). In the replication UK cohort, 163,272 eligible births, 77.01% vaginal and 22.99% cesarean deliveries were included (Table S2.1). Children had a mean follow-up of 5.6b6 (SD 2.96) years and 6.77 (SD 4.54) years, in the Israel and UK cohorts, respectively. In both cohorts, the largest 10-year standardized risk differences were found in the following outcomes: atopy, asthma and allergy. For these outcomes, the 10-year standardized risk differences observed were 0.74% (−0.06, 1.52) for atopy, 0.64% (0.31, 0.98) for asthma and 0.47% (−0.32, 1.28) for allergy in the Israel cohort and 0.41% (−0.42, 1.15), 1.04% (0.49, 1.71), and 0.23% (0.07, 0.36) in the UK cohort respectively (Table 3, Fig. 3, 4).

**Table 3.** Estimated 10-yr childhood disease-free risk differences for comparing vaginal and cesarean deliveries.

|  |  |  |  |  |  |  |
| --- | --- | --- | --- | --- | --- | --- |
| Full study population |  |  |  |  |  |  |
| Childhood health outcome | Israel cohort |  |  | UK cohort |  |  |
|  | Number of incident cases |  | 10-yr risk difference (95% CI) | Number of incident cases |  | 10-yr risk difference (%) |
|  | Vaginal (n=200,879) | Cesarean (n=37,280) |  | Vaginal (n=125,743) | Cesarean (n=37,529) |  |
| Atopy | 55,505 (27.63%) | 11,483 (30.8%) | 0.74% (-0.06, 1.52) | 35,805 (28.47%) | 10,747 (28.64%) | 0.41% (-0.42, 1.15) |
| Asthma | 11,900 (5.92%) | 2,756 (7.39%) | 0.64% (0.31, 0.98) | 9,117 (7.25%) | 2,789 (7.43%) | 1.04% (0.49, 1.71) |
| Allergy | 34,045 (16.95%) | 6,924 (18.57%) | 0.47% (-0.32, 1.28) | 1,270 (1.01%) | 467 (1.24%) | 0.23% (0.07, 0.36) |
| ADHD | 6,119 (3.05%) | 1,485 (3.98%) | 0.34% (-0.16, 0.82) | 516 (0.41%) | 114 (0.3%) | -0.17% (-0.33, 0.03) |
| Atopic dermatitis | 20,019 (9.97%) | 4,201 (11.27%) | 0.26% (-0.23, 0.7) | 30,234 (24.04%) | 9,031 (24.06%) | -0.1% (-0.87, 0.63) |
| Autistic spectrum disorder | 1,212 (0.6%) | 400 (1.07%) | 0.24% (0.02, 0.46) | 1,887 (1.5%) | 592 (1.58%) | 0.03% (-0.27, 0.39) |
| Death | 583 (0.29%) | 169 (0.45%) | 0.13% (0.03, 0.24) | 209 (0.17%) | 73 (0.19%) | 0.01% (-0.97, 0.03) |
| Type 1 diabetes mellitus | 79 (0.04%) | 20 (0.05%) | -0.01% (-0.05, 0.03) | 33 (0.03%) | 14 (0.04%) | 0.16% (-0.01, 0.09) |
| Negative control (arm fracture) | 3,581 (1.78%) | 707 (1.9%) | -0.05% (-0.42, 0.35) | 3,278 (2.61%) | 951 (2.53%) | 0.26% (-0.09, 0.74) |
| Celiac | 784 (0.39%) | 123 (0.33%) | -0.12% (-0.26, 0.05) | 223 (0.18%) | 81 (0.22%) | 0.0% (-0.09, 0.09) |
| Autoimmune diseases | 1,062 (0.53%) | 178 (0.48%) | -0.13% (-0.29, 0.04) | 597 (0.47%) | 200 (0.53%) | 0.12% (-0.04, 0.3) |
| Elective CD subpopulation |  |  |  |  |  |  |
| Childhood health outcome | Israel cohort |  |  | UK cohort |  |  |
|  | Number of incident cases |  | 10-yr risk difference (95% CI) | Number of incident cases |  | 10-yr risk difference (%) |
|  | Vaginal (n=200,879) | Cesarean (n=16,379) |  | Vaginal (n=125,743) | Cesarean (n=15,479) |  |
| Atopy | 55,505 (27.63%) | 4,861 (29.68%) | -0.29% (-1.53, 0.9) | 35,805 (28.47%) | 4,383 (28.32%) | 0.02% (-1.02, 1.77) |
| Asthma | (5.92%) 11,900 | 1,194 (7.29%) | 0.37% (-0.09, 0.92) | 9,117 (7.25%) | 1,151 (7.44%) | 0.96% (0.05, 2.33) |
| Allergy | 34,045 (16.95%) | 2,898 (17.69%) | -0.13% (-1.51, 1.28) | 1,270 (1.01%) | 196 (1.27%) | 0.24% (0.02, 0.55) |
| ADHD | (3.05%) 6,119 | 585 (3.57%) | 0.2% (-0.6, 0.92) | 516 (0.41%) | 53 (0.34%) | -0.05% (-0.34, 0.26) |
| Atopic dermatitis | (9.97%) 20,019 | 1,746 (10.66%) | 0.33% (-0.27, 0.95) | 30,234 (24.04%) | 3,668 (23.7%) | -0.32% (-1.39, 1.32) |
| Autistic spectrum disorder | (0.6%) 1,212 | 178 (1.09%) | 0.19% (-0.05, 0.47) | 1,887 (1.5%) | 249 (1.61%) | -0.09% (-0.58, 0.61) |
| Death | 583 (0.29%) | 66 (0.4%) | 0.07% (-0.04, 0.22) | 209 (0.17%) | 23 (0.15%) | -0.03% (-0.1, 0.0) |
| Type 1 diabetes mellitus | 79 (0.04%) | 8 (0.05%) | 0.0% (-0.1, 0.0) | 33 (0.03%) | 9 (0.06%) | 0.08% (0.0, 0.17) |
| Negative control (arm fracture) | 3,581 (1.78%) | 306 (1.87%) | -0.14% (-0.72, 0.38) | 3,278 (2.61%) | 418 (2.7%) | 0.42% (-0.25, 1.43) |
| Celiac | 784 (0.39%) | 52 (0.32%) | -0.26% (-0.42, -0.05) | 223 (0.18%) | 43 (0.28%) | 0.8% (-0.06, 0.23) |
| Autoimmune diseases | 1,062 (0.53%) | 78 (0.48%) | -0.23% (-0.4, -0.04) | 597 (0.47%) | 100 (0.65%) | 0.28% (0.07, 0.59) |

ADHD - Attention deficit hyperactivity Disorder

**Figure 3:**
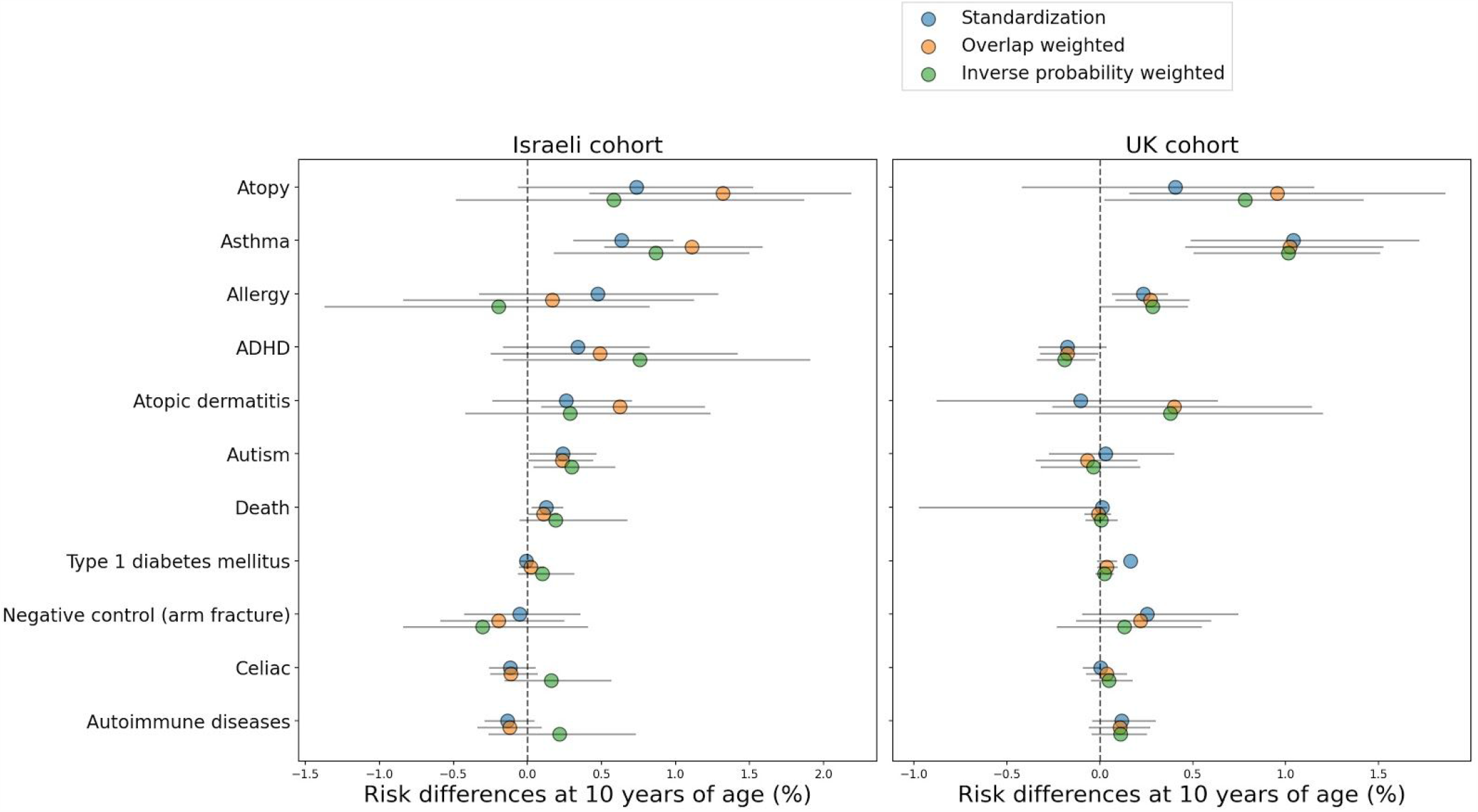
Estimated 10-yr risk difference (%) between children born vaginally or through CD. Results are shown in the Israeli cohort (Left) and UK cohort (Right), when using Standardization (blue circles), weighting with Overlap weights (orange circles) and weighting with Inverse-Probability-Weighting (green circles), sorted by standardized risk differences in the Israel cohort. Black lines represent 95% confidence intervals.

**Figure 4:**
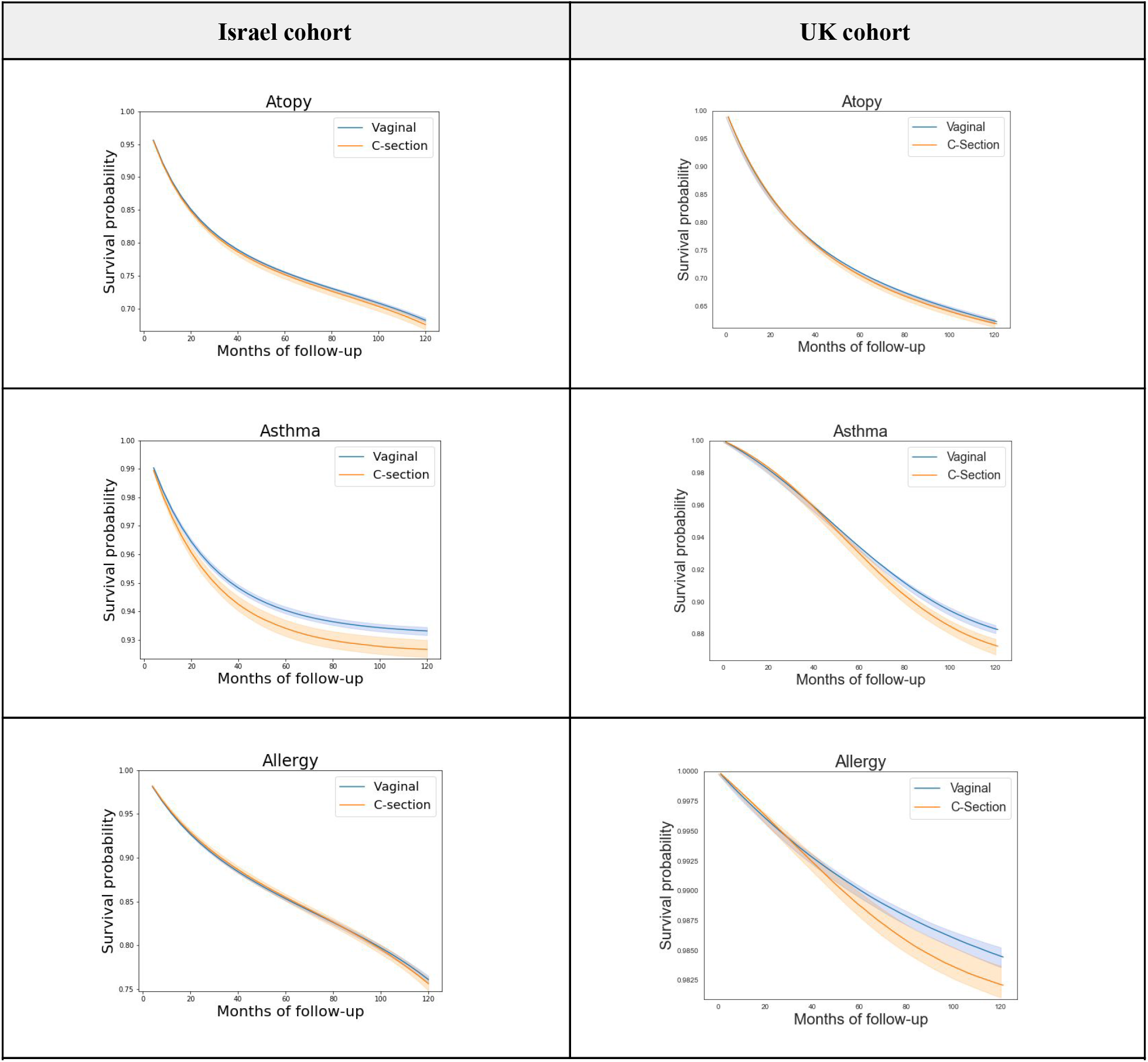
Childhood disease-free survival curves comparing vaginal and cesarean deliveries for top 3 childhood outcomes. Standardized survival curves with 95% CI of children born through vaginal delivery (blue) and CD (orange) are shown by months of follow-up. **Left:** Israel cohort, full study population. **Right:** UK cohort, full study population

A small risk difference was found in the Israel cohort for ADHD, 0.34% (−0.16, 0.82) and for ASD, 0.24% (0.02, 0.46), however, these differences were not found in the UK cohort. An average effect of 0.10 (0.07, 0.12) and 0.92 (0.68, 1.14) was found for BMI z-score at age 5-6 years old and for the number of respiratory infection incidents until 5 years of age in the Israel cohort accordingly (Table 4). Of note, in all of these outcomes, a similar trend was observed in all estimation strategies: IPW, OW, and standardization, with the exception of IPW on the allergy outcome in the Israeli cohort (Tables 3,4). Additional predefined long term pediatric outcomes, including death, type 1 diabetes mellitus, celiac, and autoimmune diseases had a negligible risk difference both in the Israel and UK cohort. Forearm fracture, which was used as a negative control (see Methods) also had a negligible risk difference, further validating our findings. As additional stringent analyses, we next studied the effect of CD in specific subpopulations as follows:

**Table 4.** Estimated childhood obesity, antibiotics intake and clinic visits differences for comparing vaginal and cesarean deliveries

| **Full study population** |  |  |  |  |  |  |
| --- | --- | --- | --- | --- | --- | --- |
| Childhood health outcome | Israel cohort |  |  | UK cohort |  |  |
|  | Number of children with sufficient data |  | ATE difference | Number of children with sufficient data |  | ATE difference |
|  | Vaginal | Cesarean |  | Vaginal | Cesarean |  |
| BMI z-score at age 5-6 years old | 99,345 (83.6%) | 19,454 (16.4%) | 0.10 (0.07, 0.12) | - | - | - |
| Respiratory infections - number of incidents until 5 years of age | 107,654 (83.8%) | 20,864 (16.2%) | 0.92 (0.68, 1.14) | 70,705 (78.24%) | 19,666 (21.76%) | 0.12 (0.08, 0.15) |
| **Elective CD subpopulation** |  |  |  |  |  |  |
| BMI z-score at age 5-6 years old | 99,345 (92.5%) | 8,058 (7.5%) | 0.10 (0.08, 0.12) | - | - | - |
| Respiratory infections - number of incidents until 5 years of age | 107,654 (92.6%) | 8,577 (7.4%) | 1.11 (0.91, 1.35) | 70,705 (89.63%) | 8,181 (10.37%) | 0.10 (0.06, 0.15) |

BMI z-score calculated according to 2000 CDC Growth Charts for the United States <sup>36</sup>, ATE - Average Treatment Effect

### Elective CD subpopulation

While some studies have shown a similar association between elective and unscheduled CD and pediatric outcomes ^31,32^, others have shown different correlations depending on the type of CD performed ^33,34^. possibly due to the fact that unscheduled CDs could result from an emergency during labor, and therefore additional factors might affect the newborn’s health. We therefore created a subgroup of children born by elective CD for both cohorts. However, while in the UK cohort, information on the type of CD was available, in the Israeli cohort it was not, and was therefore estimated by other parameters (see Methods). Overall, 217,258 children from the Israel cohort and 141,222 children from the UK cohort were included in these analyses (Tables S6.5, S6.6). In both subgroups, standardized risk differences were still observed for asthma 0.37% (−0.09, 0.92) in the Israel cohort and 0.96% (0.05, 2.33) in the UK cohort respectively. However, the results for the other outcomes were not consistent in both subgroups (Table 3).

### Clinics matched subpopulation

Potential confounding factors such as environment, socioeconomic status and standard of care may have a substantial impact on the validity of the results. To investigate the sensitivity of our estimates to these factors, we analyzed a subpopulation that included 66,464 children born by either CD or vaginal deliveries grouped by maternal clinics and year of birth (see Methods, Table S6.7). In this subgroup, similarly to the full Israel cohort, a small risk difference was observed for ASD 0.44% (0.08, 0.88). Standardized risk differences for atopy, asthma and allergy were 0.93% (−0.36, 2.02), 0.31% (−0.14, 0.82) and 0.23% (−1.15, 1.47) respectively. Some of these risks are smaller than those seen in the full Israel population, and they were not consistent across all analysis methods (Table S6.2, Fig. S6.1), possibly due to the relatively small sample size of this group.

### Siblings matched subpopulation

Finally, as it has been priviously shown that siblings studies might deal well with potential unobserved confounders related to environmental and genetic factors ^35^ we analyzed a subpopulation of 3,936 children that included pairs of same-sex siblings born in discordant birth modes (see Methods, Table S6.8). In this subgroup we observed risk differences of 0.4% (−4.43, 5.92) for atopy, 1.7% (−0.37, 3.84) for asthma and 1.85% (−4.62, 8.32) for allergy, which were also consistent in all analysis methods, apart from IPW on asthma (Table S6.2, Fig. S6.1). Despite the large CIs, which might stem from the very small size of this subpopulation, the same trend is observed for these outcomes in this subpopulation.

Standardized survival curves for all outcomes, in the Israel and UK cohorts can be seen in Fig. S6.2. Unadjusted Kaplan-Meier curves for all outcomes and full results that include OW and IPW, as well as all sensitivity analyses can be found in Section 4 and 6 of the Supplementary appendix.

## Discussion

In this study, we utilized data from the largest HMO in Israel in order to assess long term pediatric adverse outcomes of CD, compared to vaginal delivery. We analysed the effect of delivery mode on several predefined outcomes among 238,159 children for a mean follow-up period of 5.66 (SD 2.96) years in Israel, and 163,272 children with a mean follow-up of 6.77 (SD 4.54) years in the UK. We revealed that CD had an effect on the occurence of asthma, atopy, allergy and the number of respiratory infection events by the age of 5 years old both in the Israel and UK cohorts. An effect of CD on BMI z-scores at the age of 5 years old was also observed in the Israel cohort, but this result could not be validated in the UK cohort since routine anthropometric measurements for children were not available. Although results were not replicated for all outcomes in the different subgroups analysed, it may be a result of insufficient power to detect these differences in these small scale cohorts, as similar trends were often observed.

The effect of mode of delivery on pediatric outcomes is hypothesized to be mediated by several mechanisms ^37^. These include: hormonal surges and exposure to different levels of physical stress during labor, which may trigger protective developmental processes in the newborns and play a role in normal postnatal physiological development ^38^; perturbations in the transmission of maternal microbiome to the infant during CD, which may result in different establishment and diversity of the microbiota thus possibly affecting future childhood health ^39,40^; changes in the regulation of gene expression as a result of alterations in epigenetic patterns such as DNA-methylation ^41^; abnormal short-term immune responses observed in infants born in CD, such as reduced expression of inflammatory markers ^7,42^; and exposure to general anesthesia and anesthetic medications that may cross the placental barrier ^43^.

While evidence regarding the effect of birth mode on future long term outcomes of children is accumulating ^1,44^, conducting an RCT to assess these effects is unfeasible and may be considered unethical. For example, in a recently published meta-analysis that included 80 studies on the effects of CD, only one RCT on the effects of CD was identified ^44^. However, observational studies may be restricted by failure to control for confounders, study design limitations and inappropriate selection of groups for comparison. To overcome some of these challenges, we adopted the *target trial* framework which relies on counterfactual reasoning which facilitates a systematic methodology for evaluation of observational studies and allows transportability of our estimates ^13^. We further analysed if the effect observed is sustained in a replication cohort and in several stringent sensitivity analysis, including changes to the inclusion criteria, adjustment for other potential confounders and changes to model assumptions.

Among all pediatric outcomes analysed, CD had the largest effect on atopy and asthma development (0.74% and 0.64% accordingly in Israel cohort, 0.41% and 1.04% accordingly in UK cohort). Risk differences for asthma were also observed in several sensitivity analysis subpopulations; 0.37% (−0.09, 0.92) in the estimated elective CD subpopulation, 0.96% (0.05, 2.33) in the UK elective CD subpopulation, 0.31% (−0.14, 0.82) in the clinics matched subpopulation and 1.7% (−0.37, 3.84) in the siblings matched subpopulation. Notably, although the magnitude of these risk differences are relatively small, the relative risk of asthma diagnosis by 10 years of age increased by almost 10% in children born by CD in the Israeli cohort and by 8.91% in the UK cohort. The association between CD and the development of asthma was previously demonstrated in numerous studies. A meta-analysis found an overall increase of 20% in the risk of future asthma in children who were delivered by CD ^45,46^. A possible mechanism for this association was recently demonstrated in a study on the gut microbiome of children born by cesarean delivery showing an increased asthma risk only in children in whom microbiome composition at 1 year of age still retained a CD microbial signature, suggesting a role of altered maturation of the gut microbiota in the increased risk observed ^40^. Another hypothesis underlying this association is an altered pulmonary physiology as a result of a delayed removal of amniotic fluid from the lung of infants that are born by CD, resulting in transient tachypnea of the newborn and higher incidence of respiratory distress syndrome after birth ^47^, which may increase the risk for asthma in the future.

Interestingly, we found an average difference of 0.92 and 0.12 for the number of incidents of respiratory infections until the age of 5 years old, in Israel and the UK accordingly. The association between CD and number of respiratory infections was previously demonstrated ^48^, and may partially mediate the effect observed on asthma, as viral infections are important causes of wheezing illnesses in children of all age ranges ^49^ and evidence demonstrating a link between early viral infections and asthma inception and exacerbations is accumulating ^50^.

The effect of CD on the incidence of atopy in children in our study is mostly driven by its effects on the development of asthma and allergy, each affected by CD when analysed separately. In the Israel cohort, the effect on atopy is larger partially due to the fact that in this cohort, atopic dermatitis is also positively affected by CD in contrast to the UK cohort (Table 3). As expected, there is an overlap between these diagnoses (as presented for the Israel cohort in Fig. S4.11). Several previous studies did not find an association between CD and atopy ^5,44^ or atopic dermatitis ^51,52^. However, for some of these studies the follow-up time was relatively short (1-3 years) ^51,52^. Meta-analyses analysing this effect display significant heterogeneity between studies ^5,44^.

A smaller risk difference was found for ADHD, 0.34% (−0.16, 0.82), and ASD, 0.24% (0.02, 0.46), between infants born by CD and those born by vaginal delivery in the Israeli cohort. These differences were smaller in the estimated elective CD subpopulation - 0.2% (−0.6, 0.92) and 0.19% (−0.05, 0.47), and even smaller in the UK cohort - −0.17% (−0.33, 0.03) and 0.03% (−0.27, 0.39), for ADHD and ASD respectively. Other subpopulations and methods used as sensitivity analyses resulted in wide estimates which included zero for both outcomes. Previous studies resulted in opposite conclusions regarding the effect of CD on the development of ADHD and ASD. While some studies found that children born by CS are roughly 20% more likely to be diagnosed with ASD ^6^, others concluded no association exists ^53^. A previous study highlighted a possible association between general anesthesia during CD and ASD ^54^. Lack of information on social parameters and variable diagnosis procedures of ADHD and ASD in different health systems might explain the variability in our results. For example, it was previously shown that a diagnosis of ASD spectrum disorder at a younger age is associated with geographic proximity to a higher number of medical specialists diagnosing ASD (neurologists and psychiatrists) ^55^. Further research with more comprehensive environmental information is therefore needed to better estimate the effects on these outcomes.

Finally, we have found CD had an average difference of 0.10 for BMI z-scores at age 5 years old. Very similar differences were found in all sensitivity analyses performed. The association of CD with subsequent obesity of the offspring was vastly studied, with mixed conclusions regarding the existence and magnitude of the effect. While some studies did not find any effect of delivery mode on childhood overweight ^56^, two meta-analyses of the literature concluded that CD is associated with an increased risk of subsequent obesity in offspring ^57,58^. It has been hypothesized that other than the above mentioned mechanisms, the difference may also arise from an altered level of appetite regulation hormones, as evident from a lower concentration of circulating ghrelin ^59^, and a lower umbilical leptin concentration ^60^ in infants born by CD.

Our study has several strengths. First, we analyzed a large and comprehensive nationwide dataset including both maternal and offspring’s data, with a long follow-up period, allowing as high as 10 years of follow-up for some of the children in the cohort. Second, we replicated our estimates using the same computational methods on an independent cohort which vary in both genetic and environmental factors, providing further validation to our findings. Finally, while the relation between CD and several long term childhood outcomes has been previously investigated in many observational studies, most have concentrated on associations rather than explicitly pursuing a causal estimate. We believe that our unique dataset, both in terms of the length of the follow-up period and in terms of the representation of a nationwide population, alongside state-of-the-art causal inference methods, allowed us to more accurately estimate the causal effect of CD on pediatric outcomes.

However, our study also has several limitations. Our dataset does not contain information on some potential confounding factors such as maternal nutritional and lifestyle habits, environmental exposures, parental education and detailed family socioeconomic information. For example, it has been previously postulated that a delay in breastfeeding could provide a confounding influence on the effect of CD on childhood outcomes, as delivery via CD is associated with lower rates of breastfeeding and early breastfeeding cessation ^61^. However, several studies found that adjustment for breastfeeding does not change the association between CD and asthma. In addition, our data did not include explicit information on whether the CD was a result of an emergency during labour versus an elective procedure, a differentiation that might be important. Although some studies found a similar association of the two CD types with pediatric outcomes ^31,32^, one study showed an association only between elective CD and risk of obesity at 12 months ^62^. To try to overcome this we utilized an additional in-hospital dataset, and analyzed a subpopulation of CDs which were classified as elective with a high probability. Although this does not replace a clear indication of whether the CD was elective, it further eliminates potential confounders which stem from emergencies during labor. Exploiting the fact that the UK replication cohort included distinction between emergency and elective CD, we were able to analyze the subpopulation of elective CD in this cohort. Our data also lack information on other potential confounding factors related to labor, such as information on mode of anesthesia and usage of anesthetic medications during labor or surgery. Finally, our results are only valid for term, appropriate for gestational age infants. Although the rate of CD delivery is potentially higher for these children, they introduce a large number of possible confounders and therefore were excluded from the analysis.

In conclusion, by emulating a *target trial* on two cohorts from different countries, we explicitly estimate and find a small causal effect of CD on several pediatric health outcomes, including asthma, atopy, allergy, respiratory infections and childhood BMI. For other outcomes, such as ASD and autoimmune diseases, an increased risk was not observed consistently when employing different methods and across both countries. Our findings might contribute to the ongoing discussion on the optimal rate of CD, with an emphasis on its adversarial effect on long term pediatric health and may enhance discussions between clinicians and parents regarding these risks. In addition, it may pave the way to future research on the mechanism underlying these effects and possible intervention strategies targeting them.

## Data Availability

The data that support the findings of this study originates from Clalit healthcare. Restrictions apply to the availability of these data and so are not publicly available.

## Code availability

Code for the causal survival analysis used to estimate the findings in this study can be found on Github: https://github.com/ayya-keshet/Causal-Survival-Analysis.

## Authors contribution

A.K, H.R, S.S, S.B.H conceived the project, designed and conducted the analyses, interpreted the results and wrote the manuscript and are listed in random order. G.A, M.B.B, C.Y and I.G conducted the analyses and wrote the manuscript. P.A, A.B.H and E.H interpreted the results. A.W and E.S conceived and directed the project and analyses, designed the analyses, interpreted the results, wrote the manuscript, and supervised the project.

## Ethics Declarations

The study protocol in this research was approved by the institutional review board (IRB) of the Rabin Medical Center, experiment protocol number 0158-19-RMC. Informed consent was waived by the IRB, as this is a retrospective study based on unidentified data taken from EHRs.

## Acknowledgements

We thank Gabi Barabash, Elad Barkan, Iris Kalka, Amir Gavrieli, Nitzan Artzi and members of the Segal group for fruitful discussions. We thank Uri Shalit for his inspirational course on Causal inference and useful comments.

## Supplementary Appendix

1. Data sources
2. Study cohort characteristics
3. Baseline covariates definitions
4. Pediatric outcomes definitions
5. Propensity model analysis
6. Expanded results and sensitivity analysis
7. DAGs for CD and pediatric health

## 1. Data sources

### a. Clalit EHRs - Israeli Cohort

Clalit is the largest of four health maintenance organizations (HMO) in Israel and one of the largest in the world. Serving over 4.4 million individuals, it includes over half of the Israeli population which is diverse in terms of race, ethnicity and socioeconomic status. Clalit owns and operates approximately 1,500 primary care clinics and 14 hospitals. Israel’s adoption of EHRs started during the mid-1990’s and provision of healthcare services have been recorded in EHRs for more than 20 years. Clalit’s member population is stable (< 2% annual turnover), the data is longitudinal (spans from birth to death), includes both claims and direct clinical data and is linked through a unique anonymized identifier. The data captures administrative and clinical information across hospitals (inpatient and emergency department settings), primary care clinics, specialty clinics, pharmacies, laboratories, and clinical measures. The data are unique in their breadth, historical depth and harmonization (single EMR software), and as they originate from an operating HMO - they are constantly being generated.

## 2. Study cohort characteristics

CD prevalence rate by year

**Figure S2.1:**
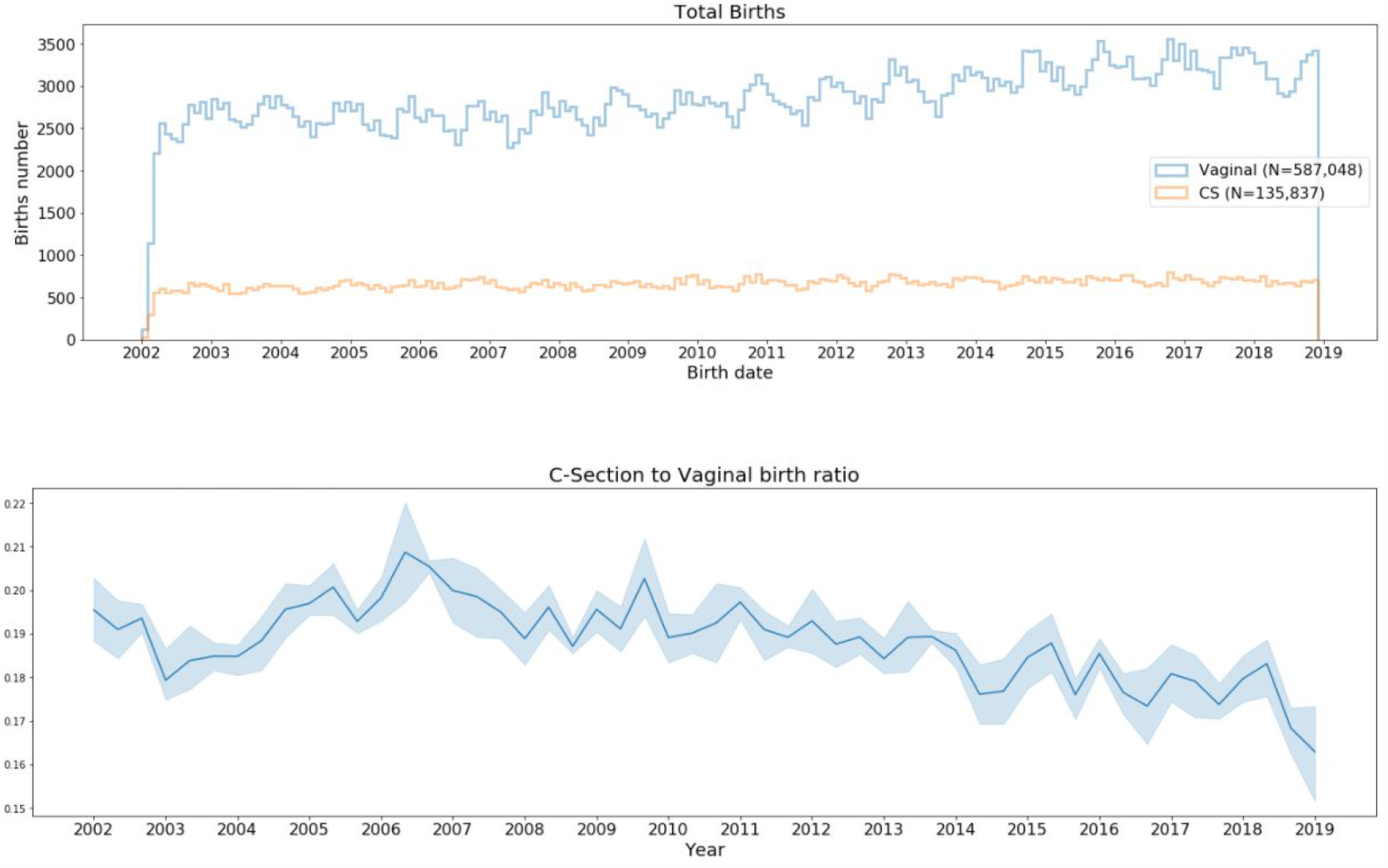
Birth rates by years - Israel cohort. **Top:** Birth rates in the years 2002 to 2019 in Clalit database; Blue-vaginal delivery, Orange-CD. **B:** CD to vaginal delivery births ratio in the years 2002 to 2019.

**Table S2.1.** Baseline characteristics of the study population - UK cohort.

| **Table S2.1. Baseline characteristics of the study population - UK cohort** |  |  |  |
| --- | --- | --- | --- |
| **Characteristic, mean (SD) or counts %** | **Vaginal (n = 125,743)**<br>**(77.01%)** | **Cesarean (n =37,529 )**<br>**22.99(%)** | **All (n = 163,272)** |
| **Infant characteristics** |  |  |  |
| Sex - Male | 63,755 (50.70%) | 19,875 (52.96%) | 83,630 (51.20%) |
| Birth weight (grams) (N=67,692) | 3,478 (453) | 3,510 (499) | 3,485 (464) |
| Gestational age at delivery (weeks) (N=158,264) | 40.24 (1.28) | 39.78 (1.41) | 40.14 (1.32) |
| **Maternal characteristics** |  |  |  |
| Maternal age (years) | 30.32 (5.59) | 32.07 (5.37) | 30.73 (5.56) |
| Maternal weight pre-pregnancy (Kg) (N=52,042) | 69.09 (16.36) | 72.87 (18.16) | 69.99 (16.88) |
| Maternal height (meters) (N=154,297) | 1.65 (0.07) | 1.63 (0.07) | 1.64 (0.07) |
| Diabetes Mellitus | 730 (0.58%) | 606 (1.61%) | 1336 (0.82%) |
| Chronic Hypertension | 528 (0.42%) | 287 (0.76%) | 815 (0.50%) |
| Gravidity | 0.82 (1.21) | 0.78 (1.16) | 0.81 (1.20) |
| Previous cesarean deliveries | 3206 (2.55%) | 8033 (21.40%) | 11,239 (6.88%) |
| Sister's cesarean deliveries, percent* | - | - | - |
| **Pregnancy characteristics** |  |  |  |
| Gestational Diabetes | 1,699 (1.35%) | 1,084 (2.89%) | 2,783 (1.70%) |
| Gestational Hypertension | 1,248 (0.99%) | 740 (1.97%) | 1988 (1.22%) |
| Gestational weight gain z-score | - | - | - |

Gestational weight gain z-score was calculated according to Hutcheon et al. 2013.
\*Sister's CD percent was calculated as the percentage of CDs over all sister's pregnancies which ended prior to the relevant participant's date of birth.

## 3. Baseline covariates definitions

The following list describes the generation mechanism for each of the baseline covariates extracted from the data. We defined “index date” as the date of birth. All baseline covariates were taken prior to the index date. Covariates were defined by a trained gynecologist and separated to three interest times; pre-pregnancy, during pregnancy and during labor\birth.

### A. Pre-pregnancy

a. Maternal diagnoses
  i. Diabetes diagnosis was defined by either the presence of one Diabetes code (ICD-9 codes of 250.xx), an Hemoglobin A1c test with value of at least 6.5% or a Glucose blood test with value of at least 200 mg/dL.
  ii. Hypertension was defined by the presence of at least one Hypertension code (ICD-9 codes 642.2X, 642.9X)
  iii. Diagnoses from previous pregnancies were defined by the presence of at least on ICD-9 code: shoulder dystocia (ICD-9 codes 660.4X) perineal tear (stage 3 or more) (ICD-9 codes 664.2X, 664.3X, 664.6X).
b. Maternal Anthropometrics
  i. Weight and height measurements were used for calculation of mean BMI and change in BMI before and during pregnancy.
  ii. Systolic and Diastolic Blood pressure mean values.
c. Maternal Ancestry: 15 features breaking down the origin of the patient’s ancestors, as logged in their country of origin. World’s countries were clustered into 14 categories, corresponding to Israel’s major ancestry groups: North Africa, Iraq, Iran, Yemen, East Europe, West Europe, ex-USSR, North America, Latin America, Arab, Mediterranean, Ethiopia, Asia and Africa. Another feature logs the percentage of unknown origin. The distribution of the different ancestry indices in the cohort is presented in Fig. S3.1.
d. Maternal socio-economic data: Although personal socio-economic data were not available, we generated estimates using the locality of most of the patient’s clinic visits and data available by Israel’s Central Bureau of Statistics. Features include locality type (length 20, 1-hot vector) and locality religion breakdown (length 5, summing to 1 vector).
e. Maternal and paternal age at birth.
f. Maternal family cesarean section delivery prevalence: Utilizing the family connectivity of the data we constructed a feature which summarizes the percentage of cesarean section deliveries amongst the mother’s sisters, which can affect the risk of the mother to give birth in a cesarean section delivery ^63^ Obstetrical history: Number of previous deliveries and number of previous cesarean deliveries.

**Figure S3.1:**
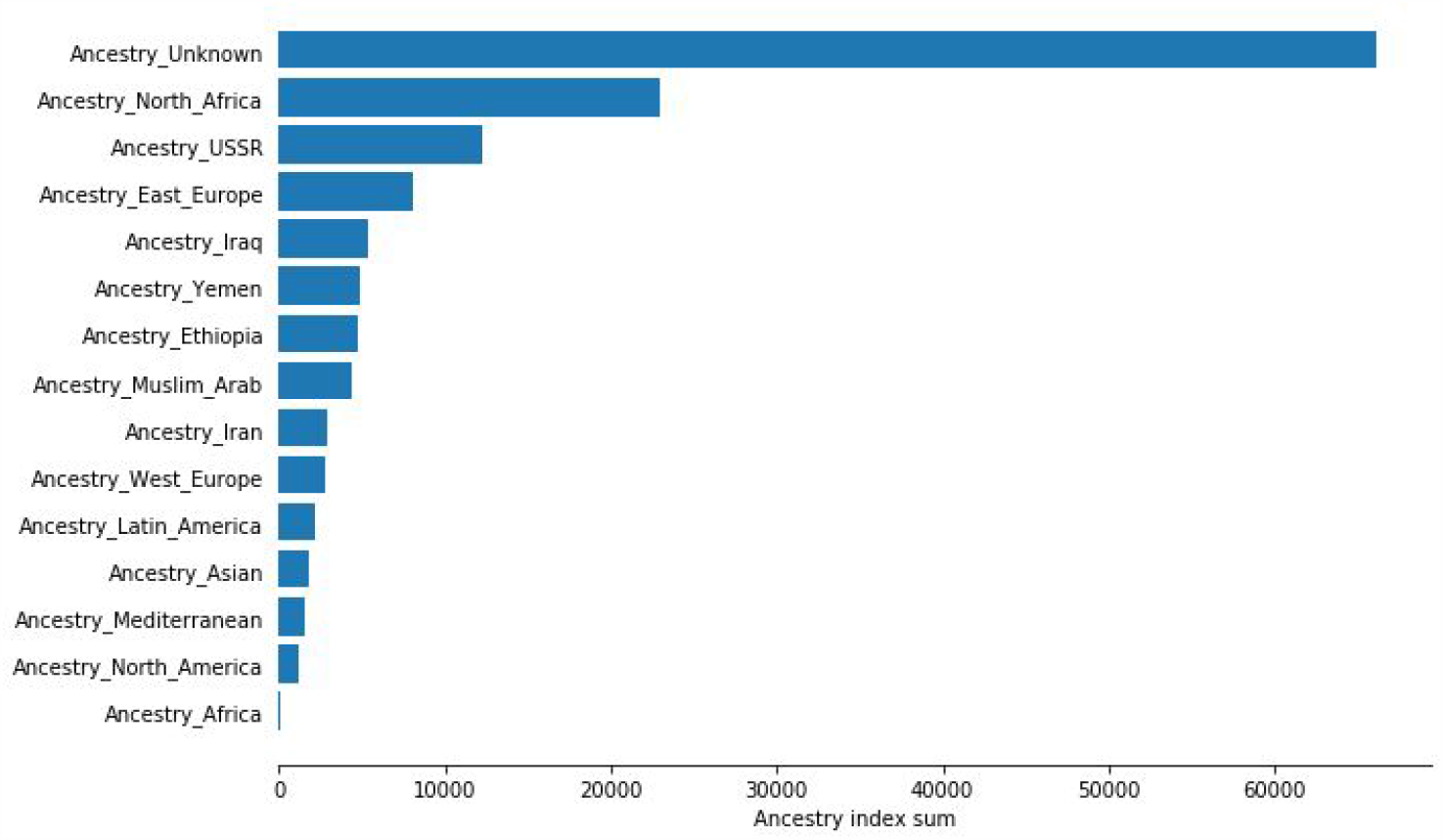
The distribution of the different maternal ancestries index in the cohort.

### B. During pregnancy

a. Gestational weight gain z-score was calculated using reference weight gain percentiles ^64^.
b. Maternal\fetal Diagnoses in the form of ICD9 codes during pregnancy were manually sorted by an Obstetrician. Diagnoses taken were those with medical relevance to birth type (vaginal\CD). Each diagnosis was defined by the presence of at least one ICD-9 code: Hypertensive disorders in pregnancy (ICD-9 codes 64233, 64232, 64231, 6423, 6423), major fetal anomalies (ICD-9 codes 6537, 65370, 655, 6558, 65580, 65581, 65582, 65583, 6559, 65590, 65591, 65592, 65593, 65594, 656, 65680, 65681, 65682, 65683, 65690, 65691, 65693, 6597, 65970, 65971, 65973, 678, 7429, 7439, 74400, 74403, 7443, 74600, 7462, 7469, 74720, 74740, 74760, 74761, 74762, 74763, 74764, 74782, 7479, 74860, 7489, 75010, 7509, 75160, 75240, 7529, 7539, 75550, 75560, 7559, 7561, 75610, 7579, 7589, 7599, 76381, 76383, 7780, V8903), Macrosomia (ICD-9 codes 7660, 7661, 766, 65664, 65663, 65662, 65661, 65660, 6566), malpresentation\malposition (ICD-9 codes 652.XX, 660.X), placental abruption (ICD-9 codes 641.X), Oligohydramnios (ICD-9 codes 658.0X) and Polyhydramnios (ICD-9 codes 657.X), Preeclampsia (ICD-9 codes
c. Gestational diabetes (GDM) was defined by the results of a glucose challenge test. Either a Glucose challenge test of 50g with a measurement above 200 mg/dL or a Glucose challenge test of 100g with one glucose measurements above the thresholds of 95, 180, 155 and 140 mg/dL under fasting conditions - fasting, one, two and three hours after glucose intake, respectively, were considered as a positive GDM diagnosis.

### C. Labor\birth

a. Diagnoses which occur during labor which are medically relevant to birth type (vaginal\CD) were selected by a gynecologist, and defined by the presence of at least one ICD-9 code in the hospital’s records: Non reassuring fetal heart rate (NRFHR) (ICD-9 codes 656.3X, 659.7X), arrest of descent\arrest of dilatation (ICD-9 codes 652.5X, 660.9X, 660.8X, 660.6X), failed induction (ICD-9 codes 659.X) and failed vacuum\forceps (ICD-9 codes 660.7X).
b. Gestational age: For children born in a Clalit hospital after 2012, gestational age information was available in the data. For women with available GCT results, performed routinely in Israel at 24-28 weeks of gestation, GCT result was used as a proxy for week 26 of gestation and then used for approximation of gestational age. Otherwise median gestational age in the Clalit birth data was taken - 39 weeks.
c. Hospital code.
d. Newborn birth weight.
e. Newborn sex.
f. Birth weekday and year day (Fig. S3.2).
g. Birth hour of day (Fig. S3.3).
h. Admission hour of the day (Fig. S3.3).

**Figure S3.2:**
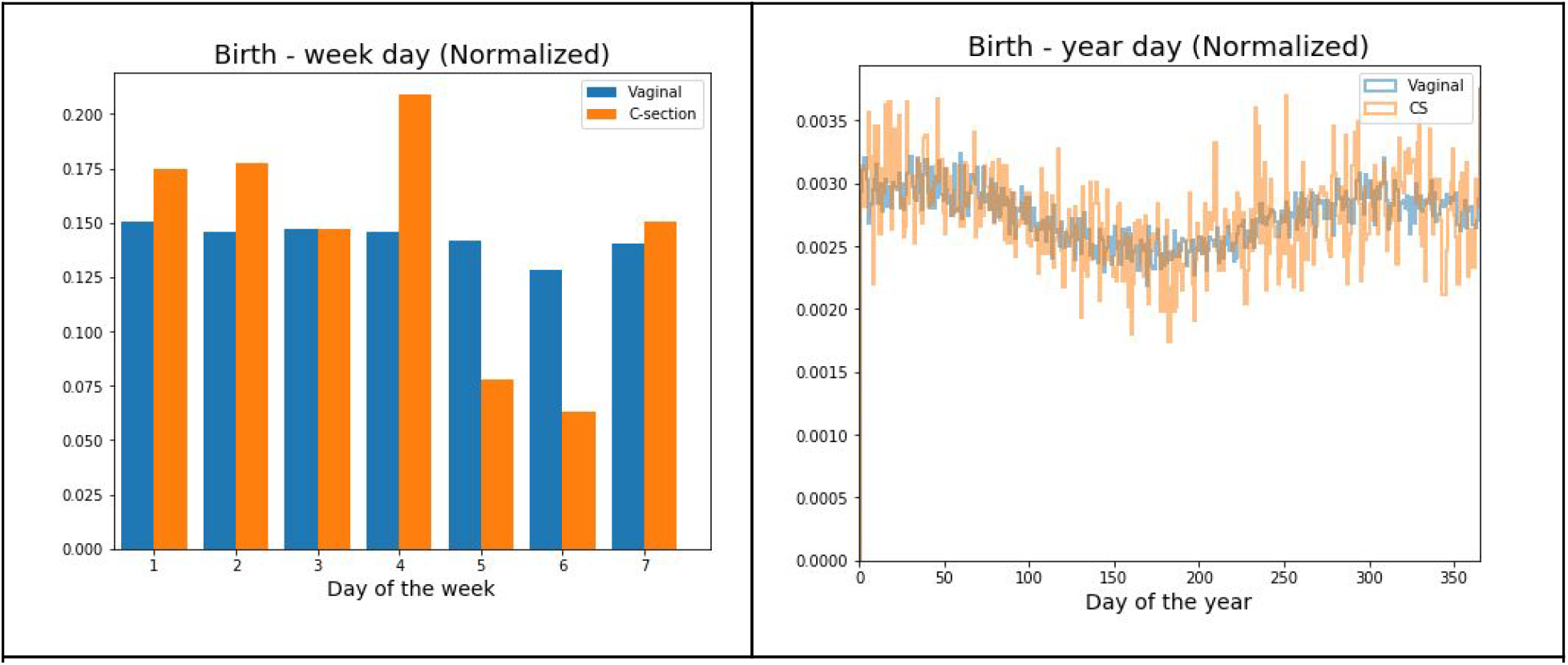
**Left:** Normalized histogram of birth week day. **Right:** Normalized distribution of birth year day

**Figure S3.3:**
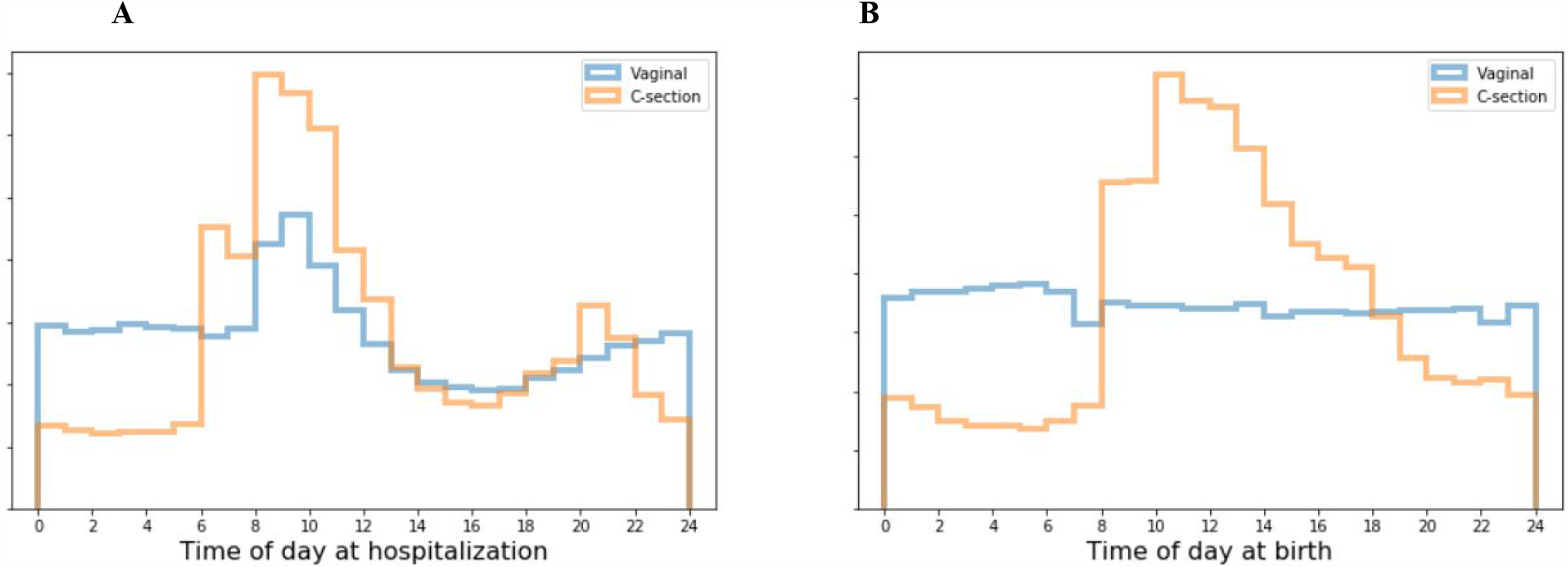
Normalized distribution of birth related time features. **A**. Time of admission to the hospital. **B**. Time of birth.

## 4. Pediatric outcomes definitions

First, potential long term childhood outcomes of cesarean section delivery were identified by previous studies that demonstrated correlations between these outcomes and delivery by cesarean section ^1,7^. Second, each outcome was defined separately by a trained pediatrician, as defined below.

Notably, the same definitions were also used in the UK cohort analysis. Diagnosis codes were converted from ICD9 and ICD10 to standardized SNOMED-CT codes. For outcome definitions requiring medication regimens, specific medications were added according to their generic and/or commercial names as used in the UK.

### A. Asthma

It has been previously demonstrated that identifying asthma cases in electronic health records is possible with high sensitivity and specificity, by combining multiple data sources ^65^. We defined asthma status based on the definition in the PheKB website https://phekb.org/phenotype/asthma-response-inhaled-steroids. As this definition refers specifically to patients with asthma who use inhaled steroids, we expanded the definition to all asthma drugs. More specifically, a diagnosis of asthma was considered if all of the following criteria were met:

1. At least 2 asthma codes on different days were present (ICD-9 codes of 493.xx)
2. At least 1 asthma drug was dispensed (Salbutamol (R03AC02), Terbutaline (R03AC03), Salmeterol (R03AC12), Seretide Cd (R03AK06), Symbicort/Duoresp (R03AK07),, Beclometasone (R03BA01), Budesonide (R03BA02), Fluticasone (R03BA05), Ipratropium Bromide (R03BB01), Salbutamol (R03CC02), Terbutaline (R03CC03), Montelukast (R03DC03), Budesonide (R01AD05), Fluticasone (R01AD08).
3. Children with the following diagnoses were censored for the asthma outcome: Cystic Fibrosis 277.00 - 277.02, Immunodeficiency 279.xx, Bronchiectasis 494.0 - 494.1, Hereditary and degenerative diseases of CNS (331), Mental Retardation (317,318,319), Congestive Heart Failure 428-429.9, Pulmonary Hypertension, Embolism 415.11-415.19, 416.0 - 416.8, 417.XX, COPD 496, lupus (710.0), RA (714.0), Tuberculosis (011.1), Lung cancer (162.*), Sarcoidosis (135)

**Figure S4.1:**
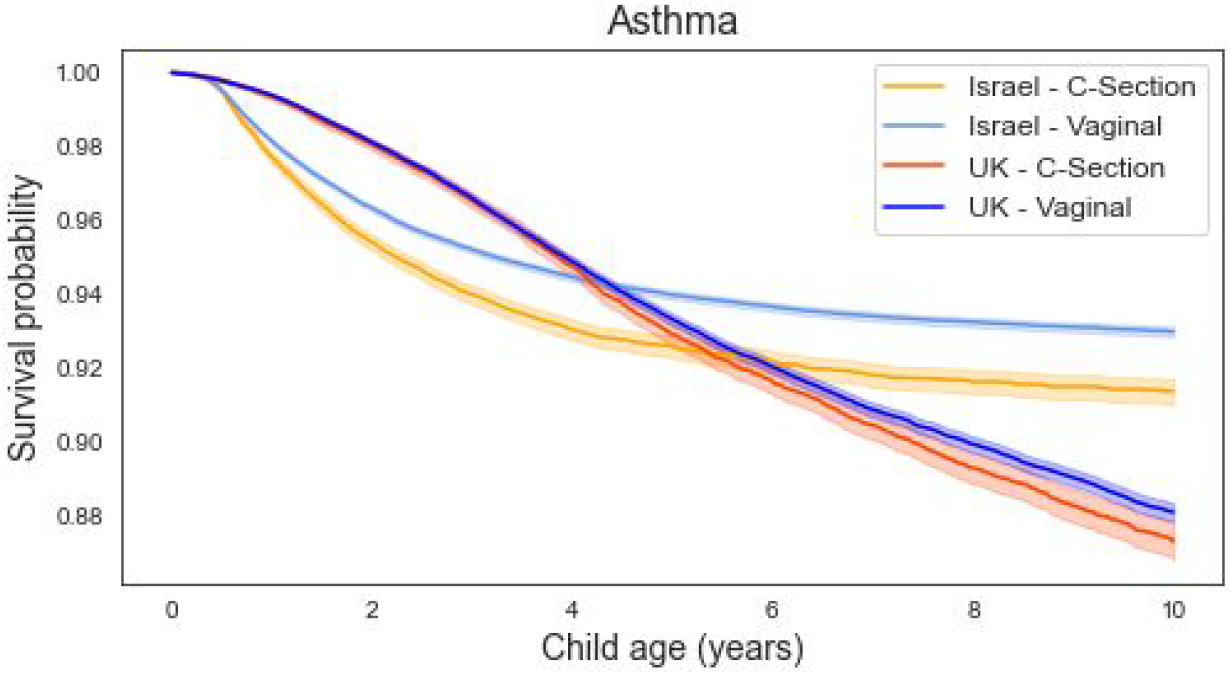
Kaplan-Meier unadjusted curves for asthma outcome in children born in vaginal delivery or CD, in Israel (orange and light blue accordingly) and UK cohorts (red and dark blue accordingly).

### B. Overweight\Obesity

We defined obesity status in accordance with health care professionals in Israel, using the CDC BMI reference percentiles ^36^. Cutoffs for normal weight, overweight, and obesity were determined using the CDC’s standard thresholds of the 85th percentile for overweight and 95th percentile for obesity. Of note, it was previously shown similar estimates of obesity risk at 5 years of age as other percentile curves such as The World Health Organization (WHO) WFL ^66^. Valid measurements were defined as being in the range of 5 CDC standard deviation scores for weight and height.

### C. Atopic dermatitis

Was defined as the presence of at least 2 atopic dermatitis codes on different days (ICD-9 codes 6918, S87, 691).

**Figure S4.2:**
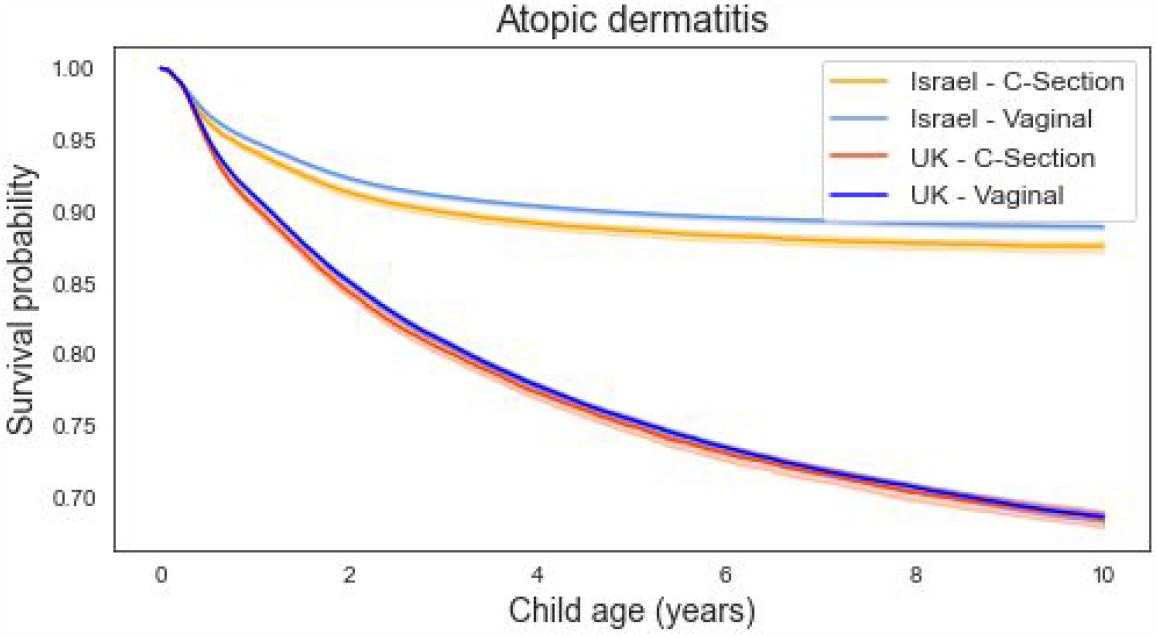
Kaplan-Meier unadjusted curves for atopic dermatitis outcome in children born in vaginal delivery or CD, in Israel (orange and light blue accordingly) and UK cohorts (red and dark blue accordingly).

### D. Celiac disease

Was defined as the presence of at least 2 Celiac codes on different days (ICD-9 code 5790).

**Figure S4.3:**
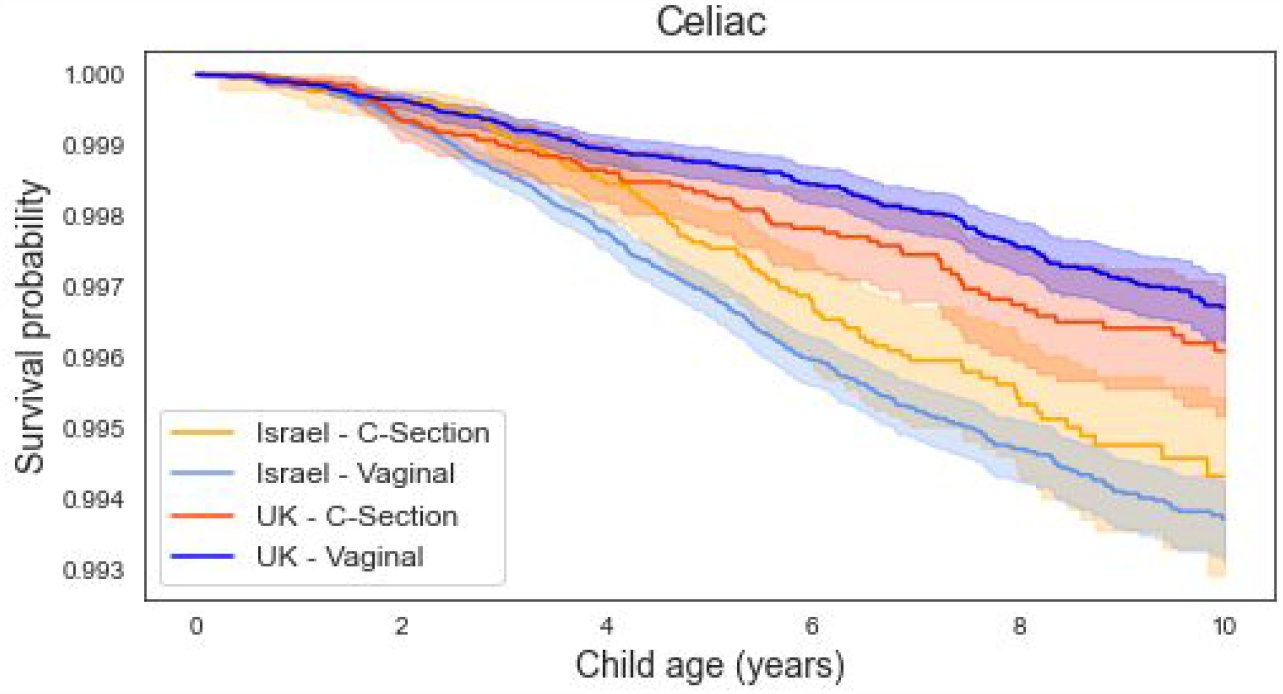
Kaplan-Meier unadjusted curves for celiac outcome in children born in vaginal delivery or CD, in Israel (orange and light blue accordingly) and UK cohorts (red and dark blue accordingly).

### E. Allergy

The task of clinical coding in allergy is complex, as allergic disorders have a wide range, they occur across the entire life course, and there are several organ systems that can be affected ^67^. Allergy was defined by the presence of one of the following certeira:

1. At least one ICD-9 diagnosis (expanded definition based on Hirsch et al. ^68^), including: Food allergy cases: milk allergy ICD-9 code (V1502, 99567) and Non-milk food allergy cases (e.g. nuts, seafood) ICD-9 code (V1501, V1503, V1504, V1505, 6931, 99560, 99561, 99563, 99564, 99565, 99566, 99568, 99569, 9957), drug allergy ICD-9 code (V140, V141, V142,V143, V144, v145, V146, V147, V148, V149, 99527, V1508), other allergies (e.g. Latex, Insects), allergic rhinitis, allergic purpura,(allergic urticaria, allergic conjunctivitis, allergic pneumonitis, allergic arthritis, allergic gastroenteritis ICD-9 code (9953, 4770, 4772, 4778, 4779, A12, 7080, R97, 2870, 5583, 7162, 38104, 37214, 4959, 4958, 38105, 71626, Z9912, V1506, v1507, v150, V1509, V1381.
2. At least one prescription of the drug Epipen (ATC codes:C01CA24)

**Figure S4.4:**
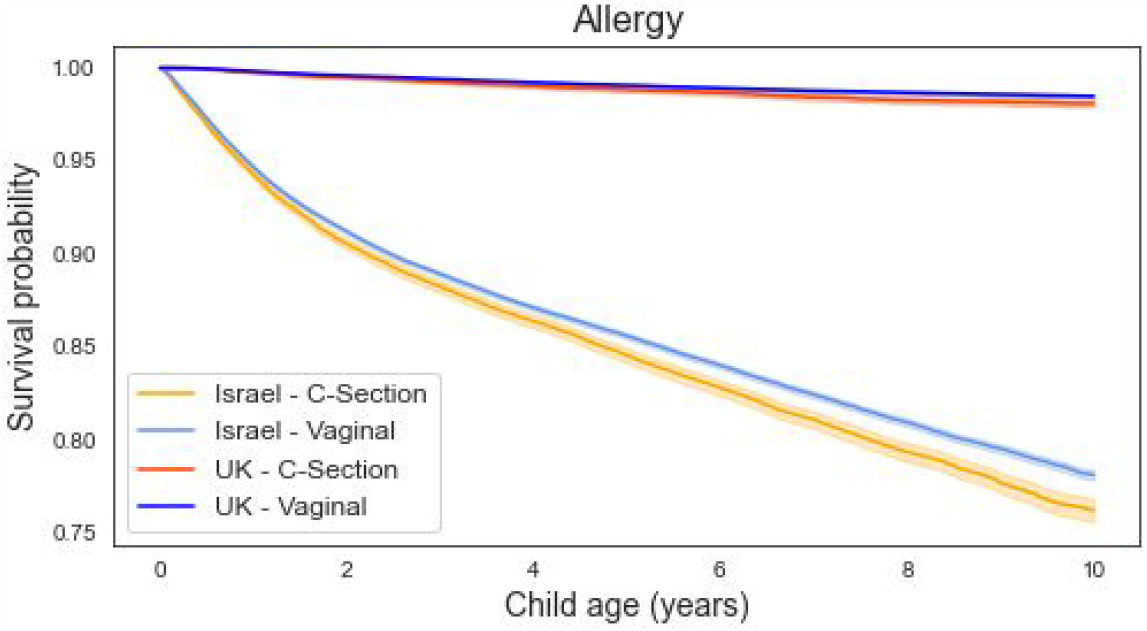
Kaplan-Meier unadjusted curves for allergy outcome in children born in Vaginal delivery or CD, in Israel (orange and light blue accordingly) and UK cohorts (red and dark blue accordingly).

### F. Autistic spectrum disorder (ASD)

Was defined based on a previous study ^69^. As information from clinical notes was not available as part of our data, ASD was defined as follows: at least 1 ICD-9 codes for ASD, Asperger’s or Pervasive Developmental Disorder-Not Otherwise Specified (PDD-NOS), ICD-9 codes (299.0, 299.80,299.9, F84.0, F84.1, F84, F84.9, F84.5, F84.8)

Children with the following diagnoses were censored for the ASD outcome: Childhood disintegrative disorder (ICD-9 code 299.1), Schizophrenia (ICD-9 codes 295.X), Tuberous Sclerosis (ICD-9 code 759.5), Fragile X Syndrome (ICD-9 code 759.83), Rett’s syndrome (F84.2) and Other specified cerebral degenerations in childhood (ICD-9 code 330.8).

**Figure S4.5:**
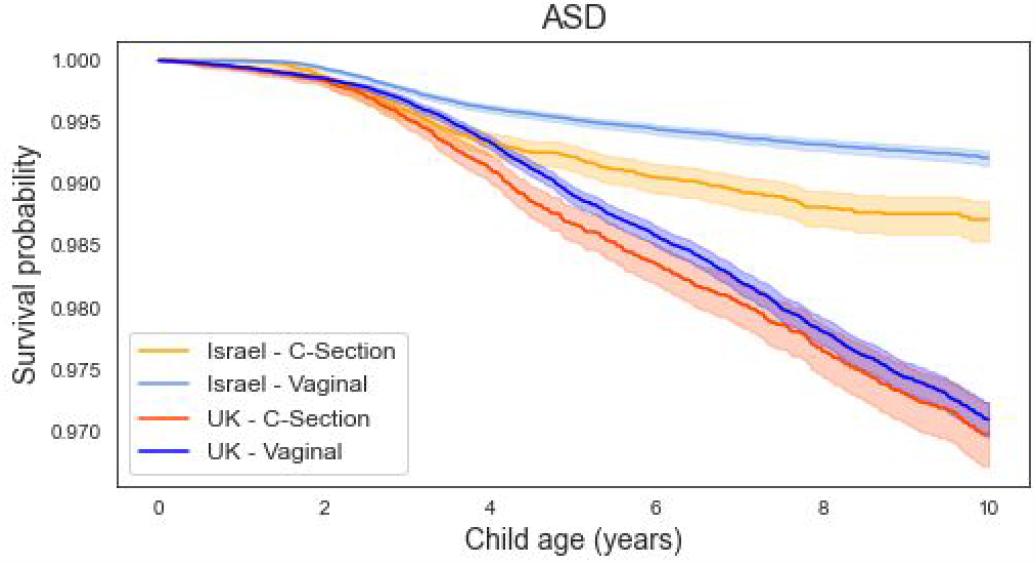
Kaplan-Meier unadjusted curves for ASD outcome in children born in vaginal delivery or CD, in Israel (orange and light blue accordingly) and UK cohorts (red and dark blue accordingly).

### G. ADHD

We defined ADHD based on the definition in the PheKB website, based on the following study ^70^. ADHD cases must be 4 years of age or older, and are defined by a diagnostic history of ADHD as determined by the relevant ICD9 codes and/or a history of ADHD medications, based on the following certeira:.

1. Age equals or greater than 4 years old and
2. At least 1 relevant ICD-9 diagnosis codes on different days was present (ICD-9 code 314.0, 314.01, 314.00, 7745, 314.2, F908, F90, F909, 314.9) and
3. Individual’s medical record includes one or more prescriptions of ADHD-related medications (as previously classified ^71^: methylphenidate (ATC code N06BA04), atomoxetine (N06BA09), amphetamine (N06BA01) and dexamphetamine (N06BA02)

Or:

1. Age equals or greater than 4 years and
2. At least two relevant ICD-9 diagnosis codes on different days were present ICD-9 code (314.0, 314.01, 314.00, 7745, 314.2, F908, F90, F909, 314.9)

Children with the following diagnoses were censored for the ADHD outcome: Dementia (2900, 2904, F03, F01), Somatoform disorder (F45, 30081, 30089, 3008, F453), Stereotyped Repetitive Movements (3073), Mental Retardation (317, F70, F71, 3182, F711, 3180, F721, F720, F701, F700, F708, F710, F728, F718, F73, F730, F731, F790), CNS Tuberculoma (132), CNS Malignancies (1983, 1917, 1910, 1911, 1912, 1913, 1914, 1915, 1916, 1917, 1918, 1919, 1920, 1922, 1928), Neurofibromatosis (23770, 23772, 2377,23771),Anoxic Brain Damage (3481), Benign Intracranial Hypertension (3482), Encephalopathy, Unspecified (3483), Compression Of Brain (3484), Cerebral Edema (3485), Temporal Sclerosis (34881), Brain Death (34882), Cerebral Calcification (348891), Other Conditions Of Brain (34889), Hypertensive Encephalopathy (4372), Other congenital anomalies of nervous system (742-742.x), Slow fetal growth and fetal malnutrition (764-764.x), Subdural and cerebral hemorrhage (767.0), Birth trauma, unspecified (767.9), Fracture of skull (800-804), Head injury, unspecified (959.01).

**Figure S4.6:**
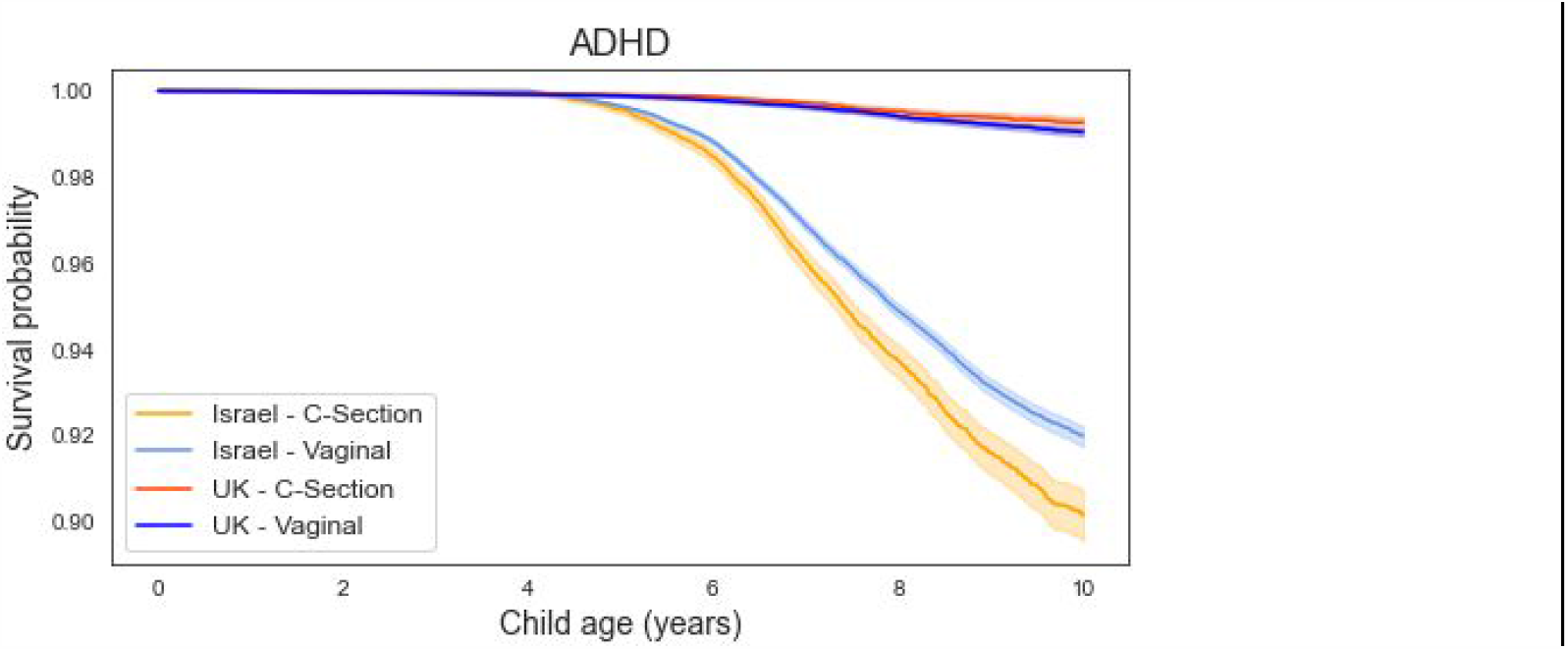
Kaplan-Meier unadjusted curves for ADHD outcome in children born in vaginal delivery or CD, in Israel (orange and light blue accordingly) and UK cohorts (red and dark blue accordingly).

### H. Inflammatory bowel disease

Was defined as the presence of at least two relevant ICD-9 diagnosis on different days. Ulcerative colitis (UC) ICD-9 codes 556.X and crohn’s disease (CD): ICD9 - 555.X

**Figure S4.7:**
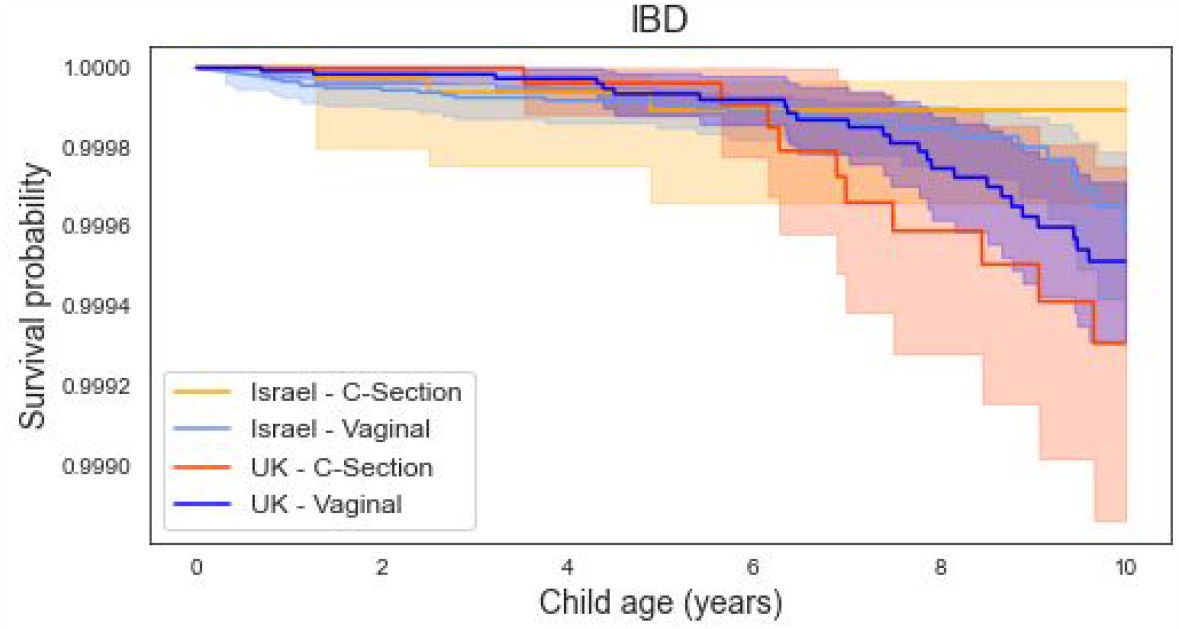
Kaplan-Meier unadjusted curves for IBD outcome in children born in vaginal delivery or CD, in Israel (orange and light blue accordingly) and UK cohorts (red and dark blue accordingly).

### I. Type 1 diabetes

We defined Type 1 diabetes based on the definition in the PheKB website, based on the following Study ^72^, with an adaptation, as it was originally used for diagnosing both type 1 & 2 diabetes. We defined that a patient has T1DM based on the following criteria:

1. One or more outpatient diabetes-related ICD-9- diagnosis codes (250, 2500, 2501, 25001, 25011, 25013,25003, 2504, 25061, 25081, 2508, 25083, 25091, 25093) as well as a local Clalit diagnosis code “Diabetes Routine Follow Up” (8). Or
2. At least two prescriptions of Insulin (ATC code A10AB0X) And

Hemoglobin A1c (HbA1C) > 6.4 in at least one blood test

Children with the following diagnoses were censored for the T1DM outcome: Adult-onset Type Diabetes Mellitus, Neonatal Diabetes Mellitus, Mody Mature Onset Diabetes In Young And Secondary Diabetes Mellitus (ICD-9 codes 25000, 25080, 25010, 7751, 25050, E118, 24901, 25020, 24900, 25012, 25002).

**Figure S4.8:**
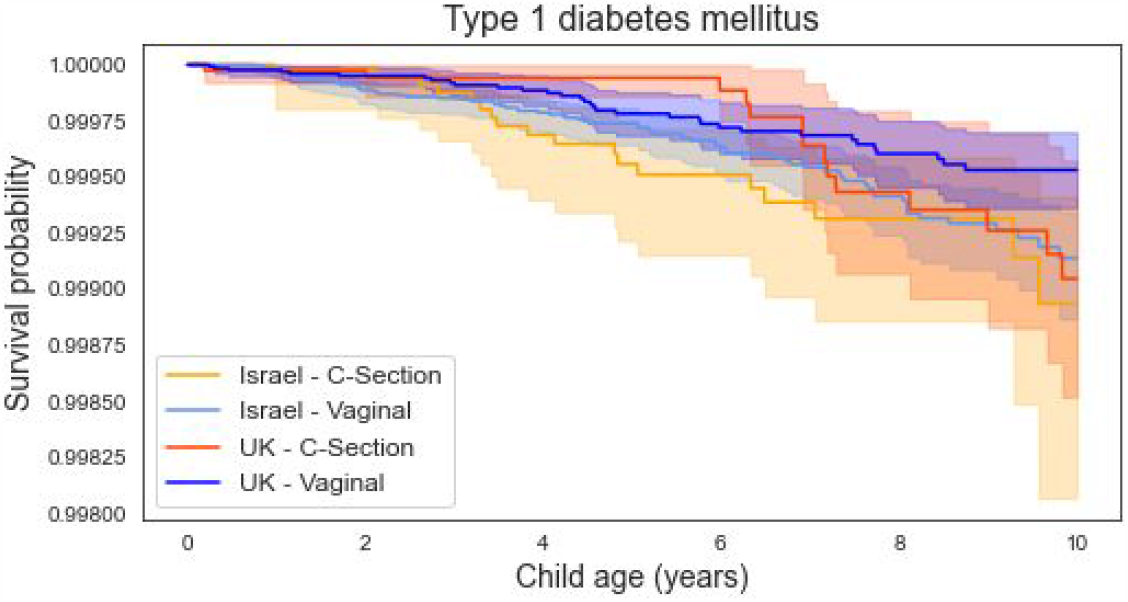
Kaplan-Meier unadjusted curves for type 1 diabetes outcome in children born in vaginal delivery or CD, in Israel (orange and light blue accordingly) and UK cohorts (red and dark blue accordingly).

### J. Juvenile idiopathic arthritis

Was defined as the presence of at least 2 relevant ICD-9 codes on different days (7143, 71430, 71432, 71433, and 71431).

**Figure S4.9:**
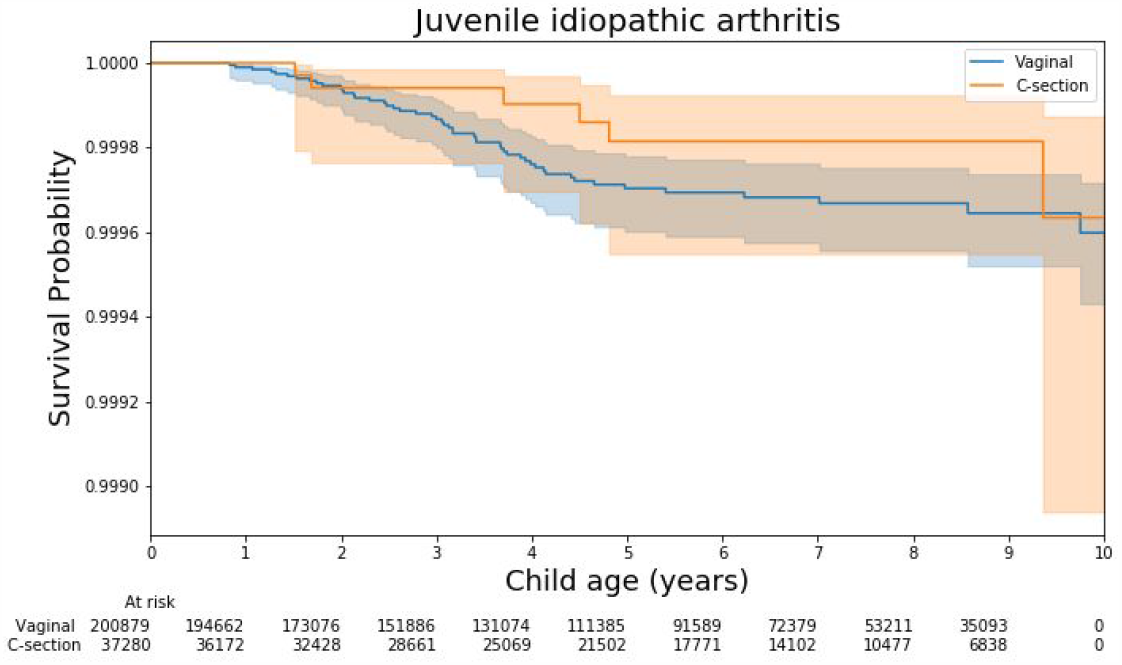
Kaplan-Meier unadjusted curves for type 1 diabetes outcome in children born in vaginal delivery (blue) or CD (orange) in Israel cohort.

### K. Respiratory infections

Acute respiratory infection (ARI) was defined as the number of the following ICD-9 diagnosis codes until the age of 5 years ^73^: nonspecific URIs (ICD-9-CM 460.X, 464.X, and 465.X), otitis media (ICD-9-CM 381.X (with the exception of 381.6 and 381.7) and 382.X, sinusitis (ICD-9-CM 461.X and 473.X), pharyngitis (ICD-9-CM 034.0, 462, and 463), acute bronchitis (ICD-9-CM 466 and 490), pneumonia (ICD-9-CM 481–486), and influenza (ICD-9-CM 487) in addition to local Clalit diagnostic codes (R81 pneumonia, H72 serous otitis media, R74 URI, H71 Acute OM

Children with the following ICD-9 codes were censored from the Respiratory infections outcome: Cystic Fibrosis 277.00 - 277.02, Immunodeficiency 279.xx, Bronchiectasis 494.0 - 494.1, Hereditary and degenerative diseases of CNS (331), Mental Retardation (317,318,319), Congestive Heart Failure 428-429.9, Pulmonary Hypertension, Embolism 415.11-415.19, 416.0 - 416.8, 417.XX, COPD 496, lupus (710.0), RA (714.0), Tuberculosis (511.1), Lung cancer (162.*), Sarcoidosis (135)

### L. Autoimmune disease

Many autoimmune diseases are very rare in the pediatric population. To identify autoimmune diseases in our data, we first extract an initial list of autoimmune diseases as defined by ‘Autoimmune Disease’ (D001327) in the MeSH® hierarchy https://meshb.nlm.nih.gov/record/ui?ui=D001327, i.e., all the descriptors with a MeSH® Tree Number starting with C20.111.

A patient was considered as having Autoimmune diseases in the presence of at least 2 relevant ICD-9 codes on different days including: Addison Disease [C20.111.163] - ICD-9 code 25541, Anemia, Hemolytic, Autoimmune [C20.111.175] - ICD-9 code 2830, Graves Disease [C20.111.555] - ICD-9 codes 2420, 24200, Hepatitis, Autoimmune [C20.111.567] - ICD-9 code 57142, Lupus Erythematosus, Systemic [C20.111.590] - ICD-9 codes 7100, 6954, Pemphigus [C20.111.736] - ICD-9 code 6944, Purpura, Thrombocytopenic, Idiopathic [C20.111.759] - ICD-9 code 28731, Thyroiditis, Autoimmune [C20.111.809] - ICD-9 code 2452, Undifferentiated Connective Tissue Diseases [C20.111.904] - ICD-9 code 7109, Autoimmune Diseases of the Nervous System [C20.111.258] included Multiple sclerosis - ICD-9 codes: 340, 3400, Acute inflammatory demyelinating polyneuropathy (Guillain-Barré syndrome) - ICD-9 code 3570 and Myasthenia Gravis - ICD-9 codes 3580 and 35800.

The following diseases were diagnosed as described above: Arthritis, Juvenile [C20.111.198] - section J, Diabetes Mellitus, Type 1 [C20.111.327] - Section I, Celiac disease - Section D.

**Figure S4.10:**
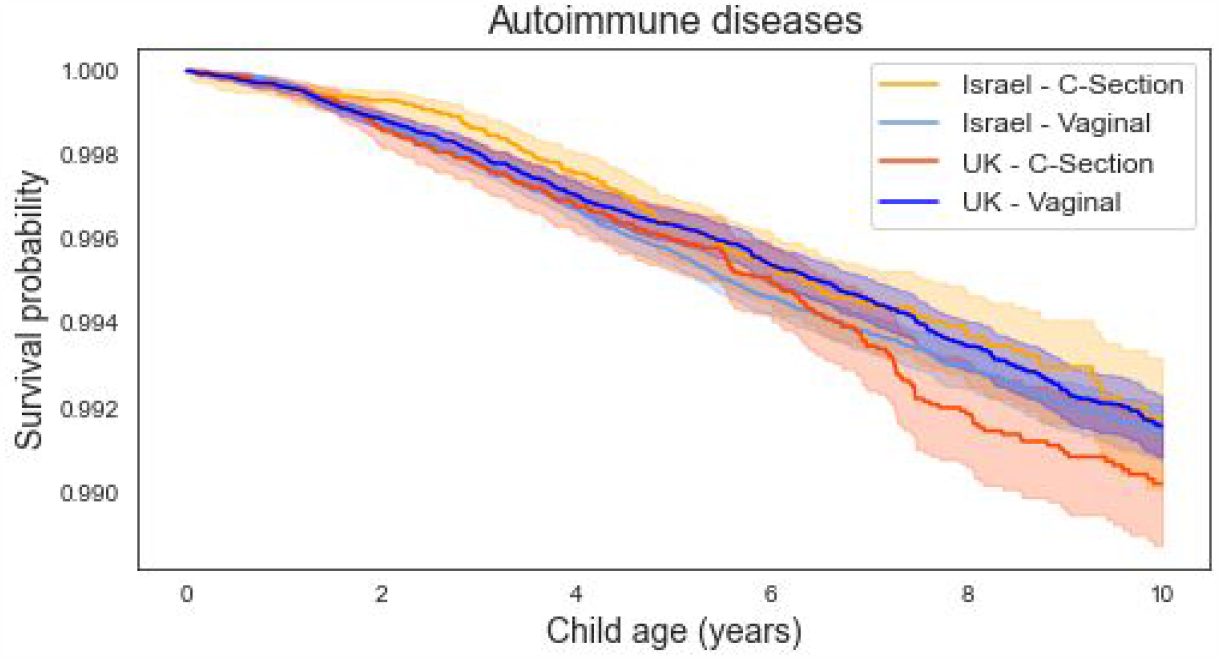
Kaplan-Meier unadjusted curves for autoimmune diseases outcome in children born in vaginal delivery or CD, in Israel (orange and light blue accordingly) and UK cohorts (red and dark blue accordingly).

### M. Atopy

Was defined as children who fulfilled the criteria described above for one of the following phenotypes: asthma (Section A), allergy (Section E) and atopic dermatitis (Section C).

**Figure S4.11:**
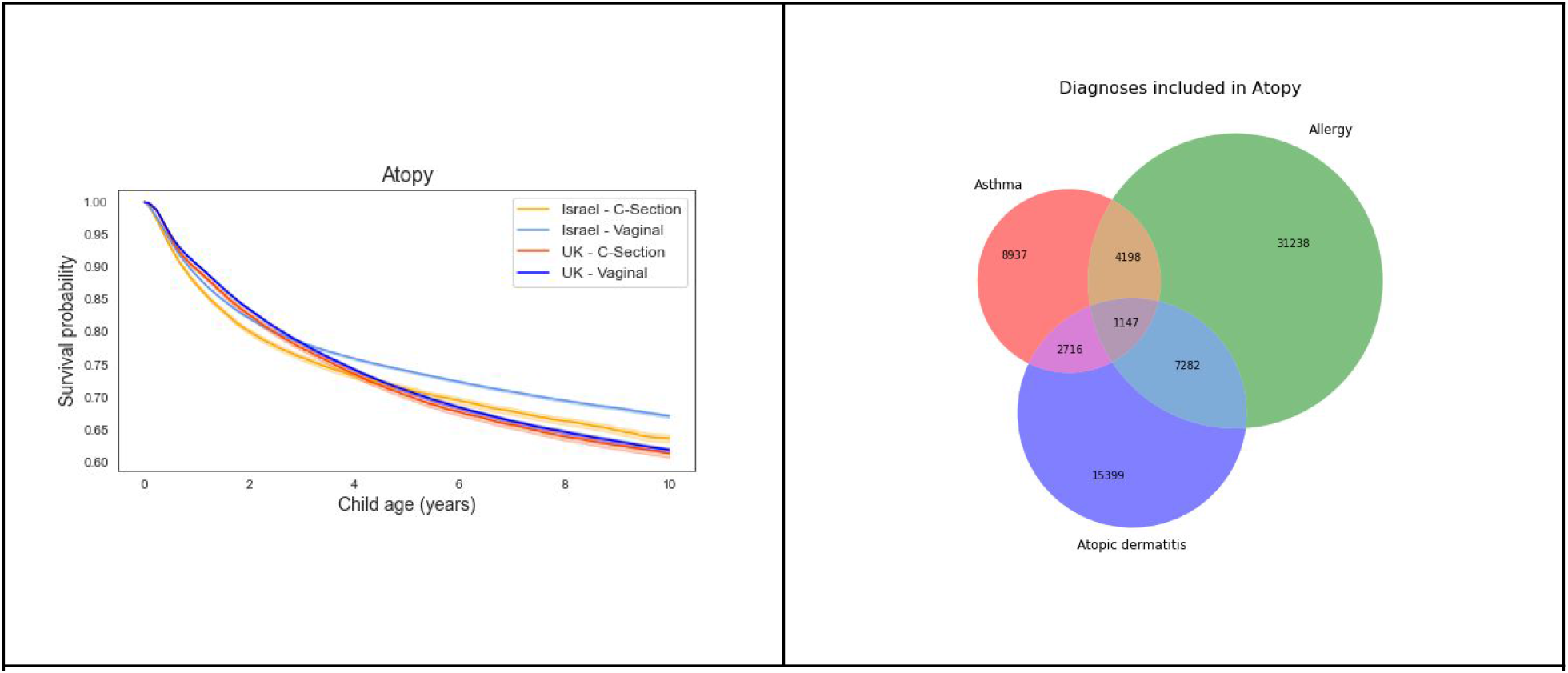
**Left:** Kaplan-Meier unadjusted curves for ADHD outcome in children born in vaginal delivery or CD, in Israel (orange and light blue accordingly) and UK cohorts (red and dark blue accordingly). **Right:** Venn diagram of the outcomes included in the atopy outcome, from Israel cohort.

### N. Negative Controls

The following pediatric outcomes were defined as negative controls, as they are not expected to be affected by c/s delivery compared to a normal vaginal delivery according to existing medical literature.

1. Fracture of upper end of radius and ulna, closed - ICD-9 code 8130.

**Figure S4-12:**
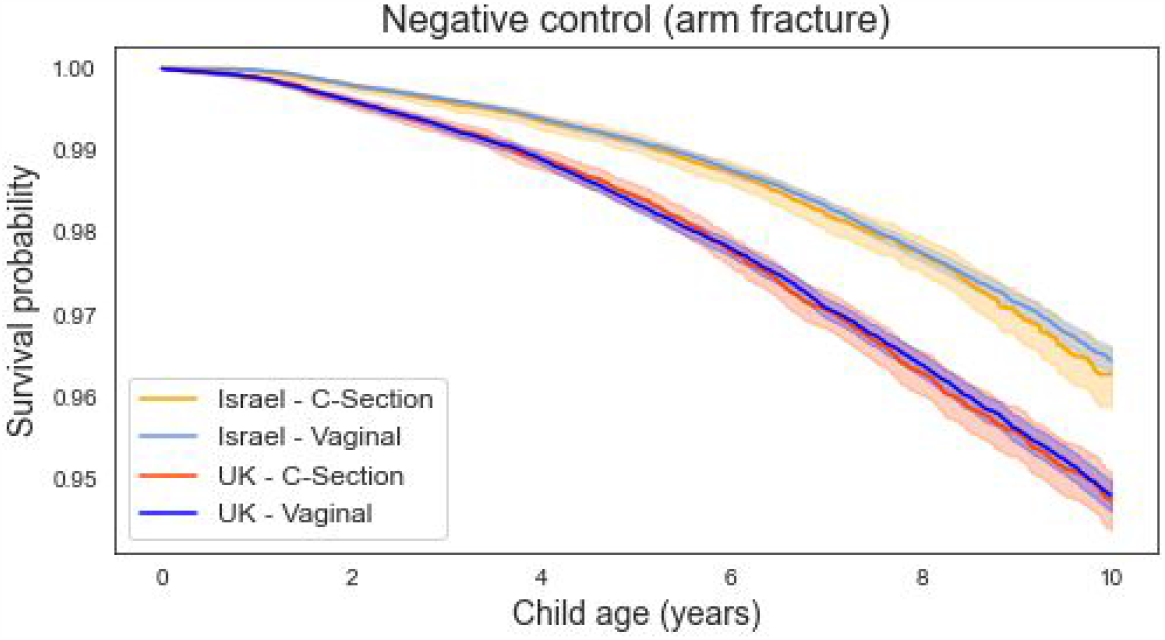
Kaplan-Meier unadjusted curves for arm fracture (negative control) outcome in children born in vaginal delivery or CD, in Israel (orange and light blue accordingly) and UK cohorts (red and dark blue accordingly).

## 5. Propensity model analysis

### Prediction model parameters

We used a gradient boosting trees model trained with the XGBOOST python package. Hyperparameters were selected following a cross-validated grid search, with the following settings selected:

- nuestimators= 500
- max_depth= 3
- colsample_bytree = 0.8
- subsample = 0.8

Missing values were not imputed prior to training the model. XGBOOST uses the method of *block propagation* in which tree splits are learned only from non-missing data, and only after that, the direction of splitting missing values is learned (by minimizing error) for the whole block of samples for missing values for that feature. Josse et al has shown that this procedure is a good option relative to various imputation methods ^74^.

**Figure S5.1:**
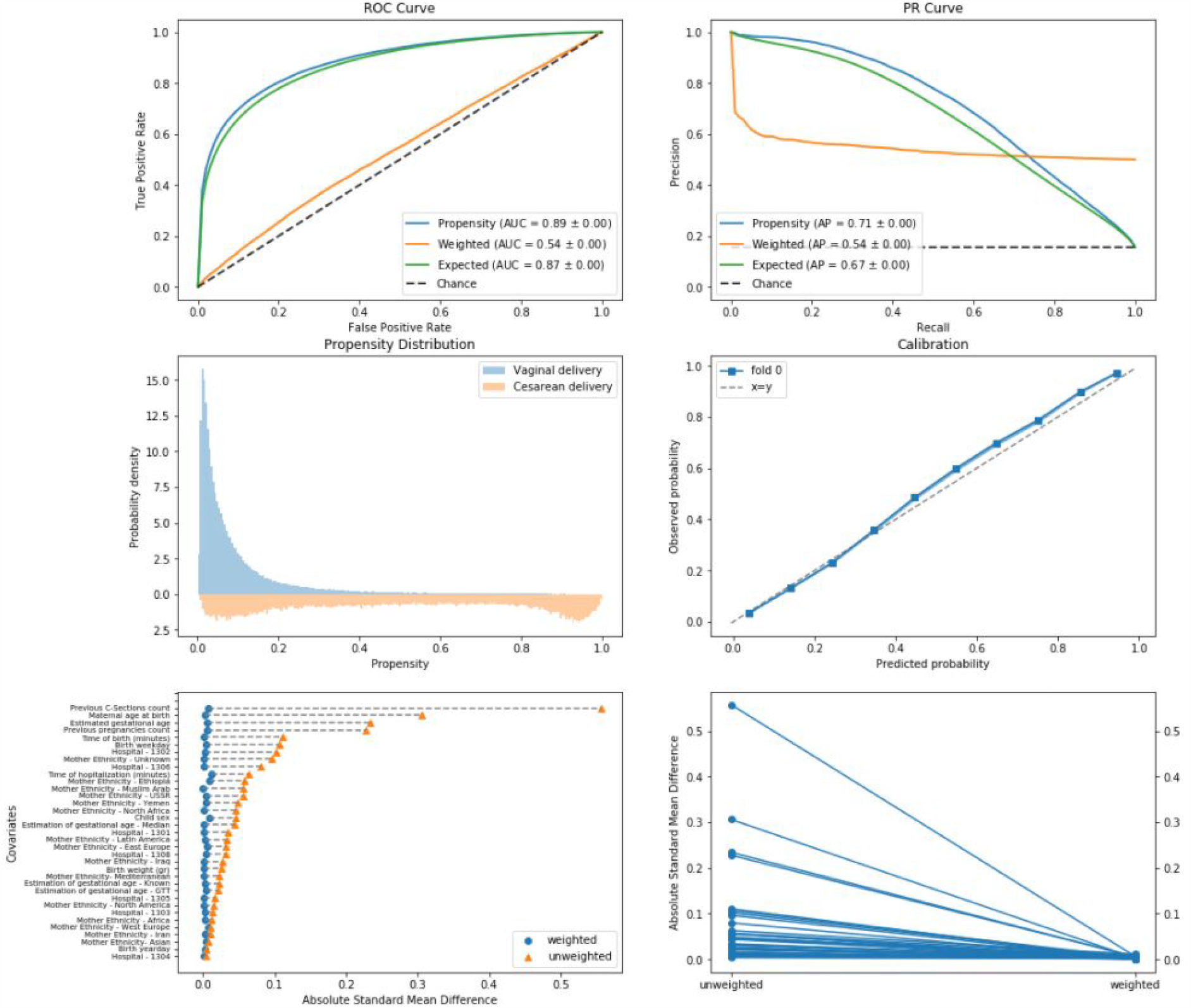
Propensity model evaluation on train set.

**Figure S5.2:**
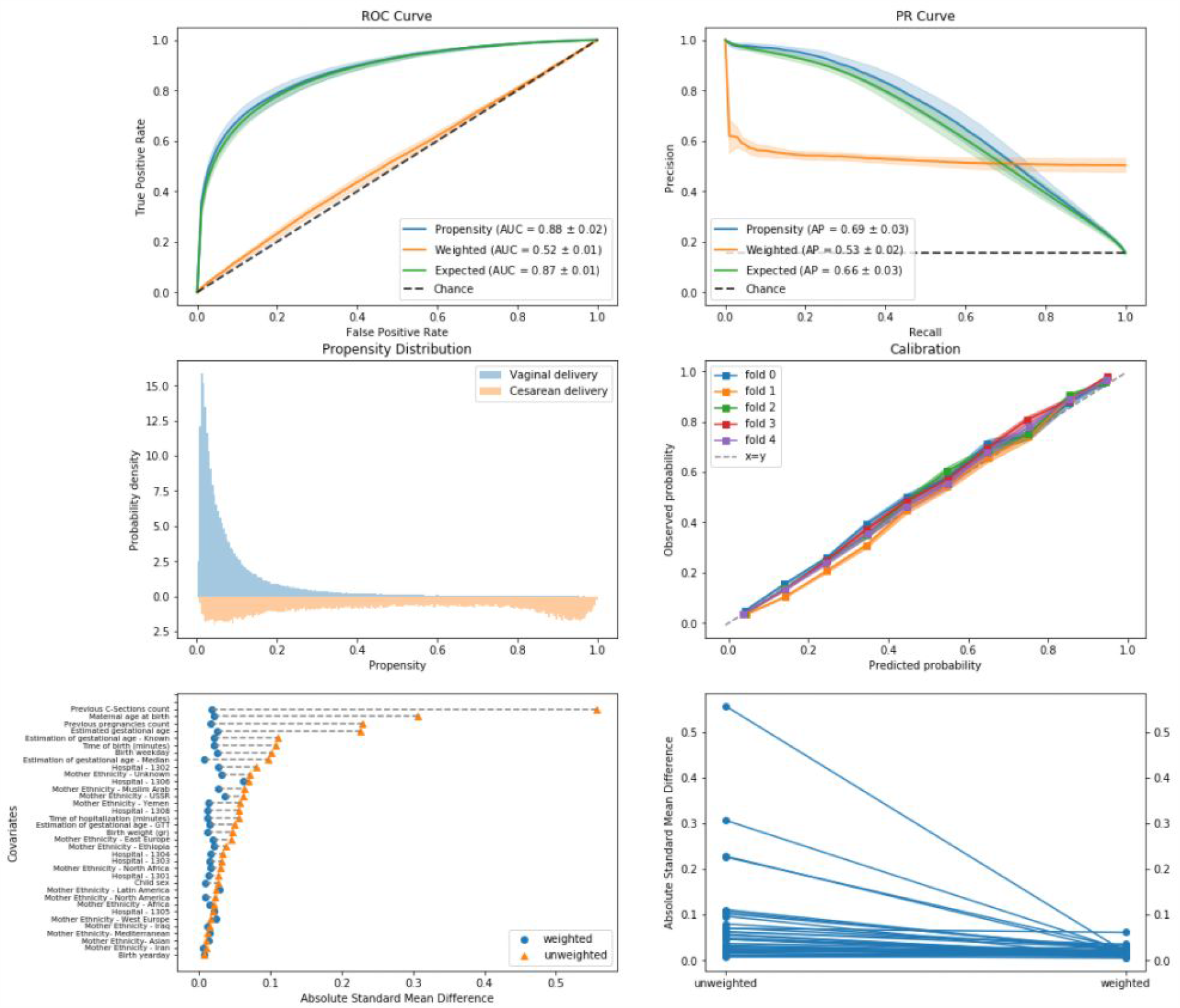
Propensity model evaluation on cross-validated test sets.

### Extended feature attribution analysis

**Figure S5.3:**
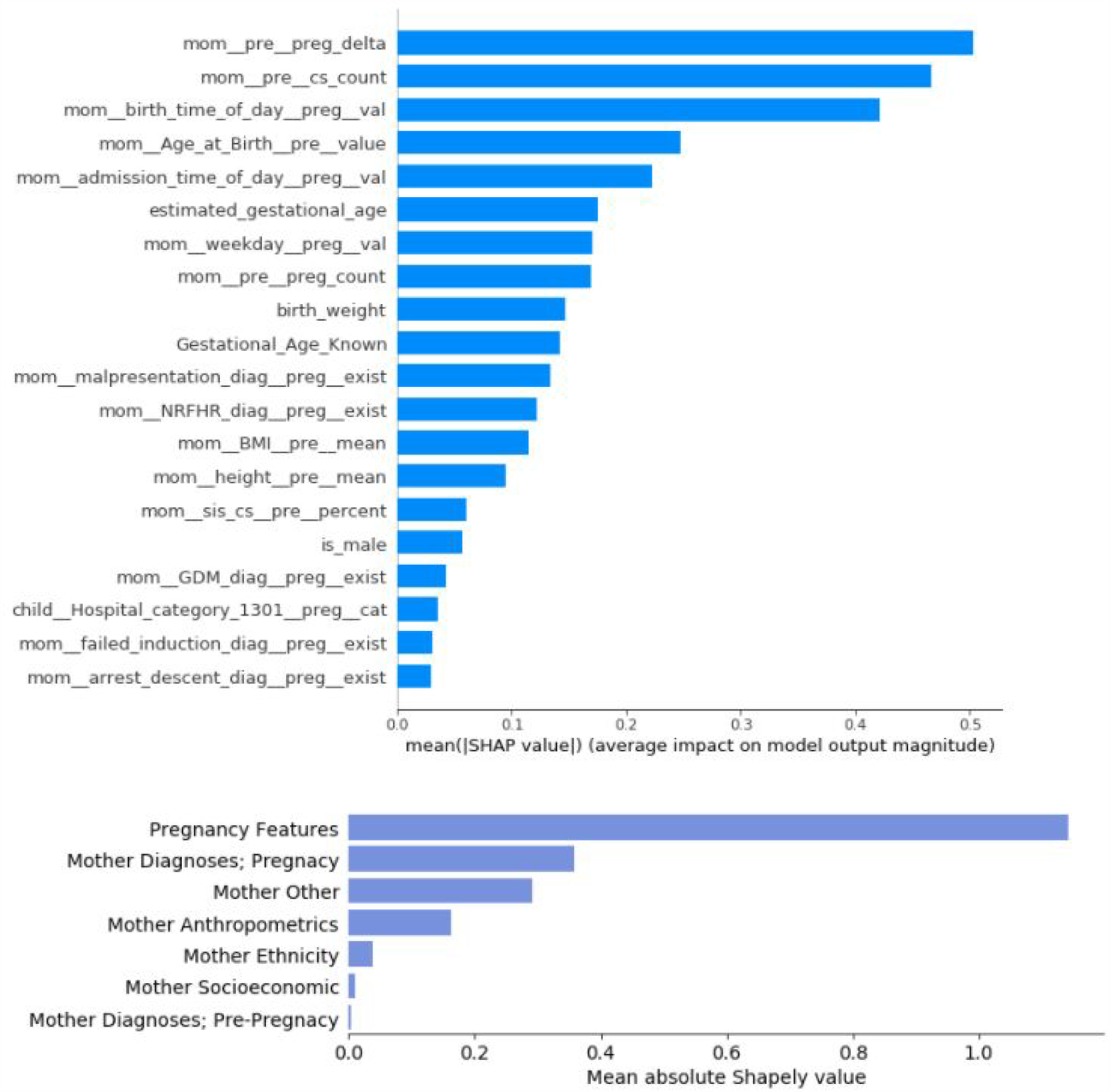
**A:** Mean absolute Shapley values (in log-odds scale) of the propensity prediction model. 20 top contributing features are shown. **B:** Mean absolute Shapley values (in log-odds scale) of groups of features. “Others” sums up remaining features not included in the rest of the groups such as blood pressure measurements and father’s age.

**Figure S5.4:**
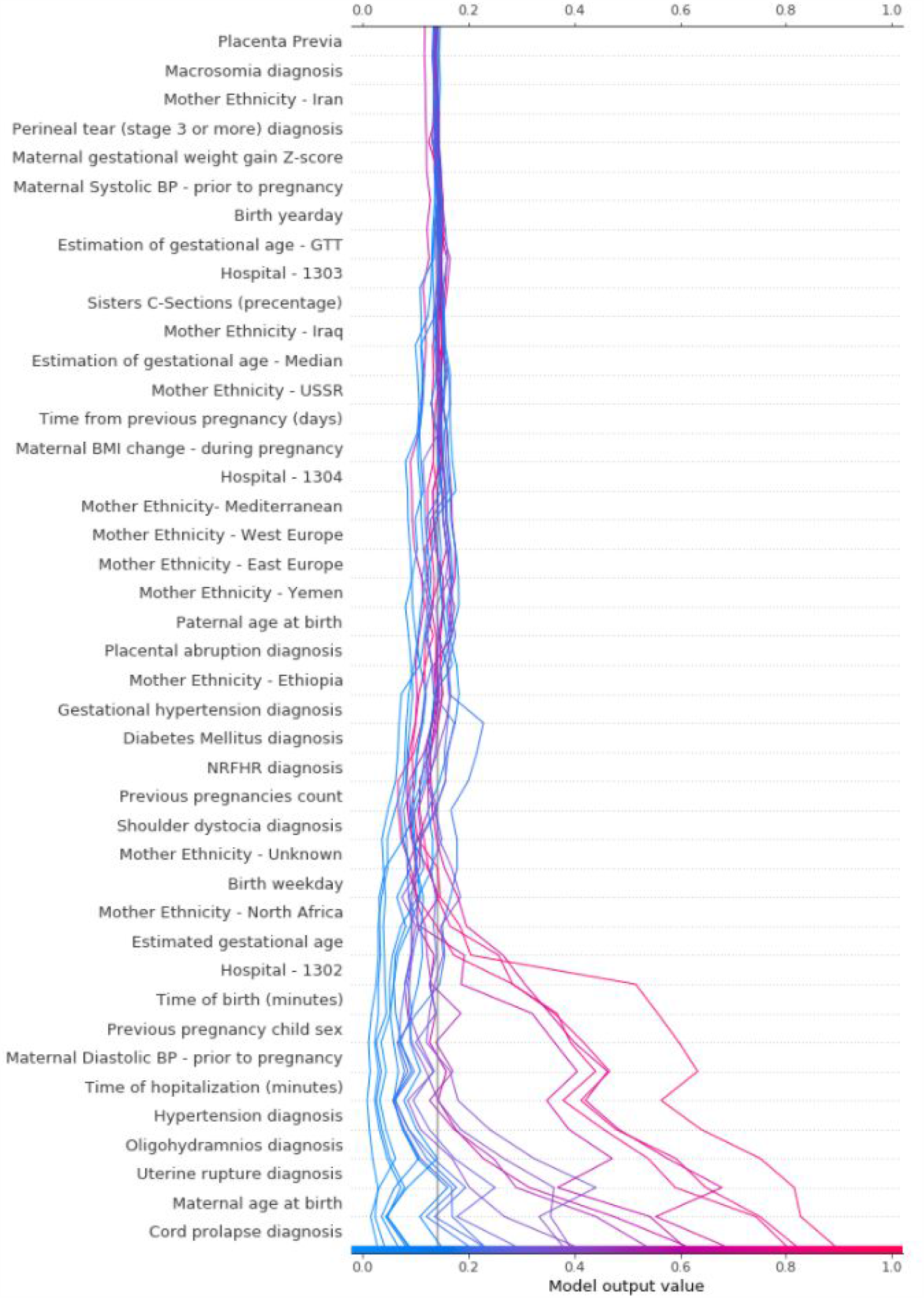
Decision plot of the probability of CD. A random sample of the population is presented. Each individual’s prediction is represented by a colored line. At the bottom, each line strikes the x-axis at its corresponding predicted probability. This value determines the color of the line. Moving from top to bottom, SHAP values for each feature are added to the model’s base value.

#### Weighting methods

Weighting methods aim to adjust for baseline confounders by creating a new weighted pseudo-population for which all variable distributions are identical in the treatment and control groups. These methods use the estimated propensity score, *e*_*i*_, to weigh each individual in the cohort. When using IPW the weight for each individual is defined as *w*_*i*_ = 1/*e*_*i*_ for treated units and *w*_*i*_ = 1/(1 − *e*_*i*_) for control units. When the treated and control populations differ, some individuals might have extreme propensity scores, which in turn leads to very high inverse-propensity weights. When using OW the weight for each individual is defined as *w*_*i*_ = 1 − *e*_*i*_ for a treated unit and *w*_*i*_ = *e*_*i*_ for a control unit ^26^. The construction of these weights allows up-weighing of individuals that were equally likely to be treated\untreated, and smoothly down-weighing individuals in the tails of the propensity score distribution. This allows for a weighted population that more closely emulated a target trial population.

**Figure S5.5:**
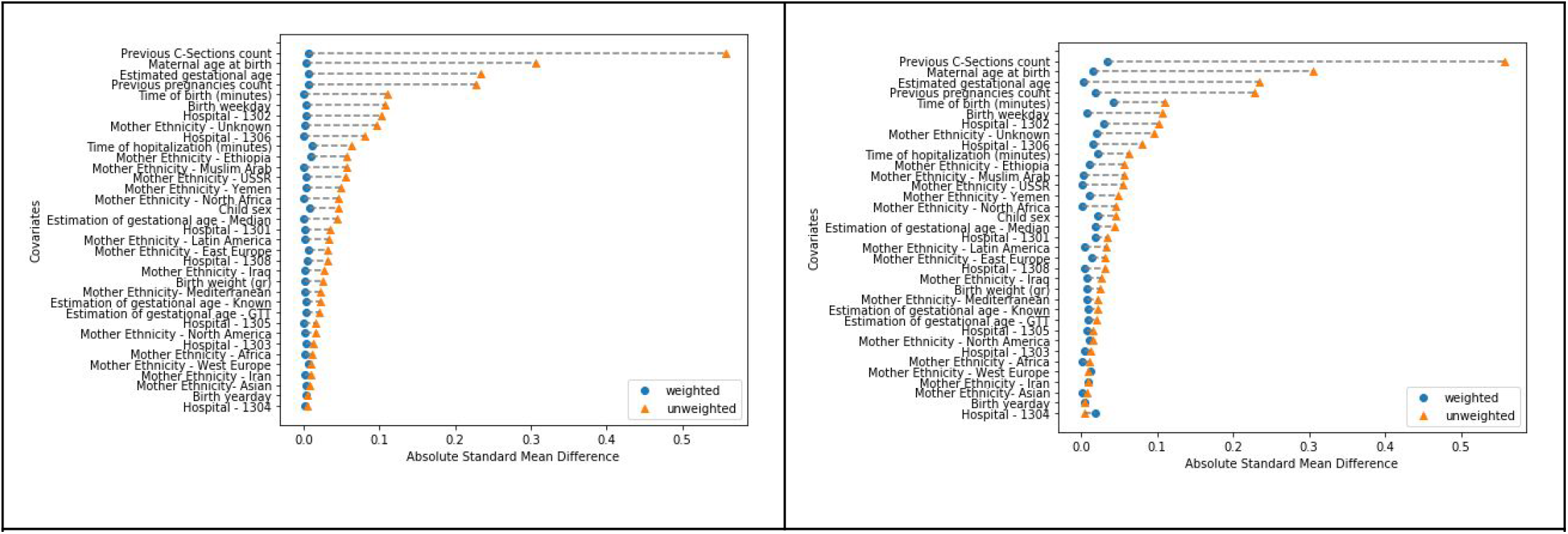
Covariates balance using Overlap Weights (OW) (left) or Inverse-Probability-weighting (IPW) (right).

## 6. Expanded results and sensitivity analysis

**Figure S6.1:**
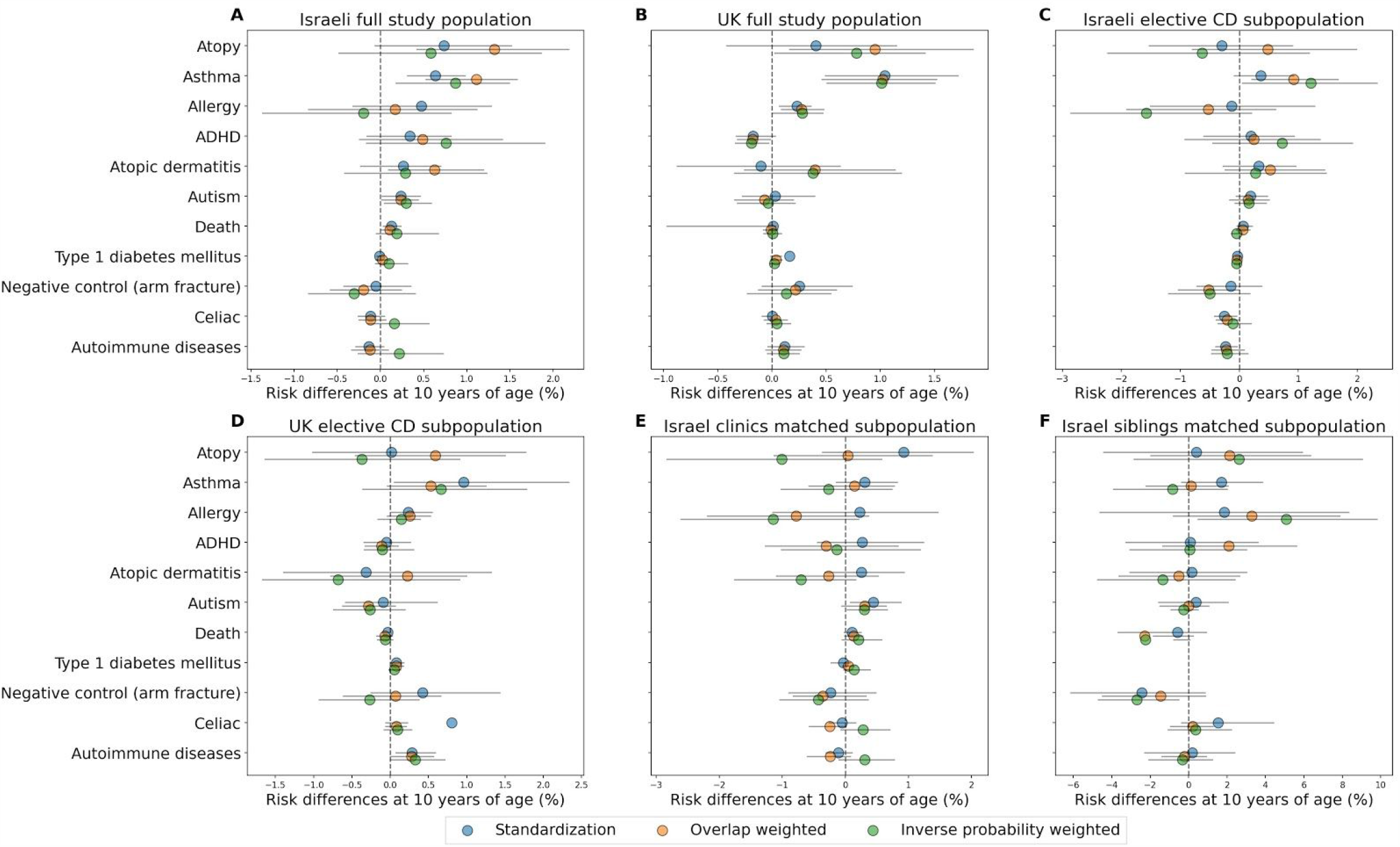
Estimated 10-yr risk difference (%) between children born vaginally or through CD. Results are shown when using Standardization (blue circles), weighting with Overlap weights (orange circles) and Weighting with Inverse-Probability-Weighting (green circles), sorted by standardized risk differences in the Israel full cohort. Black lines represent 95% confidence intervals. **A:** Israel full cohort results **B:** UK full cohort results **C:** Israel elective CD subpopulation results **D:** UK elective CD subpopulation results **E:** Israel clinics matched subpopulation results **F:** Israel siblings matched subpopulation results.

**Figure S6.2:**
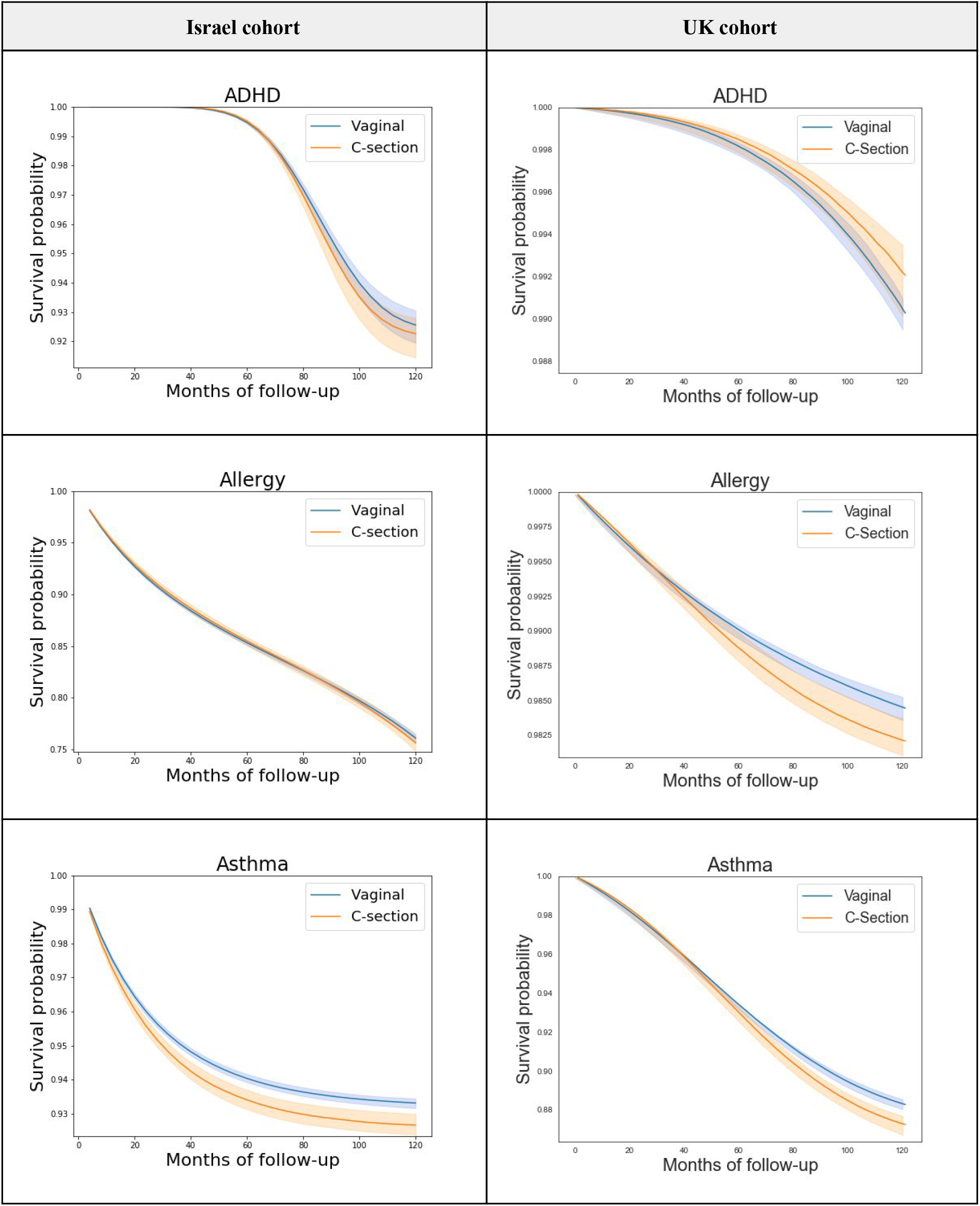
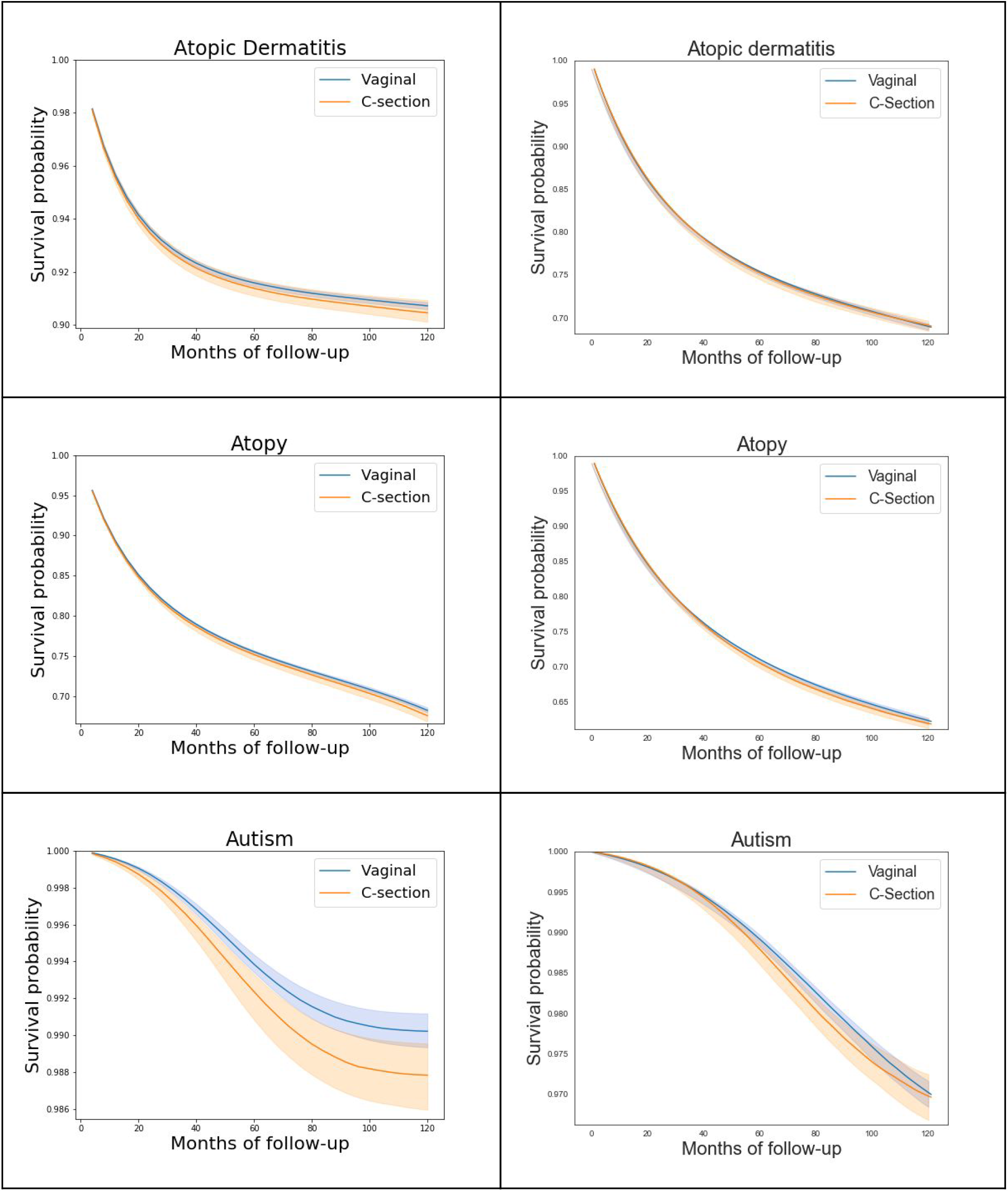
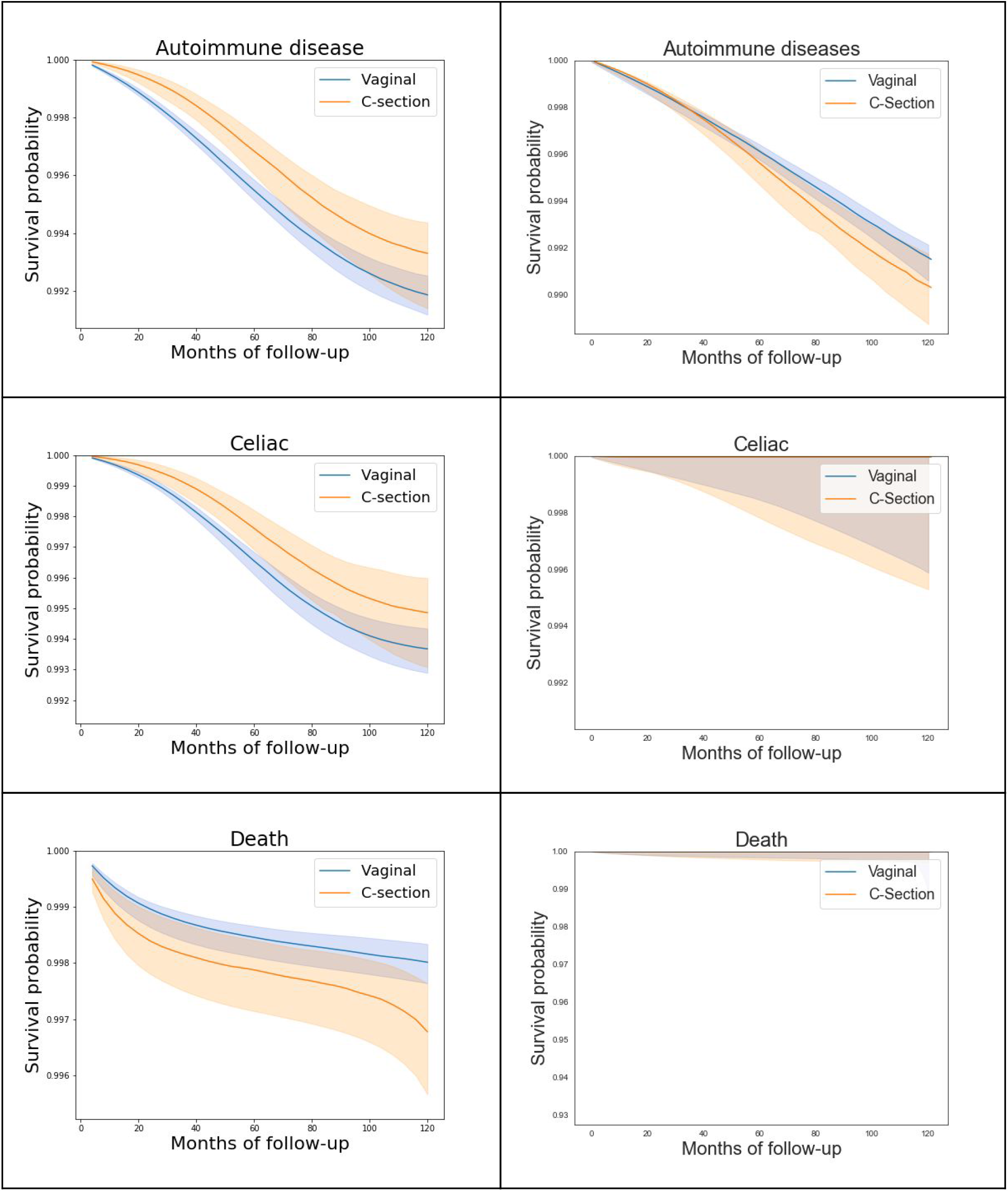
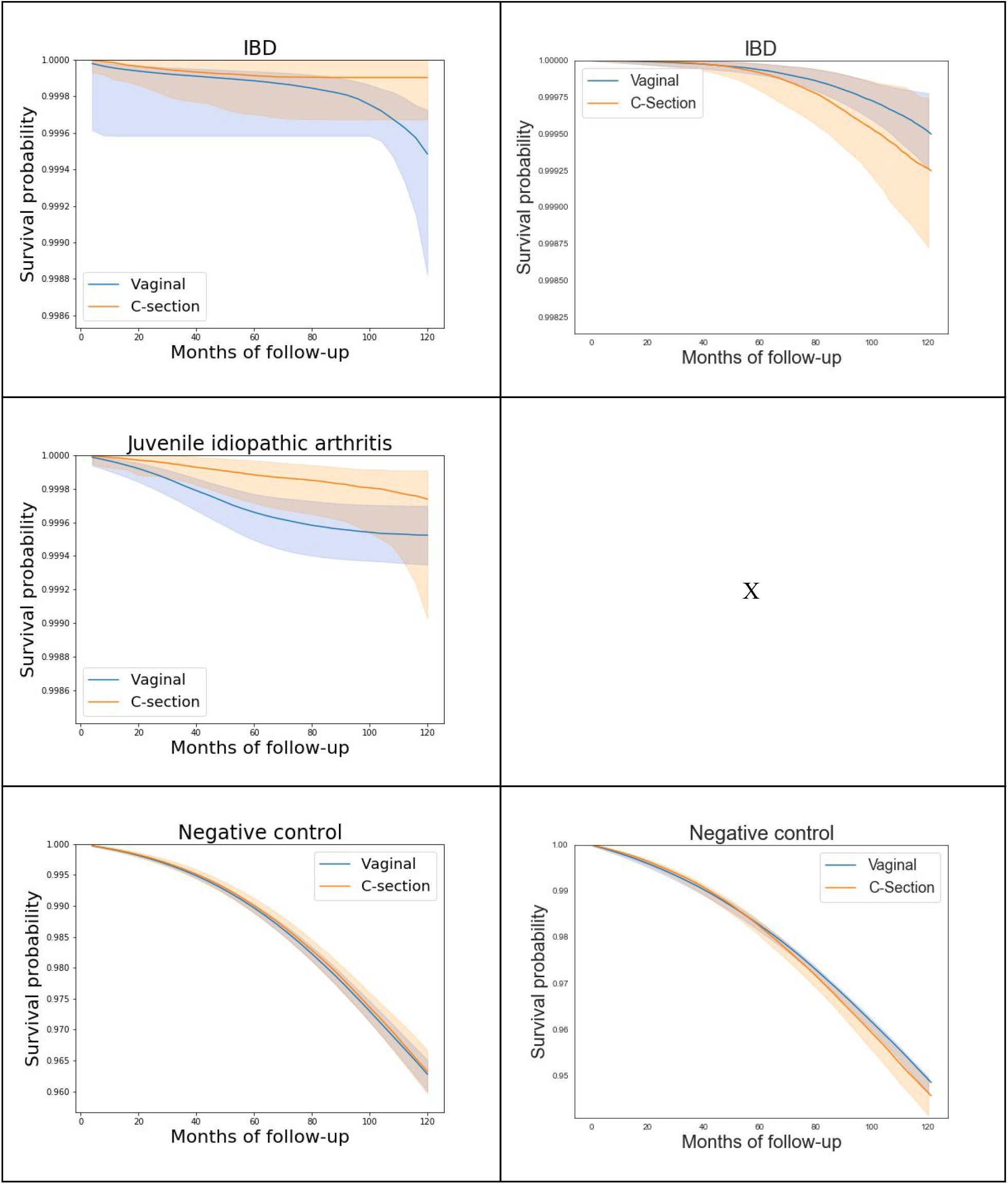
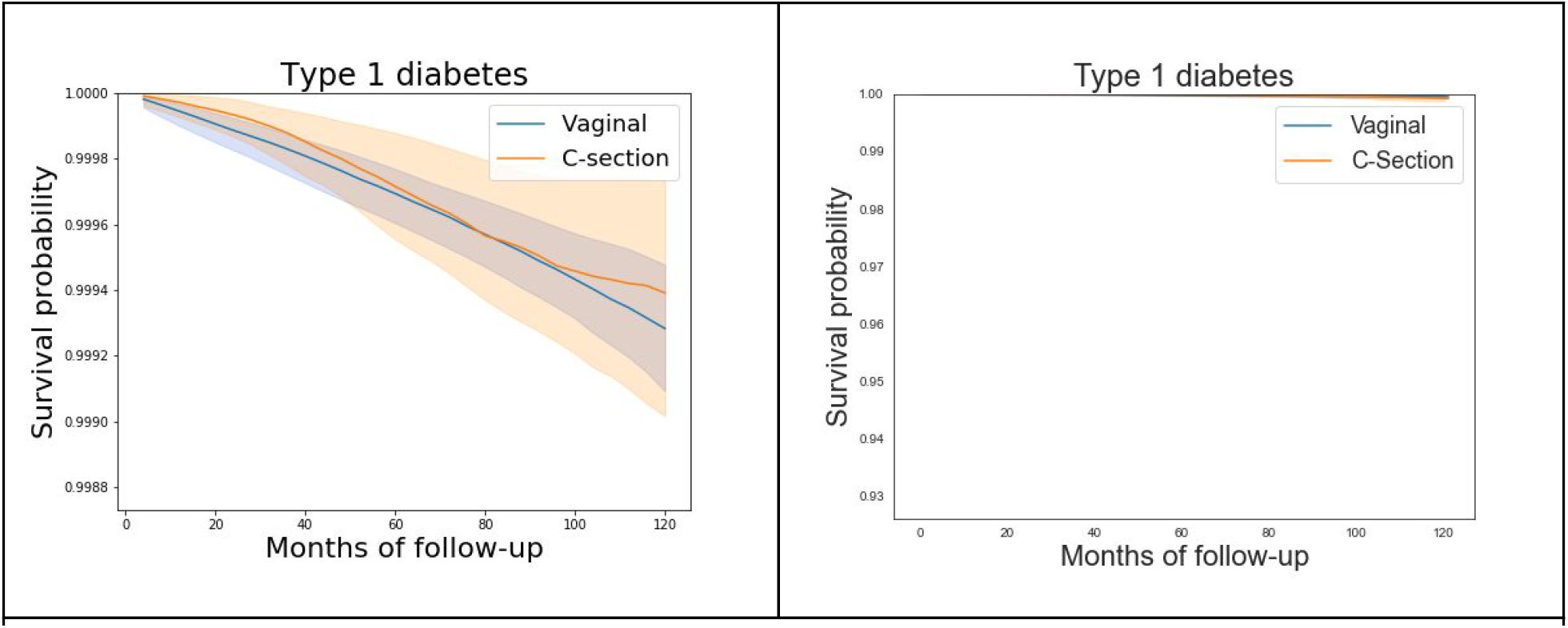
Childhood disease-free survival curves comparing vaginal and cesarean deliveries. Standardized survival curves with 95% CI of children born through vaginal delivery (blue) and CD (orange) are shown by months of follow-up. **Left:** Israel cohort, full study population. **Right:** UK cohort, full study population

**Table S6.1.**
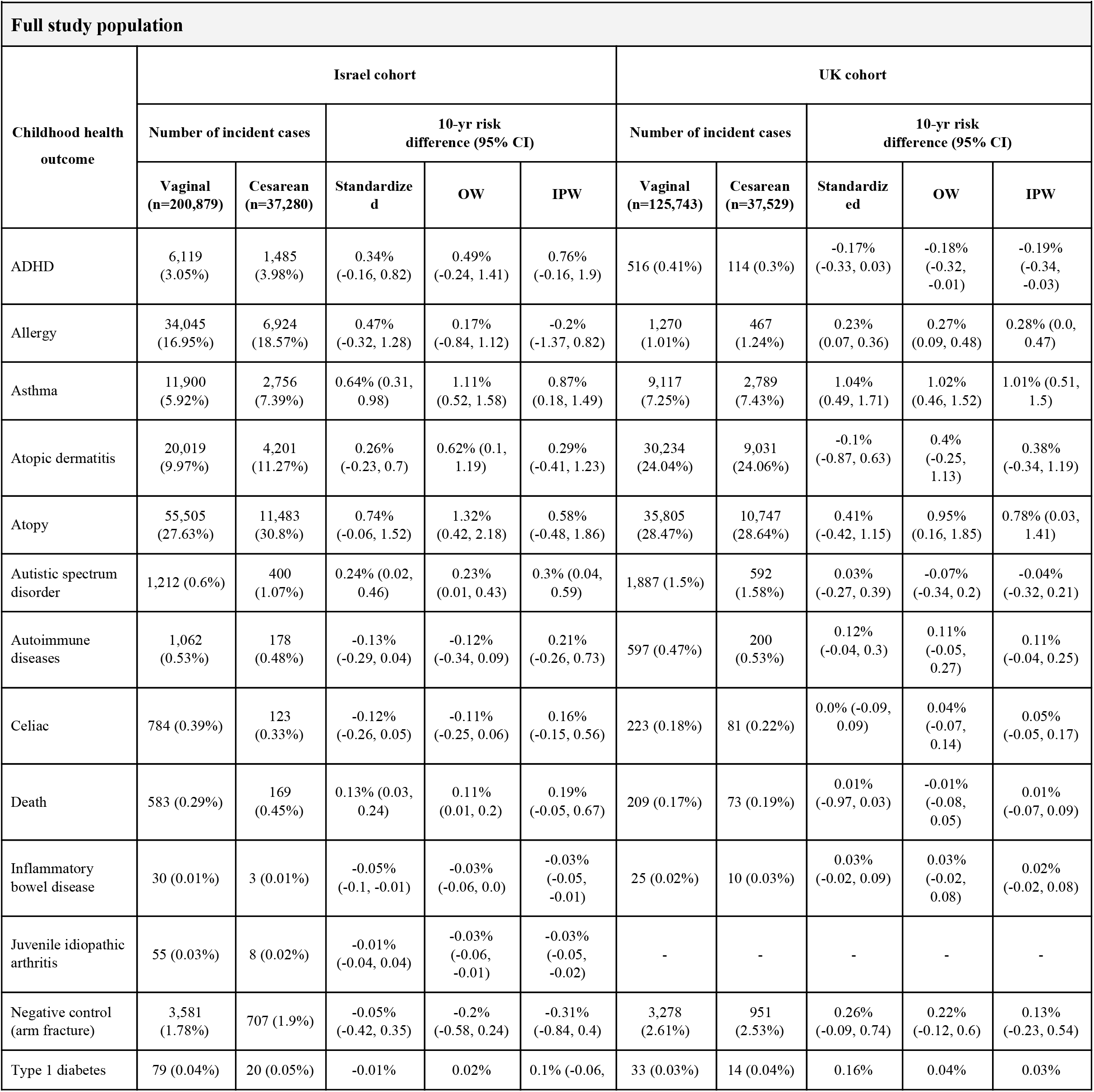
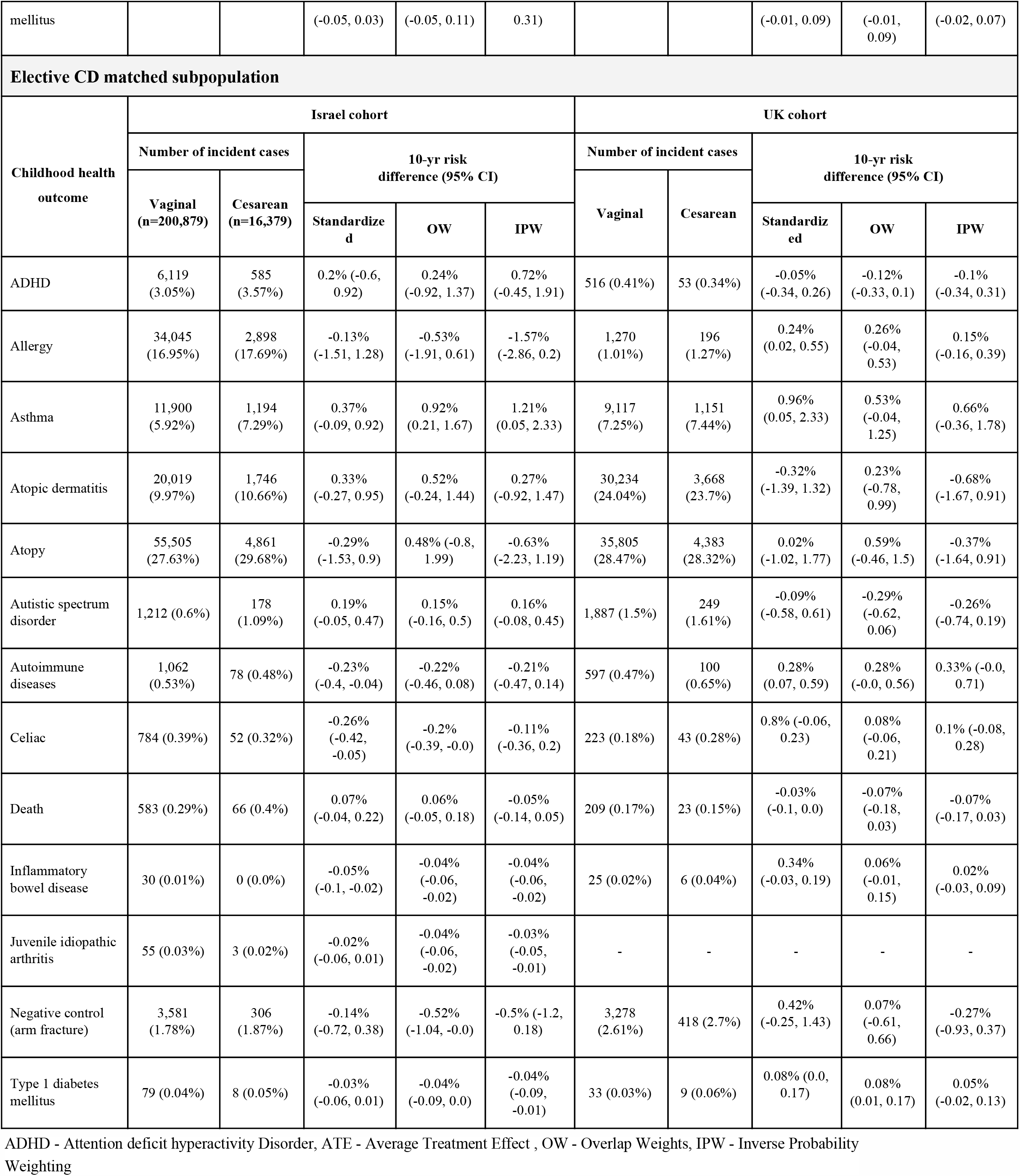
Sensitivity analysis of estimated 10-yr childhood disease-free risk differences for comparing vaginal and cesarean deliveries - standardized, OW, IPW, across two data sources

**Full study population**
| Childhood health outcome | Israel cohort |  |  |  |  | UK cohort |  |  |  |  |
| --- | --- | --- | --- | --- | --- | --- | --- | --- | --- | --- |
|  | Number of incident cases |  | 10-yr risk difference (95% CI) |  |  | Number of incident cases |  | 10-yr risk difference (95% CI) |  |  |
|  | Vaginal (n=200,879) | Cesarean (n=37,280) | Standardized | OW | IPW | Vaginal (n=125,743) | Cesarean (n=37,529) | Standardized | OW | IPW |
| ADHD | 6,119 (3.05%) | 1,485 (3.98%) | 0.34% (-0.16, 0.82) | 0.49% (-0.24, 1.41) | 0.76% (-0.16, 1.9) | 516 (0.41%) | 114 (0.3%) | -0.17% (-0.33, 0.03) | -0.18% (-0.32, -0.01) | -0.19% (-0.34, -0.03) |
| Allergy | 34,045 (16.95%) | 6,924 (18.57%) | 0.47% (-0.32, 1.28) | 0.17% (-0.84, 1.12) | -0.2% (-1.37, 0.82) | 1,270 (1.01%) | 467 (1.24%) | 0.23% (0.07, 0.36) | 0.27% (0.09, 0.48) | 0.28% (0.0, 0.47) |
| Asthma | 11,900 (5.92%) | 2,756 (7.39%) | 0.64% (0.31, 0.98) | 1.11% (0.52, 1.58) | 0.87% (0.18, 1.49) | 9,117 (7.25%) | 2,789 (7.43%) | 1.04% (0.49, 1.71) | 1.02% (0.46, 1.52) | 1.01% (0.51, 1.5) |
| Atopic dermatitis | 20,019 (9.97%) | 4,201 (11.27%) | 0.26% (-0.23, 0.7) | 0.62% (0.1, 1.19) | 0.29% (-0.41, 1.23) | 30,234 (24.04%) | 9,031 (24.06%) | -0.1% (-0.87, 0.63) | 0.4% (-0.25, 1.13) | 0.38% (-0.34, 1.19) |
| Atopy | 55,505 (27.63%) | 11,483 (30.8%) | 0.74% (-0.06, 1.52) | 1.32% (0.42, 2.18) | 0.58% (-0.48, 1.86) | 35,805 (28.47%) | 10,747 (28.64%) | 0.41% (-0.42, 1.15) | 0.95% (0.16, 1.85) | 0.78% (0.03, 1.41) |
| Autistic spectrum disorder | 1,212 (0.6%) | 400 (1.07%) | 0.24% (0.02, 0.46) | 0.23% (0.01, 0.43) | 0.3% (0.04, 0.59) | 1,887 (1.5%) | 592 (1.58%) | 0.03% (-0.27, 0.39) | -0.07% (-0.34, 0.2) | -0.04% (-0.32, 0.21) |
| Autoimmune diseases | 1,062 (0.53%) | 178 (0.48%) | -0.13% (-0.29, 0.04) | -0.12% (-0.34, 0.09) | 0.21% (-0.26, 0.73) | 597 (0.47%) | 200 (0.53%) | 0.12% (-0.04, 0.3) | 0.11% (-0.05, 0.27) | 0.11% (-0.04, 0.25) |
| Celiac | 784 (0.39%) | 123 (0.33%) | -0.12% (-0.26, 0.05) | -0.11% (-0.25, 0.06) | 0.16% (-0.15, 0.56) | 223 (0.18%) | 81 (0.22%) | 0.0% (-0.09, 0.09) | 0.04% (-0.07, 0.14) | 0.05% (-0.05, 0.17) |
| Death | 583 (0.29%) | 169 (0.45%) | 0.13% (0.03, 0.24) | 0.11% (0.01, 0.2) | 0.19% (-0.05, 0.67) | 209 (0.17%) | 73 (0.19%) | 0.01% (-0.97, 0.03) | -0.01% (-0.08, 0.05) | 0.01% (-0.07, 0.09) |
| Inflammatory bowel disease | 30 (0.01%) | 3 (0.01%) | -0.05% (-0.1, -0.01) | -0.03% (-0.06, 0.0) | -0.03% (-0.05, -0.01) | 25 (0.02%) | 10 (0.03%) | 0.03% (-0.02, 0.09) | 0.03% (-0.02, 0.08) | 0.02% (-0.02, 0.08) |
| Juvenile idiopathic arthritis | 55 (0.03%) | 8 (0.02%) | -0.01% (-0.04, 0.04) | -0.03% (-0.06, -0.01) | -0.03% (-0.05, -0.02) | - | - | - | - | - |
| Negative control (arm fracture) | 3,581 (1.78%) | 707 (1.9%) | -0.05% (-0.42, 0.35) | -0.2% (-0.58, 0.24) | -0.31% (-0.84, 0.4) | 3,278 (2.61%) | 951 (2.53%) | 0.26% (-0.09, 0.74) | 0.22% (-0.12, 0.6) | 0.13% (-0.23, 0.54) |
| Type 1 diabetes | 79 (0.04%) | 20 (0.05%) | -0.01% | 0.02% | 0.1% (-0.06, | 33 (0.03%) | 14 (0.04%) | 0.16% | 0.04% | 0.03% |

| mellitus |  |  | (-0.05, 0.03) | (-0.05, 0.11) | 0.31) |  |  | (-0.01, 0.09) | (-0.01, 0.09) | (-0.02, 0.07) |
| --- | --- | --- | --- | --- | --- | --- | --- | --- | --- | --- |
| **Elective CD matched subpopulation** |  |  |  |  |  |  |  |  |  |  |
| Childhood health outcome | Israel cohort |  |  |  |  | UK cohort |  |  |  |  |
|  | Number of incident cases |  | 10-yr risk difference (95% CI) |  |  | Number of incident cases |  | 10-yr risk difference (95% CI) |  |  |
|  | Vaginal (n=200,879) | Cesarean (n=16,379) |  |  |  | Vaginal | Cesarean |  |  |  |
|  |  |  | Standardized | OW | IPW |  |  | Standardized | OW | IPW |
| ADHD | 6,119 (3.05%) | 585 (3.57%) | 0.2% (-0.6, 0.92) | 0.24% (-0.92, 1.37) | 0.72% (-0.45, 1.91) | 516 (0.41%) | 53 (0.34%) | -0.05% (-0.34, 0.26) | -0.12% (-0.33, 0.1) | -0.1% (-0.34, 0.31) |
| Allergy | 34,045 (16.95%) | 2,898 (17.69%) | -0.13% (-1.51, 1.28) | -0.53% (-1.91, 0.61) | -1.57% (-2.86, 0.2) | 1,270 (1.01%) | 196 (1.27%) | 0.24% (0.02, 0.55) | 0.26% (-0.04, 0.53) | 0.15% (-0.16, 0.39) |
| Asthma | 11,900 (5.92%) | 1,194 (7.29%) | 0.37% (-0.09, 0.92) | 0.92% (0.21, 1.67) | 1.21% (0.05, 2.33) | 9,117 (7.25%) | 1,151 (7.44%) | 0.96% (0.05, 2.33) | 0.53% (-0.04, 1.25) | 0.66% (-0.36, 1.78) |
| Atopic dermatitis | 20,019 (9.97%) | 1,746 (10.66%) | 0.33% (-0.27, 0.95) | 0.52% (-0.24, 1.44) | 0.27% (-0.92, 1.47) | 30,234 (24.04%) | 3,668 (23.7%) | -0.32% (-1.39, 1.32) | 0.23% (-0.78, 0.99) | -0.68% (-1.67, 0.91) |
| Atopy | 55,505 (27.63%) | 4,861 (29.68%) | -0.29% (-1.53, 0.9) | 0.48% (-0.8, 1.99) | -0.63% (-2.23, 1.19) | 35,805 (28.47%) | 4,383 (28.32%) | 0.02% (-1.02, 1.77) | 0.59% (-0.46, 1.5) | -0.37% (-1.64, 0.91) |
| Autistic spectrum disorder | 1,212 (0.6%) | 178 (1.09%) | 0.19% (-0.05, 0.47) | 0.15% (-0.16, 0.5) | 0.16% (-0.08, 0.45) | 1,887 (1.5%) | 249 (1.61%) | -0.09% (-0.58, 0.61) | -0.29% (-0.62, 0.06) | -0.26% (-0.74, 0.19) |
| Autoimmune diseases | 1,062 (0.53%) | 78 (0.48%) | -0.23% (-0.4, -0.04) | -0.22% (-0.46, 0.08) | -0.21% (-0.47, 0.14) | 597 (0.47%) | 100 (0.65%) | 0.28% (0.07, 0.59) | 0.28% (-0.0, 0.56) | 0.33% (-0.0, 0.71) |
| Celiac | 784 (0.39%) | 52 (0.32%) | -0.26% (-0.42, -0.05) | -0.2% (-0.39, -0.0) | -0.11% (-0.36, 0.2) | 223 (0.18%) | 43 (0.28%) | 0.8% (-0.06, 0.23) | 0.08% (-0.06, 0.21) | 0.1% (-0.08, 0.28) |
| Death | 583 (0.29%) | 66 (0.4%) | 0.07% (-0.04, 0.22) | 0.06% (-0.05, 0.18) | -0.05% (-0.14, 0.05) | 209 (0.17%) | 23 (0.15%) | -0.03% (-0.1, 0.0) | -0.07% (-0.18, 0.03) | -0.07% (-0.17, 0.03) |
| Inflammatory bowel disease | 30 (0.01%) | 0 (0.0%) | -0.05% (-0.1, -0.02) | -0.04% (-0.06, -0.02) | -0.04% (-0.06, -0.02) | 25 (0.02%) | 6 (0.04%) | 0.34% (-0.03, 0.19) | 0.06% (-0.01, 0.15) | 0.02% (-0.03, 0.09) |
| Juvenile idiopathic arthritis | 55 (0.03%) | 3 (0.02%) | -0.02% (-0.06, 0.01) | -0.04% (-0.06, -0.02) | -0.03% (-0.05, -0.01) | - | - | - | - | - |
| Negative control (arm fracture) | 3,581 (1.78%) | 306 (1.87%) | -0.14% (-0.72, 0.38) | -0.52% (-1.04, -0.0) | -0.5% (-1.2, 0.18) | 3,278 (2.61%) | 418 (2.7%) | 0.42% (-0.25, 1.43) | 0.07% (-0.61, 0.66) | -0.27% (-0.93, 0.37) |
| Type 1 diabetes mellitus | 79 (0.04%) | 8 (0.05%) | -0.03% (-0.06, 0.01) | -0.04% (-0.09, 0.0) | -0.04% (-0.09, -0.01) | 33 (0.03%) | 9 (0.06%) | 0.08% (0.0, 0.17) | 0.08% (0.01, 0.17) | 0.05% (-0.02, 0.13) |

ADHD - Attention deficit hyperactivity Disorder, ATE - Average Treatment Effect , OW - Overlap Weights, IPW - Inverse Probability Weighting

**Table S6.2.** Sensitivity analysis of estimated 10-yr childhood disease-free risk differences for comparing vaginal and cesarean deliveries - standardized, OW, IPW

|  |  |  |  |  |  |
| --- | --- | --- | --- | --- | --- |
| Clinics matched subpopulation |  |  |  |  |  |
| Childhood health outcome | Number of incident cases |  | 10-yr risk difference (95% CI) |  |  |
|  | Vaginal (n=33,232) | Cesarean (n=33,232) |  |  |  |
|  |  |  | Standardized | OW | IPW |
| ADHD | 1,207 (3.48%) | 1,343 (4.03%) | 0.27% (-0.45, 1.24) | -0.31% (-1.27, 0.83) | -0.13% (-1.01, 1.19) |
| Allergy | 5,869 (16.92%) | 6,110 (18.35%) | 0.23% (-1.15, 1.47) | -0.78% (-2.18, 0.37) | -1.14% (-2.61, 0.21) |
| Asthma | 2,234 (6.44%) | 2,437 (7.32%) | 0.31% (-0.14, 0.82) | 0.14% (-0.58, 0.77) | -0.27% (-1.02, 0.74) |
| Atopic dermatitis | 3,614 (10.42%) | 3,752 (11.27%) | 0.25% (-0.33, 0.93) | -0.27% (-1.09, 0.52) | -0.7% (-1.76, 0.17) |
| Atopy | 9,757 (28.13%) | 10,207 (30.66%) | 0.93% (-0.36, 2.02) | 0.04% (-1.13, 1.38) | -1.01% (-2.83, 0.57) |
| Autistic spectrum disorder | 209 (0.6%) | 368 (1.11%) | 0.44% (0.08, 0.88) | 0.31% (-0.06, 0.65) | 0.3% (0.02, 0.66) |
| Autoimmune diseases | 183 (0.53%) | 166 (0.5%) | -0.11% (-0.31, 0.1) | -0.24% (-0.6, 0.08) | 0.3% (-0.11, 0.78) |
| Celiac | 136 (0.39%) | 112 (0.34%) | -0.05% (-0.26, 0.16) | -0.25% (-0.57, 0.01) | 0.28% (-0.07, 0.7) |
| Death | 81 (0.23%) | 151 (0.45%) | 0.1% (-0.02, 0.25) | 0.13% (-0.01, 0.28) | 0.21% (-0.05, 0.57) |
| Inflammatory bowel disease | 4 (0.01%) | 3 (0.01%) | 0.0% (-0.06, 0.07) | -6.0% (-99.9, 0.02) | -11.01% (-100.0, 0.01) |
| Juvenile idiopathic arthritis | 8 (0.02%) | 8 (0.02%) | -0.03% (-0.17, 0.07) | -0.02% (-0.04, 0.01) | -0.02% (-0.04, 0.0) |
| Negative control (arm fracture) | 632 (1.82%) | 628 (1.89%) | -0.23% (-0.9, 0.48) | -0.36% (-0.83, 0.32) | -0.43% (-1.04, 0.36) |
| Type 1 diabetes mellitus | 14 (0.04%) | 20 (0.06%) | -0.04% (-0.23, 0.07) | 0.04% (-0.04, 0.14) | 0.14% (-0.03, 0.39) |
| Siblings matched subpopulation |  |  |  |  |  |
| Childhood health outcome | Number of incident cases |  | 10-yr risk difference (95% CI) |  |  |
|  | Vaginal (n = 1,968) | Cesarean (n = 1,968) |  |  |  |
|  |  |  | Standardized | OW | IPW |
| ADHD | 59 (2.53%) | 73 (3.07%) | 0.08% (-3.27, 3.61) | 2.08% (-1.35, 5.6) | 0.07% (-3.05, 3.02) |
| Allergy | 316 (13.56%) | 362 (15.21%) | 1.85% (-4.62, 8.32) | 3.29% (-0.8, 7.86) | 5.08% (0.48, 9.81) |
| Asthma | 131 (5.62%) | 136 (5.71%) | 1.7% (-0.37, 3.84) | 0.12% (-2.22, 2.05) | -0.84% (-3.91, 2.01) |
| Atopic dermatitis | 211 (9.06%) | 219 (9.2%) | 0.17% (-3.06, 3.01) | -0.51% (-3.63, 2.65) | -1.34% (-4.75, 2.4) |
| Atopy | 560 (24.03%) | 590 (24.79%) | 0.4% (-4.43, 5.92) | 2.14% (-1.96, 6.34) | 2.61% (-2.84, 9.03) |
| Autistic spectrum disorder | 13 (0.56%) | 15 (0.63%) | 0.38% (-1.56, 2.05) | -0.01% (-1.5, 1.03) | -0.27% (-0.92, 0.5) |
| Autoimmune diseases | 16 (0.69%) | 9 (0.38%) | 0.19% (-2.29, 2.38) | -0.23% (-1.4, 0.9) | -0.33% (-2.08, 1.24) |
| Celiac | 11 (0.47%) | 7 (0.29%) | 1.52% (-0.37, 4.43) | 0.2% (-0.95, 1.26) | 0.37% (-1.07, 2.21) |
| Death | 8 (0.34%) | 14 (0.59%) | -0.58% (-3.68, 0.92) | -2.29% (-1.84, 0.24) | -2.24% (-0.78, 0.07) |
| Inflammatory bowel disease | - | - | - | - | - |
| Juvenile idiopathic arthritis | - | - | - | - | - |
| Negative control (arm fracture) | 43 (1.85%) | 33 (1.39%) | -2.43% (-6.13, 0.85) | -1.46% (-4.49, 0.87) | -2.69% (-4.71, -0.52) |
| Type 1 diabetes mellitus | - | - | - | - | - |

ADHD - Attention deficit hyperactivity Disorder, ATE - Average Treatment Effect , OW - Overlap Weights, IPW - Inverse Probability Weighting

**Table S6.3.** Estimated childhood obesity, antibiotics intake and clinic visits differences for comparing vaginal and cesarean deliveries, across two data sources

| Full study population |  |  |  |  |  |  |  |  |  |  |
| --- | --- | --- | --- | --- | --- | --- | --- | --- | --- | --- |
| Childhood health outcome | Israel cohort |  |  |  |  | UK cohort |  |  |  |  |
|  | Number of children with sufficient data |  | ATE difference |  |  | Number of children with sufficient data |  | ATE difference |  |  |
|  | Vaginal | Cesarean |  |  |  | Vaginal | Cesarean |  |  |  |
|  |  |  | Standardized | OW | IPW |  |  | Standardized | OW | IPW |
| BMI z-score at age 5-6 years old | 99,345 (83.6%) | 19,454 (16.4%) | 0.10 (0.07, 0.12) | 0.10 (0.07, 0.12) | 0.22 (0.17, 0.29) | - | - | - | - | - |
| Respiratory infections - number of incidents until 5 years of age | 107,654 (83.8%) | 20,864 (16.2%) | 0.92 (0.68, 1.14) | 0.96 (0.72, 1.21) | 1.43 (0.95, 1.89) | 70,705 (78.24%) | 19,666 (21.76%) | 0.12 (0.08, 0.15) | 0.14 (0.10, 0.17) | 0.14 (0.11, 0.17) |
| Elective CD subpopulation |  |  |  |  |  |  |  |  |  |  |
| BMI z-score at age 5-6 years old | 99,345 (92.5%) | 8,058 (7.5%) | 0.10 (0.08, 0.12) | 0.12 (0.08, 0.17) | 0.22 (0.01, 0.38) | - | - | - | - | - |
| Respiratory infections - number of incidents until 5 years of age | 107,654 (92.6%) | 8,577 (7.4%) | 1.11 (0.91, 1.35) | 1.02 (0.62, 1.44) | 1.63 (0.24, 3.81) | 70,705 (89.63%) | 8,181 (10.37%) | 0.10 (0.06, 0.15) | 0.12 (0.07, 0.17) | 0.11 (0.06, 0.16) |

BMI - Body Mass Index, ATE - Average Treatment Effect , OW - Overlap Weights, IPW - Inverse Probability Weighting
BMI z-score calculated according to 2000 CDC Growth Charts for the United States <sup>36</sup>

**Table S6.4.**
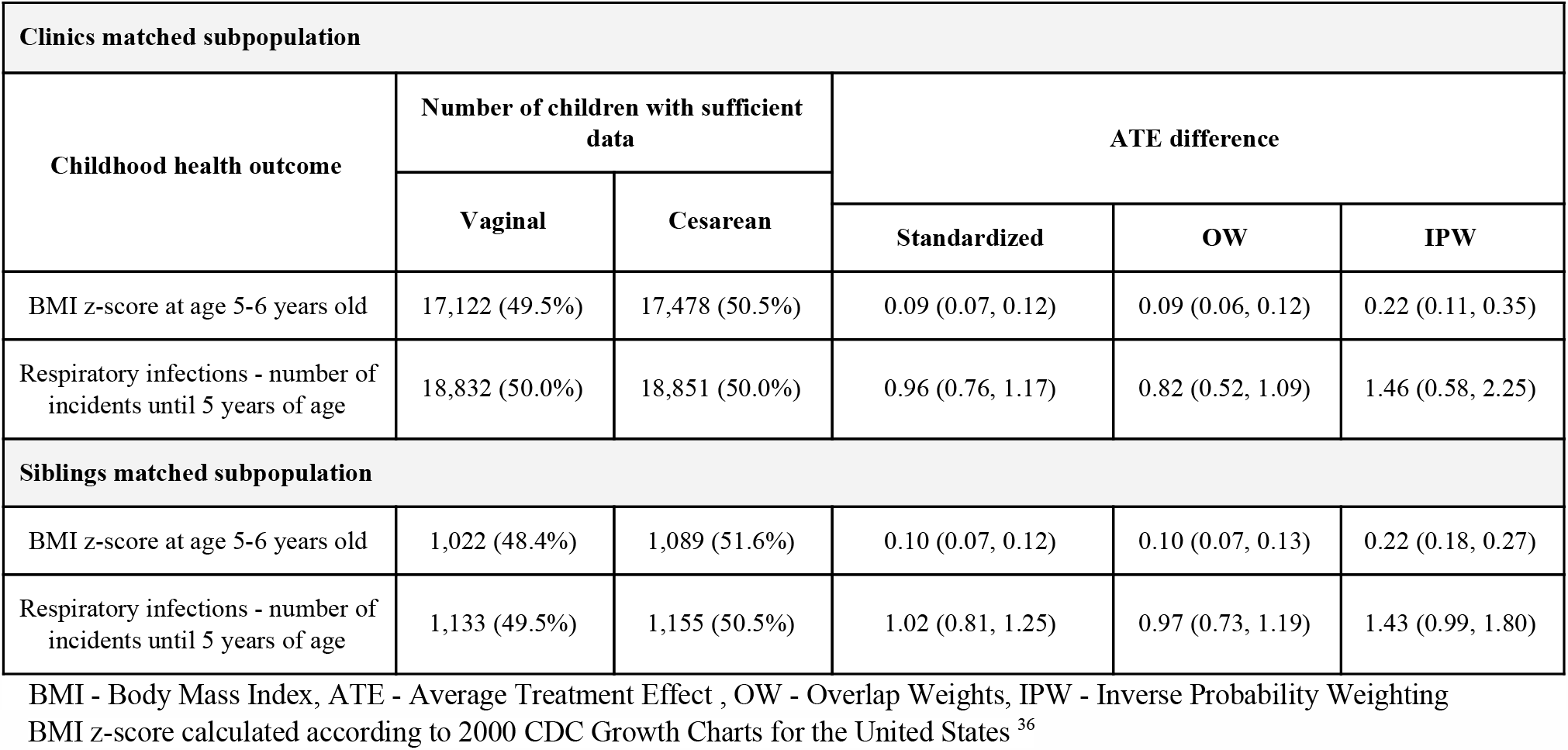
Estimated childhood obesity, antibiotics intake and clinic visits differences for comparing vaginal and cesarean deliveries.

BMI - Body Mass Index, ATE - Average Treatment Effect , OW - Overlap Weights, IPW - Inverse Probability Weighting BMI z-score calculated according to 2000 CDC Growth Charts for the United States <sup>36</sup>
|  |  |  |  |  |  |
| --- | --- | --- | --- | --- | --- |
| Clinics matched subpopulation |  |  |  |  |  |
| Childhood health outcome | Number of children with sufficient data |  | ATE difference |  |  |
|  | Vaginal | Cesarean |  |  |  |
|  |  |  | Standardized | OW | IPW |
| BMI z-score at age 5-6 years old | 17,122 (49.5%) | 17,478 (50.5%) | 0.09 (0.07, 0.12) | 0.09 (0.06, 0.12) | 0.22 (0.11, 0.35) |
| Respiratory infections - number of incidents until 5 years of age | 18,832 (50.0%) | 18,851 (50.0%) | 0.96 (0.76, 1.17) | 0.82 (0.52, 1.09) | 1.46 (0.58, 2.25) |
| Siblings matched subpopulation |  |  |  |  |  |
| BMI z-score at age 5-6 years old | 1,022 (48.4%) | 1,089 (51.6%) | 0.10 (0.07, 0.12) | 0.10 (0.07, 0.13) | 0.22 (0.18, 0.27) |
| Respiratory infections - number of incidents until 5 years of age | 1,133 (49.5%) | 1,155 (50.5%) | 1.02 (0.81, 1.25) | 0.97 (0.73, 1.19) | 1.43 (0.99, 1.80) |

BMI - Body Mass Index, ATE - Average Treatment Effect , OW - Overlap Weights, IPW - Inverse Probability Weighting
BMI z-score calculated according to 2000 CDC Growth Charts for the United States<sup>36</sup>

### Elective CD subpopulation

**Table S6.5.** Baseline characteristics of the elective CD subpopulation - Israel cohort.

|  |  |  |  |
| --- | --- | --- | --- |
| Characteristic, mean (SD) or counts % | Vaginal (n = 200,879)<br>(92.5%) | Cesarean (n = 16,379)<br>(7.5%) | All (n = 217,258) |
| **Neonatal characteristics** |  |  |  |
| Sex - Male | 102,717 (51.13%) | 8,633 (52.71%) | 111,350 (51.25%) |
| Birth weight (gr) | 3,291.02 (391.48) | 3,277.03 (451.44) | 3,289.96 (396.33) |
| Gestational age at delivery (weeks) | 39.32 (1.14) | 38.13 (0.79) | 39.23 (1.16) |
| **Maternal characteristics** |  |  |  |
| Maternal age (years) | 29.78 (5.34) | 33.12 (5.16) | 30.03 (5.4) |
| Maternal weight pre-pregnancy (Kg) | 62.37 (12.78) | 68.54 (15.93) | 62.86 (13.17) |
| Maternal height (meters) | 1.62 (0.06) | 1.62 (0.07) | 1.62 (0.06) |
| Diabetes Mellitus | 2,461 (1.23%) | 520 (3.17%) | 2,981 (1.37%) |
| Chronic Hypertension | 1,642 (0.82%) | 443 (2.7%) | 2,085 (0.96%) |
| Gravidity | 0.77 (0.81) | 0.63 (0.62) | 0.76 (0.8) |
| Previous cesarean deliveries | 6,604 (3.29%) | 7,123 (43.49%) | 13,727 (6.32%) |
| Sister's cesarean deliveries, percent* | 12.25 (27.98) | 19.24 (35.12) | 12.66 (28.5) |
| **Pregnancy characteristics** |  |  |  |
| Gestational Diabetes | 9,439 (4.7%) | 1,785 (10.9%) | 11,224 (5.17%) |
| Gestational Hypertension | 1,867 (0.93%) | 253 (1.54%) | 2,120 (0.98%) |
| Gestational weight gain z-score | -0.51 (2.42) | -0.36 (2.54) | -0.49 (2.43) |

Gestational weight gain z-score was calculated according to <sup>64</sup>
\*Sisters CD percent was calculated as the percentage of CDs over all sister's pregnancies which ended prior to the relevant date of birth.

**Table S6.6.** Baseline characteristics of the elective CD subpopulation - UK cohort.

|  |  |  |  |
| --- | --- | --- | --- |
| Characteristic, mean (SD) or counts % | Vaginal (n = 125,743)<br>(89.04%) | Cesarean (n = 15,479)<br>(10.96%) | All (n =141,222) |
| **Neonatal characteristics** |  |  |  |
| Sex - Male | 63,755 (50.70%) | 7,964 (51.45%) | 71,719 (50.78%) |
| Birth weight (gr) (N=58,695) | 3,478 (453) | 3,473 (487) | 3,478 (457) |
| Gestational age at delivery (weeks)<br>(N=137,011) | 40.24 (1.28) | 39.31 (1.20) | 40.15 (1.30) |
| **Maternal characteristics** |  |  |  |
| Maternal age (years) | 30.32 (5.59) | 32.92 (5.15) | 30.61 (5.57) |
| Maternal weight pre-pregnancy (Kg)<br>(N=44,520) | 69.09 (16.36) | 73.68 (18.32) | 69.60 (16.65) |
| Maternal height (meters) (N=133,343) | 1.65 (0.07) | 1.63 (0.07) | 1.64 (0.07) |
| Diabetes Mellitus | 730 (0.58%) | 328 (2.12%) | 1058 (0.75%) |
| Chronic Hypertension | 528 (0.42%) | 138 (0.89%) | 666 (0.47%) |
| Gravidity | 0.82 (1.21) | 1.01 (1.28) | 0.84 (1.22) |
| Previous cesarean deliveries | 3206 (2.55%) | 5085 (32.85%) | 8291 (5.87%) |
| Sister's cesarean deliveries, percent* | - | - | - |
| **Pregnancy characteristics** |  |  |  |
| Gestational Diabetes | 1699 (1.35%) | 483 (3.12%) | 2182 (1.55%) |
| Gestational Hypertension | 1,248 (0.99%) | 243 (1.57%) | 1491 (1.06%) |
| Gestational weight gain z-score | - | - | - |

Gestational weight gain z-score was calculated according to <sup>64</sup>
\*Sisters CD percent was calculated as the percentage of CDs over all sister's pregnancies which ended prior to the relevant date of birth.

### Clinics matched subpopulation

**Table S6.7.** Baseline characteristics of the clinics matched subpopulation.

|  |  |  |  |
| --- | --- | --- | --- |
| **Characteristic, mean (SD) or counts %** | **Vaginal (n = 33,232) (50.0%)** | **Cesarean (n = 33,232) (50.0%)** | **All (n = 66,464)** |
| **Neonatal characteristics** |  |  |  |
| Sex - Male | 16,910 (50.88%) | 18,072 (54.38%) | 34,982 (52.63%) |
| Birth weight (gr) | 3,294.54 (392.77) | 3,306.15 (460.67) | 3,300.34 (428.11) |
| Gestational age at delivery (weeks) | 39.29 (1.14) | 38.73 (1.24) | 39.01 (1.22) |
| **Maternal characteristics** |  |  |  |
| Maternal age (years) | 30.2 (5.14) | 32.12 (5.4) | 31.16 (5.36) |
| Maternal weight pre-pregnancy (Kg) | 62.46 (12.87) | 66.94 (15.57) | 64.78 (14.5) |
| Maternal height (meters) | 1.62 (0.06) | 1.61 (0.07) | 1.62 (0.06) |
| Diabetes Mellitus | 423 (1.27%) | 837 (2.52%) | 1,260 (1.9%) |
| Chronic Hypertension | 289 (0.87%) | 725 (2.18%) | 1,014 (1.53%) |
| Gravidity | 0.72 (0.78) | 0.54 (0.65) | 0.63 (0.72) |
| Previous cesarean deliveries | 1,063 (3.2%) | 10,433 (31.39%) | 11,496 (17.3%) |
| Sister's cesarean deliveries, percent* | 12.94 (29.29) | 18.75 (34.37) | 15.7 (31.93) |
| **Pregnancy characteristics** |  |  |  |
| Gestational Diabetes | 1,655 (4.98%) | 3,207 (9.65%) | 4,862 (7.32%) |
| Gestational Hypertension | 336 (1.01%) | 520 (1.56%) | 856 (1.29%) |
| Gestational weight gain z-score | -0.47 (2.42) | -0.35 (2.56) | -0.4 (2.5) |

Gestational weight gain z-score was calculated according to <sup>64</sup>
\*Sisters CD percent was calculated as the percentage of CDs over all sister's pregnancies which ended prior to the relevant date of birth.

### Siblings matched subpopulation

**Table S6.8.** Baseline characteristics of the siblings matched subpopulation

|  |  |  |  |
| --- | --- | --- | --- |
| Characteristic, mean (SD) or counts % | Vaginal (n = 1,968) (50.0%) | Cesarean (n = 1,968) (50.0%) | All (n = 3,936) |
| **Neonatal characteristics** |  |  |  |
| Sex - Male | 1,119 (56.86%) | 1,119 (56.86%) | 2,238 (56.86%) |
| Birth weight (gr) | 3,314.51 (395.12) | 3,286.42 (468.41) | 3,300.47 (433.49) |
| Gestational age at delivery (weeks) | 39.4 (1.15) | 39.04 (1.28) | 39.22 (1.23) |
| **Maternal characteristics** |  |  |  |
| Maternal age (years) | 29.99 (4.93) | 29.95 (5.42) | 29.97 (5.18) |
| Maternal weight pre-pregnancy (Kg) | 63.5 (13.44) | 63.55 (13.51) | 63.53 (13.47) |
| Maternal height (meters) | 1.61 (0.06) | 1.61 (0.07) | 1.61 (0.06) |
| Diabetes Mellitus | 36 (1.83%) | 31 (1.58%) | 67 (1.7%) |
| Chronic Hypertension | 25 (1.27%) | 17 (0.86%) | 42 (1.07%) |
| Gravidity | 0.94 (0.66) | 0.65 (0.78) | 0.8 (0.74) |
| Previous cesarean deliveries | 1,061 (53.91%) | 24 (1.22%) | 1,085 (27.57%) |
| Sister's cesarean deliveries, percent* | 14.25 (30.57) | 15.83 (31.38) | 15.03 (30.97) |
| **Pregnancy characteristics** |  |  |  |
| Gestational Diabetes | 99 (5.03%) | 153 (7.77%) | 252 (6.4%) |
| Gestational Hypertension | 20 (1.02%) | 22 (1.12%) | 42 (1.07%) |
| Gestational weight gain z-score | -0.56 (2.55) | -0.43 (2.62) | -0.48 (2.59) |

Gestational weight gain z-score was calculated according to <sup>64</sup>
\*Sisters CD percent was calculated as the percentage of CDs over all sister's pregnancies which ended prior to the relevant date of birth.

## 7. Elective CD classification model

### Elective CD classification model parameters

We used a gradient boosting trees model trained with the XGBOOST python package. Hyperparameters were selected with the following settings:

- nuestimators= 50
- max_depth= 3

### Elective CD classification model evaluation

**Figure S7.1.**
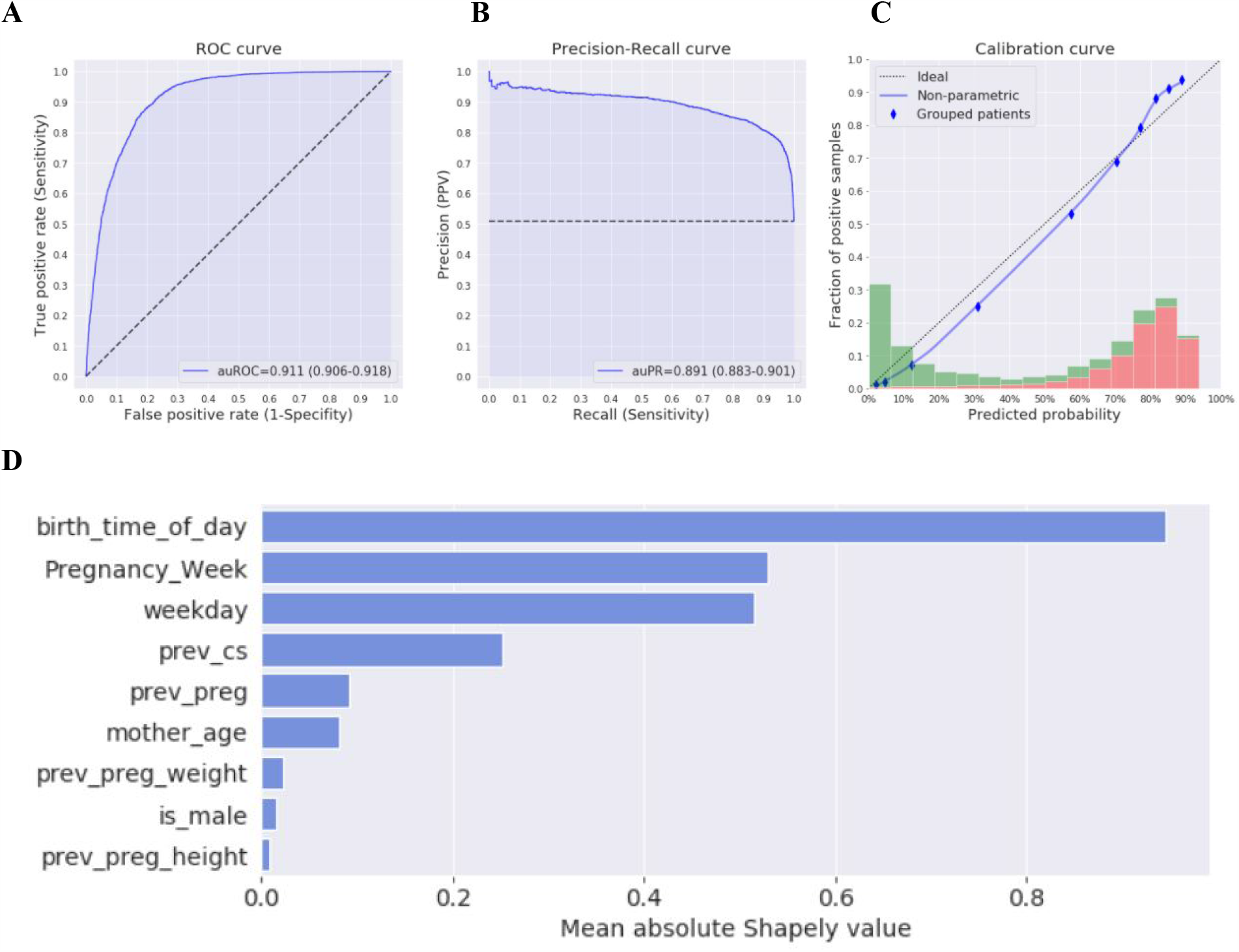
Elective CD classification model. **A:** Receiver operating characteristic (ROC) curve of our model. **B:** Precision-Recall curve of our model. **C:** Calibration curve. Blue dots represent deciles of predicted probabilities. Dotted diagonal line represents an ideal calibration. Histogram at the bottom: predicted probabilities of children born in emergency CD (green) and elective CD (red). **D:** Mean absolute Shapley values (in log-odds scale) of the model features.

**Figure S7.2.**
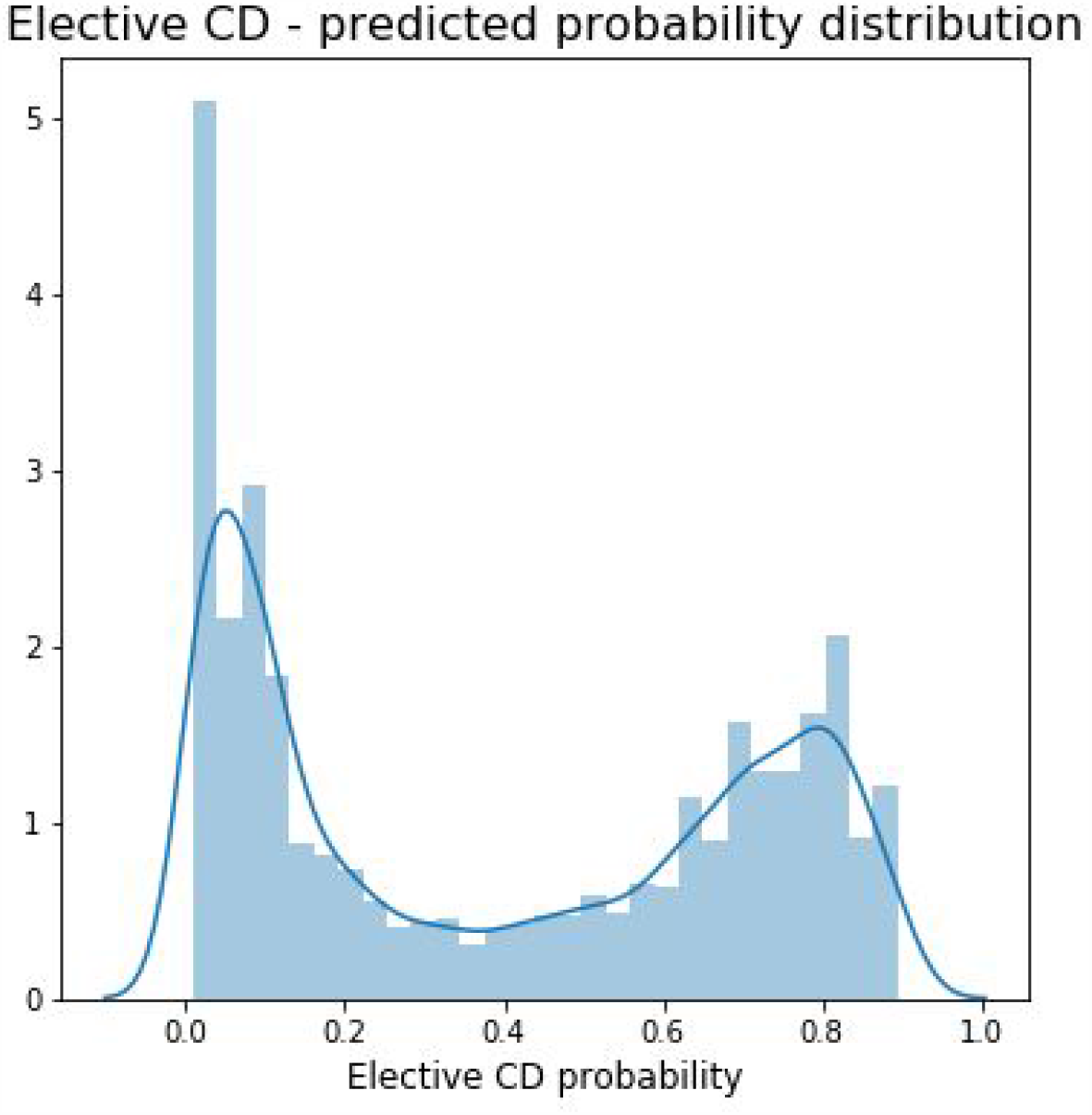
Elective CD model in Clalit EHRs. Elective CD probability distribution as predicted by the model on Clalit EHRs database. CDs with probability of 0.5 and higher were considered as elective.

## 8. DAGs for CD and pediatric health

**Figure S8.1:**
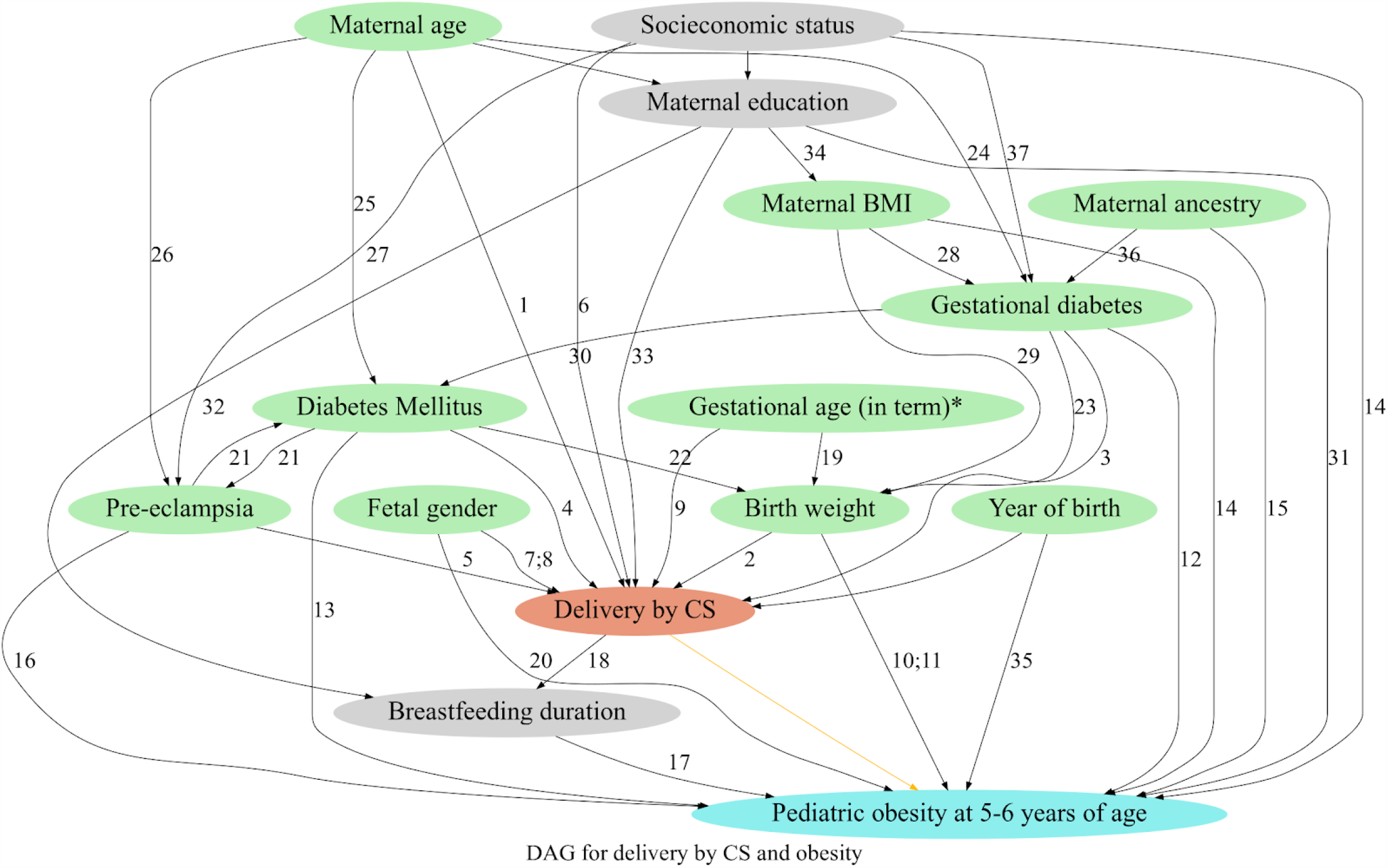
Directed acyclic graph (DAG) for cesarean delivery and pediatric obesity at 5-6 years of age. DAG derived from literature and expert knowledge - nodes represent variables and arrows represent causal associations. Exposure, delivery by CS, colored in coral. Outcome, pediatric obesity at 5-6 years of age, colored in light blue. Variable nodes are colored in light green if they were available in our data, and light grey otherwise. *Our study includes only infants born in term delivery and therefore the factors that are associated with preterm birth are not accounted for. Numbers represent available sources of literature describing the associations, references for these associations are given in section 8 of the supplementary appendix.

